# Priming of Multiple HIV Neutralizing B Cell Precursors in Humans

**DOI:** 10.64898/2026.05.02.26352058

**Authors:** Chen-Hao Yeh, Stephen R. Walsh, Ruth J. Parsons, Qianyi E. Zhang, Bhishem Thakur, Kijun Song, Kejun Liu, Hélène Fradin Kirshner, Elizabeth Van ltallie, Matthew Clark, William O. Hahn, Shengli Song, Liwen Lin, Hsin-I Huang, Jade Lugo, Alex Quan, Yue Xue, Yue Chen, Ryan Tuck, Katayoun Mansouri, Taylor N. Spence, Helena A. Laukaitis, Rob Parks, Margaret Barr, Elise Schweer, Nicholas Levering, Amanda Eaton, Shaunna Shen, Katarzyna Janowska, Victor Ilevbare, Graham Johnson, Peng Wang, Derek Cain, Aria Arus-Altuz, Elizabeth Donahue, Ilya Zhbannikov, Madison Berry, Sravani Venkatayogi, Joshua S. Martin Beem, Ollivier Hyrien, Pei-Chun Yu, Laura L. Polakowski, India Tindale, Claudio Yurdadon, Randy Burnham, Jessica Andriesen, Kshitij Wagh, Michael S. Seaman, Emmanuel B. Walter, Garnett Kelsoe, M. Juliana McElrath, Lawrence Corey, Kenneth Mayer, Persephone Borrow, Wilton Williams, Kevin O. Saunders, Kevin Wiehe, David Montefiori, Robert J. Edwards, Priyamvada Acharya, Lindsey Baden, Barton F. Haynes

**Affiliations:** Duke Human Vaccine Institute, Duke University School of Medicine, Durham, NC, 27710, USA; Department of Medicine, Duke University School of Medicine, Durham, NC 27710, USA; Mass General Brigham, Boston, MA 02119, USA; Harvard Medical School, Boston, MA 02115, USA; Department of Surgery, Duke University School of Medicine, Durham, NC 27710, USA; Department of Integrative Immunobiology, Duke University School of Medicine, Durham, NC 27710, USA; Department of Biochemistry, Duke University School of Medicine, Durham, NC 27710, USA; Fred Hutchinson Cancer Center, Seattle, WA 98109, USA; National Institute of Allergy and Infectious Diseases; Department of Medicine, Division of Allergy and Infectious Disease, University of Washington, Seattle, WA 98195, USA; Beth Israel Deaconess Medical Center, Boston, MA 02215, USA; The Fenway Institute, Boston, MA 02215, USA; Centre for Immuno-Oncology, Nuffield Department of Clinical Medicine, University of Oxford, Oxford, OX3 7DQ, United Kingdom; Department of Molecular Genetics and Microbiology, Duke University School of Medicine, Durham, NC 27710, USA

**Keywords:** Human Immunodeficiency Virus, B cell, transmitted/founder virus, neutralizing antibody, vaccine, clinical trial

## Abstract

Induction of broadly neutralizing antibodies (bnAbs) remains a major goal of HIV vaccine development. In a Phase I clinical trial, we evaluated the immunogenicity of CH505 transmitted/founder (TF) envelope trimer designed to prime precursors of the heavy chain third complementarity determining region (CDRH3)-dominant class of CD4-binding site (CD4bs) HIV bnAbs. Neutralizing memory B-cell lineages were isolated from all vaccine recipients, including lineages targeting the CD4bs, V1/V3, and gp120/gp41 interface. Candidate CDRH3-dominant, CD4bs bnAb precursor lineages were identified in 73% of vaccinees, including one lineage that exhibited limited heterologous neutralization of global HIV-1 isolates. Structural analyses defined the modes of Env recognition by these three antibody classes. Together, these findings demonstrate that immunization with the CH505 TF trimer can prime a diverse, polyclonal repertoire of bnAb precursor lineages in humans, providing multiple starting points for sequential immunization strategies aimed at eliciting broadly neutralizing HIV-1 antibodies.

**HIGHLIGHTS:**

- CH505 TF trimer immunogen induced autologous tier-2 neutralizing B cell lineages in all vaccine recipients.
- S365P-sensitive, CDRH3-dominant CD4-binding-site bnAb precursor candidates were detected in 73% of vaccinees.
- A single immunogen primed neutralizing antibody lineages targeting the CD4bs, V1/V3, and gp120/gp41 interface.
- High S365P-sensitive CD4bs responses were associated with distinct NK- and CD8⁺ T-cell states.

## INTRODUCTION

The induction of broadly neutralizing antibodies (bnAbs) remains a major goal of HIV vaccine development ^1^. Among conserved envelope (Env) epitopes, the CD4 binding site (CD4bs) is attractive due to its functional significance in mediating HIV infection ^2,3^. A rational framework for initiating bnAb lineages has emerged, in which priming immunogens are engineered to engage the subset of naïve B cells with features essential for eventual maturation to broad neutralization, followed by boosting with sequential Envs with increasing affinity gradients to guide B cell maturation toward breadth ^4–6^. Four human clinical trials have demonstrated vaccine-induced elicitation of candidate precursors of the VRC01 class of CD4bs bnAbs that use CDRH2 to mimic the β-strand interaction between the CD4 receptor and the CD4-binding loop in the gp120 subunit of Env ^7–9^. However, VRC01-class neutralizing antibodies typically require extensive somatic hypermutation and often acquire improbable mutations, insertions or deletions to achieve broad and potent neutralization ^3,10–13^. Nonetheless, recent clinical studies have demonstrated that neutralizing bnAb lineages can be successfully initiated and affinity matured in humans. The HVTN 133 trial targeting the membrane-proximal external region (MPER) has shown that naïve B cell-targeting immunogens can expand candidate bnAb precursors, initiate early somatic hypermutation, and affinity-mature to neutralize 41% of clade B and 14% of global HIV isolates ^14,15^.

A second type of CD4bs bnAbs utilizes a CDRH3-dominant mode of interaction with gp120, distinct from the CDRH2-dominant, CD4-mimicking, VRC01-class of CD4bs bnAbs ^16–18^. While typically less potent and broad than the VRC01-class of CD4bs bnAbs ^19^, the CDRH3-dominant class of CD4bs bnAbs requires fewer mutations for bnAb breadth, which may lower the barriers to their elicitation and affinity maturation ^18,19^. During studies of antibody-virus coevolution in a person living with HIV (PLWH), we isolated four members of the CH103 bnAb-lineage that recognized Env through a CDRH3-dominant mode of binding ^18^. The CH103 B cell lineage relies on a tyrosine-rich CDRH3 loop for gp120 binding and achieves cross-neutralization with limited framework diversification ^1,20^. Thus, CD4bs bnAbs of the CDRH3-dominant class are desirable targets for bnAb induction.

We used lineage tracing to infer the CH103 bnAb unmutated common ancestor (UCA), which bound to the CH505 transmitted-founder (TF) ^21^ Env stabilized trimer ^22^ with an apparent affinity (K_D_^app^) of 37.5 nM ^18^ and intrinsic affinity (K_D_) of 450 nM. CH103 bnAb intermediates acquired heterologous neutralization after antibody mutations were selected by Env diversification ^20^. We proposed that an immunogen such as CH505 TF could target and expand CDRH3-dominant bnAb precursors ^4,18,23^.

The native-like CH505 SOSIP trimer used as a prime in the HVTN 300 trial (NCT04915768) was designed with the BG505 clade A Env gp41 and the CH505 gp120 with E64K and A316W mutations added for Env stability to form the TF chimeric(ch) trimer (hereafter designated CH505 TF chTrimer) ^23–25^.

Preclinical studies in rhesus macaques confirmed that CH505 TF chTrimers can induce polyclonal tier 2 autologous neutralizing antibodies that protected from homologous SHIV CH505 mucosal challenges ^24^. Notably, the CH505 TF chTrimer lacks the N332 glycan and, in macaques, can induce V3-base-targeted neutralizing antibodies that are N332 glycan-independent ^24^. The Phase 1 HVTN 300 trial (NCT04915768) was designed to translate these findings to humans ^26^.

Here, we comprehensively profiled the neutralizing memory B (Bmem) cell repertoire elicited by CH505 TF chTrimer vaccination in HVTN 300 using high-throughput single-cell cultures ^27,28^. We show that the CH505 TF chTrimer immunization elicited not only autologous neutralizing CDRH3-dominant CD4bs antibodies but also neutralizing antibodies targeting the gp120/gp41 interface region ^29,30^, and the N332-independent V3 GDIR region, with structural features resembling those of a recently described class of N332-independent V3-directed neutralizing antibodies ^31–33^. Together, these findings provide proof-of-principle for B-cell lineage-informed immunogen design^4^ and demonstrate that a single transmitted/founder Env immunogen can simultaneously prime multiple neutralizing antibody developmental pathways in humans.

## RESULTS

### CH505 TF chTrimer elicited robust autologous tier 2 neutralization in humans

HVTN 300 was a first-in-human Phase 1 trial designed to evaluate the safety and immunogenicity of the CH505 TF chTrimer immunogen. We analyzed the B-cell repertoires of all 11 participants enrolled in Part A of the HVTN 300 trial, in which vaccine recipient received up to five doses of 300 µg CH505 TF formulated with the TLR7/8 agonist 3M-052 plus alum (**Figs. 1A and S1A**) ^34^. Safety outcomes, T-cell responses, and serum autologous neutralization activity have been reported separately^26^.

**Figure 1.**
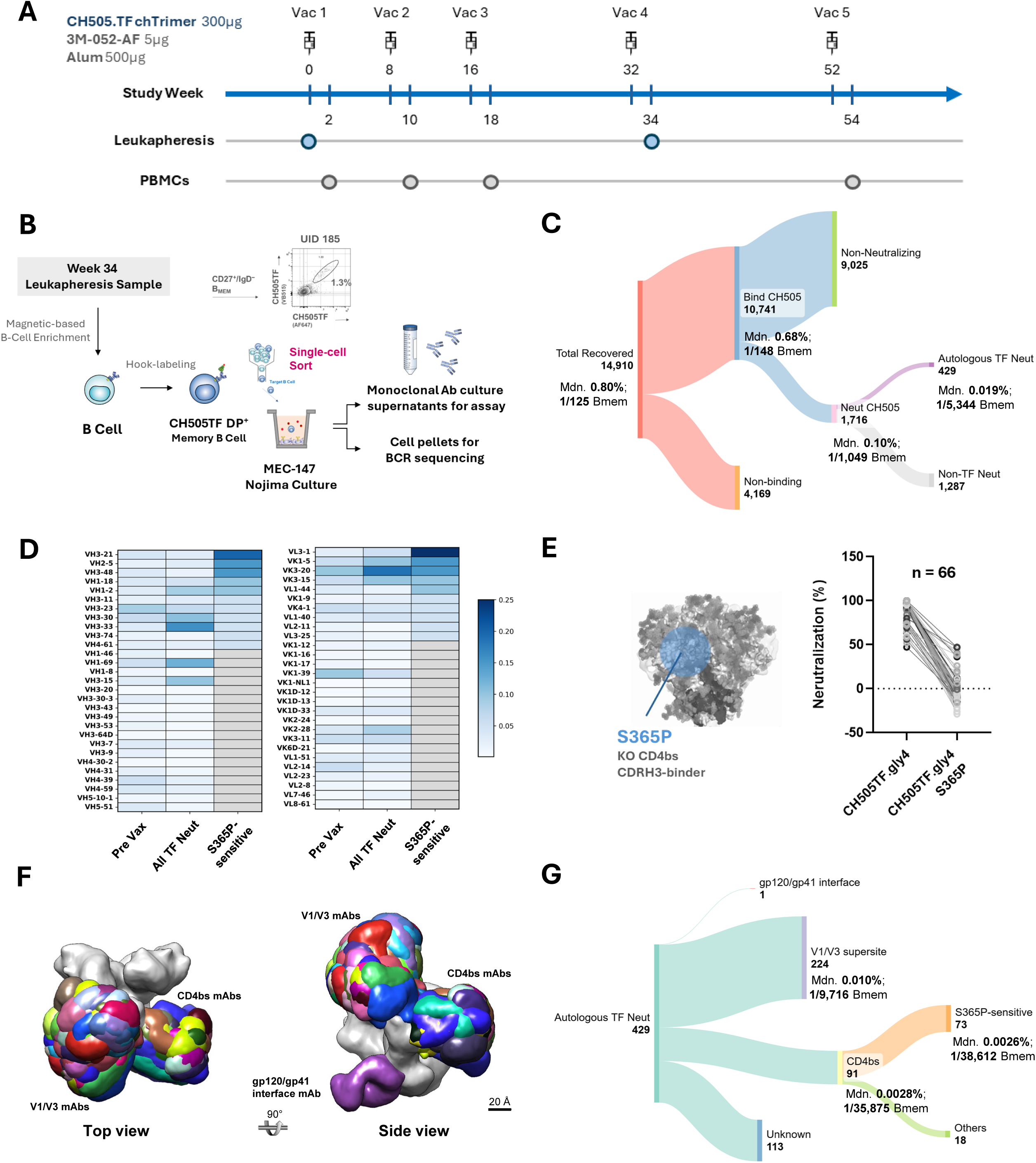
CH505 TF chTrimer immunization elicits robust serum binding responses and a diverse, polyclonal repertoire of autologous neutralizing memory B cell lineages. **(A) HVTN300 Part A Schema and Sampling Timeline.** Schematic of the HVTN 300 Part A vaccination regimen showing administration of CH505 TF SOSIP chTrimer (300 μg) formulated with 3M-052-AF (5 μg) and alum (500 μg), together with the timing of longitudinal blood draws and leukapheresis collections used for downstream B cell profiling and monoclonal culture analyses. Blue symbols indicate leukapheresis time points, and gray symbols indicate peripheral blood sampling time points. **(B) Isolation and single-cell Nojima culture of CH505 TF–reactive memory B cells from HVTN 300 leukapheresis samples.** Week 34 leukapheresis samples were subjected to magnetic bead-based B-cell enrichment, followed by fluorescent probe staining to identify CH505 TF double-positive antigen-specific memory B cells (IgD⁻/CD27^+^ Bmem). CH505 TF DP⁺ memory B cells were single-cell sorted into 96-well plates containing MEC147 feeder cells under Nojima culture conditions to support monoclonal expansion and IgG secretion. Culture supernatants were collected for downstream binding and neutralization assays, and corresponding cell pellets were retained for BCR sequencing. **(C) Stepwise classification of week 34 Nojima single-B-cell cultures recovered from CH505 TF+ memory B cells.** Sankey diagram summarizing the workflow and outcome of week 34 Nojima single-B-cell cultures derived from CH505 TF+ memory B cells in HVTN 300 Part A. Of 14,910 viable monoclonal cultures, 10,741 secreted CH505-binding antibodies, whereas binding could not be assigned for 4,169 cultures. High-throughput neutralization screening performed directly on culture supernatants identified 1,716 cultures with neutralizing activity against at least one glycan-modified CH505 TF screening pseudovirus, including 429 cultures that neutralized the autologous tier 2 CH505 TF virus. **(D) Heavy- and light-chain V-gene usage in CH505 TF–neutralizing memory B cell repertoires.** Heatmaps of clonally normalized V-gene usage (one count per lineage) among Nojima culture–derived autologous tier 2 CH505 TF-neutralizing memory B cell lineages from HVTN 300 Part A participants. Left, heavy-chain VH gene usage; right, light-chain Vκ/Vλ gene usage. Rows indicate individual V genes; columns indicate participant-level neutralizing repertoires or the pre-vaccination baseline. The Pre Vax column shows mean clonally normalized usage across all participants at baseline (10x BCR sequencing); all post-vaccination columns are derived from Nojima culture BCR sequencing. Color intensity indicates the relative frequency of each V gene within the indicated repertoire, highlighting participant-specific immunogenetic features of vaccine-elicited autologous neutralizing responses, including S365P-sensitive CD4bs CDRH3-binder lineages. **(E) Identification of putative CD4bs CDRH3-binder precursor activity by S365P sensitivity.** Schematic illustrating the S365P knockout strategy used to define CD4bs-directed, CDRH3-mediated neutralizing antibody activity within the vaccine-elicited repertoire. The S365P substitution in CH505 TF disrupts the CDRH3-binder mode of CD4 binding site recognition, allowing classification of monoclonal Nojima culture supernatants and derived mAbs as S365P-sensitive or S365P-insensitive on the basis of neutralization against matched CH505 TF viruses. Differential screening results for all 66 S365P-sensitive neutralizing cultures identified in the primary screen; an additional 7 S365P-sensitive mAbs were identified upon IC50 titration of expressed antibodies (see panel G and main text). **(F) Structural mapping reveals multiple autologous neutralizing antibody classes elicited by CH505 TF chTrimer vaccination.** Composite NSEM reconstructions of Fab–CH505 TF trimer complexes from expressed autologous tier-2 neutralizing mAbs, grouped by epitope class. Colored densities represent representative Fab footprints mapped onto the CH505 TF trimer (gray), demonstrating three major vaccine-elicited neutralizing specificities: CD4 binding site (CD4bs) antibodies, V1/V3-glycan–proximal antibodies, and a subdominant class targeting the gp120/gp41 interface region. Scale bar, 20 Å. **(G) Epitope distribution of autologous tier 2 CH505 TF-neutralizing memory B cell cultures.** Sankey diagram summarizing 429 autologous CH505 TF-neutralizing cultures identified from the total memory B cell compartment. Cultures were assigned to the CD4bs class (n = 91), V1/V3 supersite class (n = 224), gp120/gp41 interface class (n = 1), or remained unassigned (n = 113). Within the CD4bs-directed compartment, 73 cultures were S365P-sensitive and 18 were classified as other CD4bs specificities. **(C and G)** Median frequencies within total memory B cells are indicated for the major epitope classes and were calculated among participants with non-zero values.

### Unbiased memory B-cell profiling revealed a diverse vaccine-elicited neutralizing antibody repertoire

To define the vaccine-elicited B cell repertoire at single-cell resolution, we isolated antigen-specific Bmem cells from leukapheresis samples after the 4^th^ priming immunization from each HVTN300 Part A participant (**Figs. 1A and S1A**) using dual fluorophore-labeled CH505 TF SOSIP probes. Single B cells were expanded using the Nojima culture system ^28^, which enables clonal expansion and antibody secretion, yielding culture supernatants containing Ig concentrations ranging from 0.001 to 1434 µg/mL (median 9.37 µg/mL, IQR 0.30–47.04 µg/mL) (**Figs. 1B and S1B−C**). Of 14,910 viable monoclonal cultures, 10,741 (72.0%) secreted CH505 TF chTrimer-binding antibodies (**Fig. 1C**). Across participants, CH505 TF-specific Bmem cells constituted a median of 0.68% of total Bmem cells per participant (corresponding to 1 in 148 Bmem; **Fig. 1C**), indicating robust antigen-specific memory B-cell priming by CH505 TF chTrimer vaccination.

We next compared heavy- and light-chain V-gene usage among recovered vaccine-induced autologous tier 2 CH505 TF-neutralizing clonal lineages. To assess how vaccination shaped the neutralizing repertoire, we compared pre-vaccination B-cell repertoires obtained by 10x BCR sequencing with post-vaccination neutralizing lineages recovered from antigen-specific cultured B cells. These analyses revealed participant-specific patterns of VH and Vκ/Vλ gene usage among vaccine-elicited CH505 TF-neutralizing memory B cell lineages, including lineages with features consistent with S365P-sensitive CD4bs CDRH3-dominant responses (**Figs. 1D and S1D−E**). Together, these analyses demonstrated that CH505 TF chTrimer vaccination elicited a genetically diverse repertoire of neutralizing memory B-cell lineages, including lineages with features consistent with CDRH3-dominant CD4bs responses.

To benchmark the Nojima platform against conventional vaccine antigen-specific single-cell sorting and PCR, we directly compared the two approaches using paired samples from four participants (UID 123, 185, 469, 475; **Methods**). Nojima cultures recovered every S365P-sensitive neutralizing lineage detected by antigen-specific single-cell PCR analysis, while also identifying seven additional S365P-sensitive neutralizing B cell lineages (**Figs. S1F−H; Methods**).

Having established the sensitivity of the Nojima platform, we next performed TZM-bl pseudovirus neutralization assays directly on all 10,741 Nojima culture supernatants, enabling functional definition of the full vaccine-induced neutralizing antibody repertoire. This assay-first workflow allowed us to determine the frequency of neutralizing Bmem cells within the total Bmem cell compartment. Of 10,741 confirmed CH505 TF-reactive cultures, 1,716 neutralized at least one glycan-modified CH505 TF pseudovirus (CH505TF.gly4, CH505TF.gly4.S365P.2, CH505TF.G458Y.N279K/GnT1⁻, or CH505TF.G458Y.N279K.N280D/GnT1⁻) and 429 neutralized the autologous tier 2 CH505 TF pseudovirus, corresponding to a total median trial frequency of 1 neutralizing antibody-producing cell in 5,344 total Bmem cells across the trial (**Fig. 1C**). Among 429 cultures that neutralized the autologous tier 2 CH505 TF pseudovirus, 66 exhibited S365P-sensitive neutralization, a signature of CD4bs-directed, CDRH3-dominant bnAb precursor activity (**Fig. 1E**) ^35^. S365P-sensitive cultures were detected in 8 of 11 participants (73%) (**Data S1**).

### Polyclonal CDRH3-dominant CD4bs neutralizing response was associated with preferential light-chain usage

Genetic characterization of 429 autologous tier-2 neutralizing cultures revealed diverse VH/VL gene usage, variable levels of somatic hypermutation and a broad range of CDR3 lengths across participants **(Data S1)**. Whereas B cell lineage–based design strategies aim to selectively engage defined bnAb precursor classes ^36,37^, our results demonstrated that, within the S365P-sensitive subset, the CH505 chTrimer elicited a polyclonal antibody response rather than a single bnAb lineage. The prototype CDRH3-binding bnAb CH103 utilized heavy chain gene VH4-59 paired with the light-chain gene Vλ3-1. While none of the vaccine-elicited S365P-sensitive antibodies used VH4-59, 47% of the 73 S365P-sensitive cultures utilized the Vλ3-1 gene, accounting for 5 of the 20 lineages (**Data S1**). These data suggested that VH4-59 usage is not required for vaccine-elicited CDRH3-dominant CD4bs neutralization and that light-chain identity may represent a key determinant of this mode of Env recognition (**Fig. 1D**).

### CH505 TF chTrimer elicited neutralizing antibody responses targeting multiple Env epitopes

We next expressed 150 recombinant monoclonal antibodies (mAbs) selected to capture the diversity of the 429 autologous CH505 TF tier 2-neutralizing cultures. This analysis identified seven additional mAbs with S365P-sensitive, CDRH3-dominant signatures (**Fig. 1C and Data S1–S2**). Thus, a total of 73 candidate CDRH3-dominant CD4bs precursor mAbs were isolated from 8 of 11 participants, with a median frequency of 1 in 38,612 total memory B cells (0.0026%) among these participants (**Figs. 1F–G and Table S1**). We obtained negative-stain electron microscopy (NSEM) 3D reconstructions of 131 of the 150 expressed mAbs, representing 102 distinct antibody clones (**Figs. 1F, S2, Table S2 and Data S1**). Lineage-level epitope assignments for these 102 autologous CH505 TF tier 2-neutralizing B-cell clones demonstrated that the vaccine-elicited neutralizing response was broadly distributed across multiple Env epitopes rather than restricted to a single antibody specificity. The dominant specificity comprised N332-independent autologous neutralizing antibodies targeting the V1/V3-glycan site at the base of V3, which occurred at a median frequency of 1 in 9,716 total memory B cells among participants in whom this response was detected (**Table S1**). This class was followed by CDRH3-dominant CD4bs neutralizing antibodies, detected at a median frequency of 1 in 38,612 total memory B cells among the 8 responding participants, and by a single clonal lineage, DH1708, directed against the gp120–gp41 interface (**Figs. 1F–G and S2 and Table S1**).

Together, these data demonstrated that CH505 TF chTrimer vaccination elicited a clonally diverse repertoire of autologous tier 2 neutralizing antibodies in humans that targeted multiple Env epitopes, including the V1/V3-glycan region, the CD4 binding site, and the gp120/gp41 interface.

### CD4bs-targeting mAbs elicited in HVTN 300 adopted multiple structural solutions for CD4bs recognition

The CD4bs lineages DH1530 and DH1541, together with mAbs DH1783 and DH1777.1, neutralized the autologous tier 2 CH505 TF virus, bound CH505 TF chTrimers with high affinity, and competed with soluble CD4 and the prototype CH103 and CH106 Fabs for Env trimer binding (**Figs. 2A−B; Data S1−2**). Both the DH1530 and DH1541 lineages, as well as the DH1783 singleton, utilized Vλ3-1 and exhibited reduced neutralization of the S365P mutant, consistent with a CDRH3-dominant CD4bs recognition **(Data S1−S2)**. Neither DH1530 nor DH1541 or DH1783 neutralized heterologous HIV-1 strains, placing these lineages at an intermediate stage of maturation analogous to the CH103 lineage member IA4 **(Fig. 2A and Data S2)** ^18^.

**Figure 2.**
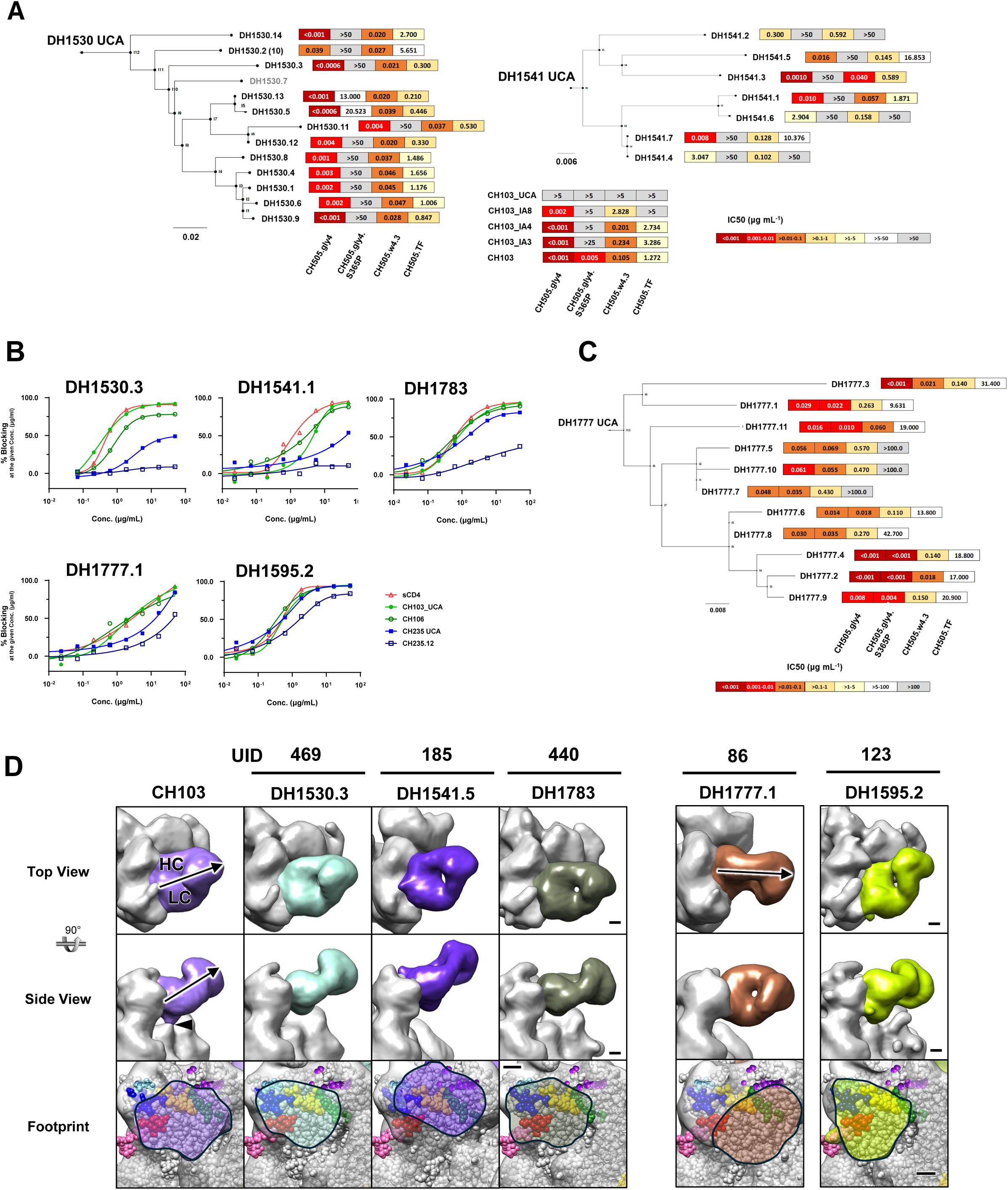
CD4bs CDRH3-binder mAbs from HVTN 300 show functional and structural convergence with prototype CH103-class antibodies, alongside alternative CD4bs solutions. **(A) Neutralization profiles of DH1530 and DH1541 CD4bs lineages across autologous CH505 TF variants.** Phylogenetic trees depict clonal relationships within the DH1530 (left) and DH1541 (right) lineages, rooted on the inferred unmutated common ancestor (UCA). For each monoclonal antibody, heatmaps report TZM-bl neutralization potency (IC₅₀, μg mL⁻1; color-coded as indicated) against the autologous CH505 TF virus panel, including CH505.gly4, CH505.gly4.S365P, CH505.w4.3, and CH505 TF. Reference CH103-lineage antibodies (CH103 UCA and maturation intermediates IA8, IA4, IA3, and mature CH103) are shown for comparison with the same virus panel. Scale bars indicate substitutions per site for each lineage tree. **(B) CD4bs blocking (competition) assays for HVTN300 mAbs.** Blocking assays measuring binding of soluble CD4 (sCD4) or CD4bs bnAbs (CH103 UCA, CH106, CH235 UCA, and CH235.12) to their corresponding CH505 Env variants in the presence of increasing concentrations of competing DH1530.3, DH1541.1, DH1783, DH1777.1 and DH1595.2 as indicated. Curves show dose-dependent inhibition relative to no-competitor controls. **(C) Neutralization profiles of DH1777 CD4bs lineage across autologous CH505 TF variants.** Phylogenetic trees depict clonal relationships within the DH1777 lineage, rooted on the inferred UCA. For each monoclonal antibody, heatmaps report TZM-bl neutralization potency (IC₅₀, μg mL⁻1; color-coded as indicated) against the autologous CH505 TF virus panel, including CH505.gly4, CH505.gly4.S365P, CH505.w4.3, and CH505 TF. Scale bars indicate substitutions per site for each lineage tree. **(D) NSEM reconstructions of DH1530.3, DH1541.5, DH1783, DH1777 and DH1595.2 mAb–CH505 TF complexes.** For each complex, the top two rows show two orthogonal views of the Fab-bound Env protomer (trimer three-fold axis tilted), and the bottom row shows the corresponding Fab footprint. Footprints are rendered with the NSEM density as a transparent surface and a fitted SOSIP model (PDB 5FYK) displayed as space-filling spheres. Env is shown in gray; Fabs are colored; structural landmarks are highlighted as indicated (CD4-binding loop, gold; V5 loop, blue; D loop, red), and selected glycans are shown (N276, pink; N463, light blue; N197, green; N187, purple). Scale bars, 20 Å (NSEM maps) and 10 Å (footprints).

In contrast, DH1777.1 neutralization was not sensitive to the S365P mutation, and the antibody exhibited modest heterologous breadth, neutralizing 6.7% of a 134 isolate global HIV-1 panel (**Fig. 2C and Table S3**). Despite sharing the VH4-59 heavy-chain gene with CH103, DH1777.1 paired with Vκ3-20 rather than Vλ3-1, possessed a longer CDRH3 of 23 amino acids compared with 15 amino acids in CH103, and carried somatic mutation frequencies of 3.86% in VH and 1.5% in VL. We identified 10 additional members of the DH1777 clonal lineage (DH1777.2–DH1777.11) by antigen-specific sorting of CH505 TF-reactive B cells followed by high-throughput paired heavy- and light-chain BCR sequencing using the 10x Genomics Chromium platform (**Fig. 2C**). Binding and neutralization analyses showed that these newly identified lineage members retained CH505 TF SOSIP trimer recognition and exhibited functional profiles similar to that of DH1777.1, including comparable autologous neutralization activity and limited heterologous breadth (**Fig. 2C and Table S3**).

Indeed, NSEM analysis revealed a CH103-like angle of approach for the S365P-sensitive DH1530 and DH1541 lineages and the DH1783 singleton (**Fig. 2D**). By contrast, the S365P-insensitive neutralizing antibody DH1777.1 adopted a more horizontal approach reminiscent of the CDRH3-dominant antibody b12 ^38^. A third distinct mode was observed in the DH1595 lineage antibody, which differed primarily in its rotation about the long axis of the Fab (**Fig. 2D**). These distinct modes of CD4bs engagement demonstrate that CH505 TF chTrimer vaccination elicited a polyclonal neutralizing antibody response, in which multiple CDRH3-dominant lineages converged on the CD4bs through alternative structural solutions.

### Cryo-EM structures reveal molecular pathways toward CDRH3-dominant CD4bs recognition

To define the molecular basis of Env recognition by CD4bs-targeting mAbs, we performed single-particle cryo-EM analyses of representative antibodies from the DH1530, DH1541, DH1786.1, DH1595, and DH1777 lineages in complex with the CH505 TF chTrimer (**Figs. 3A–D, S3–S4**). In all structures except the DH1595.2-bound complex, the Env adopted an open conformation characterized by disruption of the V1/V2 cap at the Env trimer apex. In the DH1595.2 complex, a quaternary interaction between the Fab constant domain and a neighboring Env protomer likely stabilized the Env trimer in a closed conformation (**Figs. 3A and S4F**). For the DH1530.3-, DH1595.2-, and DH1786.1-bound complexes, map resolutions enabled unambiguous backbone model building and residue-level visualization of Env–antibody interaction interfaces (**Figs. S3 and S4F−J**). In contrast, the DH1777.1-bound complex yielded an 8.39 Å reconstruction that permitted rigid-body fitting but not detailed interface modeling. To obtain higher-resolution structural information, we determined the structure of a ternary complex comprising CH505 TF SOSIP, DH1777.3 and the fusion peptide-directed antibody VRC34.01^30^. Although Env remained in an open, V1/V2 cap-disrupted conformation, VRC34.01 reduced conformational heterogeneity and improved the reconstruction to ∼6 Å resolution (C1 symmetry), enabling more detailed characterization of the DH1777 Fab/Env-interface (**Figs. S4E and S4J**).

The binding footprints of DH1595.2 and DH1786.1 were displaced from the conserved CD4 binding loop and predominantly focused on the variable Loop D and V5 regions (**Figs. 3A–B, E–F and S3**), suggesting a less favorable trajectory toward broad neutralization ^39^. In contrast, the bnAb precursor candidates DH1530.3 and DH1777.3 made their primary contacts with the conserved CD4 binding loop (**Figs. 3C–D, G–H and S3**).

Among the antibodies studied with cryo-EM, DH1530.3 exhibited the most similar binding geometry to the prototype CH103 antibody, engaging the canonical CD4bs with a comparable angle of approach and orientation (**Figs. 3C, 3G**, and **S5A**). The gp120-DH1530.3 interface revealed interactions of the DH1530.3 CDRH3 with the CD4-binding loop and loop D, recapitulating hallmark contacts of CH103 (**Figs. 3C and S5B-C**). In mature CH103, the CDRH3 plays a central role in contacting the CD4-binding loop and surrounding CD4bs elements, while the heavy chain framework region 3 (FR3) makes quaternary contacts with the electropositive surface of the neighboring protomer by utilizing negatively charged residues E75 and D76 acquired early during CH103 lineage maturation ^40^. Although DH1530.3 resembled CH103 in its primary CD4bs binding mode, its lack of FR3-mediated contacts with the neighboring gp120 protomer may prevent its ability to engage a closed Env trimer. To test whether the acquisition of heavy chain FR3 negative charges affected DH1530.3 binding to Env, we added two mutations, G75E and N76D (DH1530.3_ED), or three mutations, A74S, G75E, and N76D (DH1530.3_SED), to DH1530.3. We tested the binding of DH1530.3 and the two mutants to closed CH505 TF SOSIP Env by SPR, where the Env was constrained in the closed conformation by the closed-conformation-selective antibody PGT145 (**Figs. S5D−E**). Although DH1530.3 bound PGT145-captured Env, binding by the DH1530.3_ED and DH1530.3_SED variants was substantially enhanced, indicating that, as observed for CH103, acquisition of acidic FR3 residues during affinity maturation facilitates engagement of native closed Env trimers and may promote the development of greater neutralization potency and breadth.

**Figure 3.**
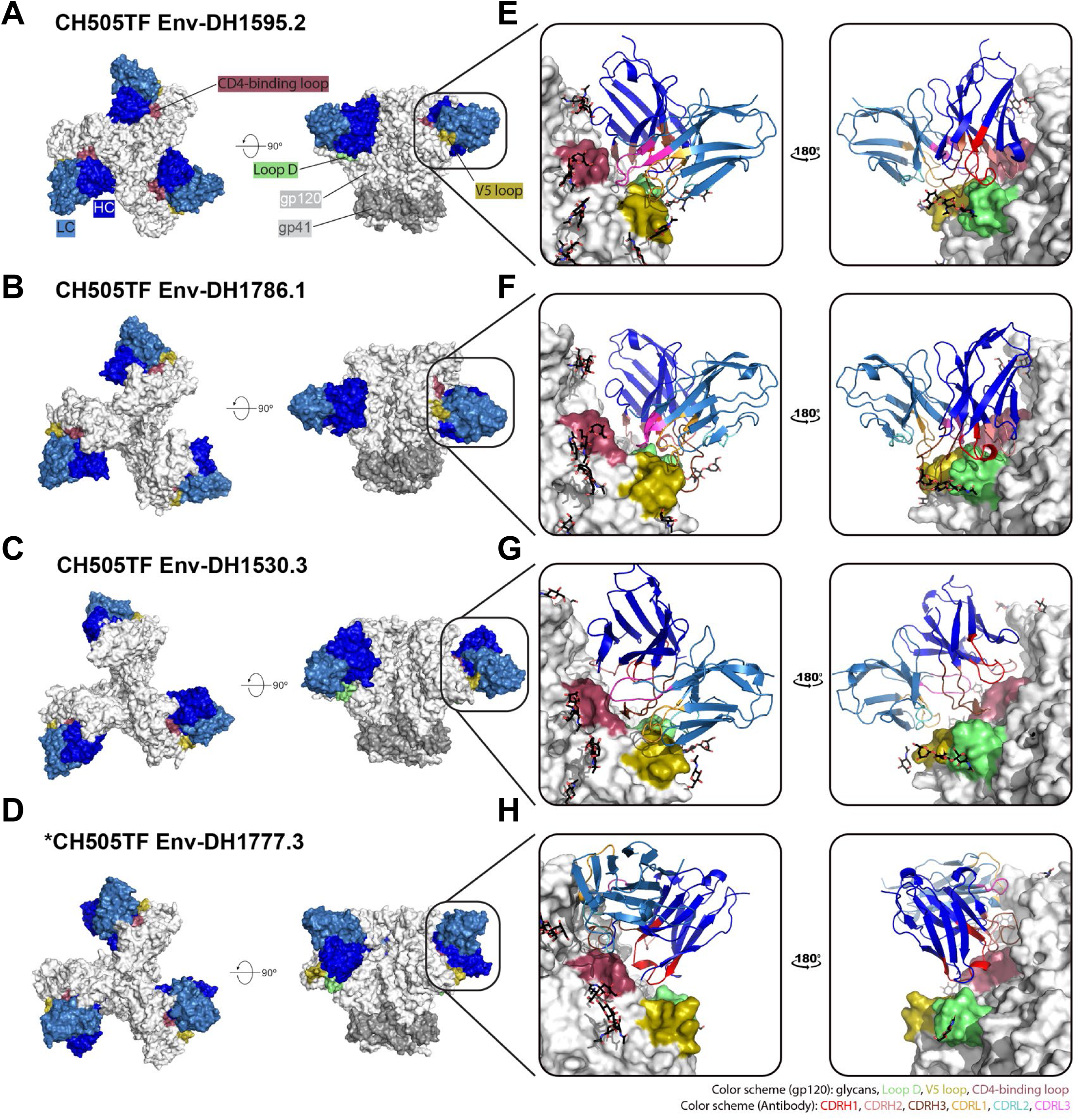
Structural diversity of vaccine-elicited CD4bs antibodies isolated in the HVTN 300 clinical trial. (A, B, C, and D) Cryo-EM structures of HVTN300 CD4bs targeting antibodies bound to CH505 TF SOSIP. Refined cryo-EM maps are shown in two orientations. gp41 is colored gray, and gp120 is colored white, with its CD4-binding loop colored raspberry, loop D colored mint, and the V5 loop colored olive. **(E, F, G, and H) Zoomed in view of the CD4bs-antibody interface**. CD4bs-targeting antibodies are shown in cartoon representation, and Env is shown in surface view. Similar color scheme as Panel A-D. Glycans, CDRH1, CDRH2, CDRH3, CDRL1, CDRL2, and CDRL3 loops are colored black, red, salmon, chocolate, light orange, aquamarine, and light magenta, respectively. *VRC34.01, which is also bound to CH505TF Env alongside DH1777.3, is not shown in the figure.

Unlike DH1530.3, DH1777-lineage mAbs did not bind PGT145-captured Env constrained in the closed conformation, indicating that partial Env opening was required for DH1777 engagement (**Figs. S5F−G**). This observation is consistent with the DH1777 footprint, which resembles that of the CDRH3-dominant bnAb b12, an antibody that preferentially recognizes an open, occluded Env conformation ^16,41^. DH1777.3 binding was centered on the CD4-binding loop, while being shifted away from Loop D and V5 and toward the V2 apex region. The DH1777 CDRH3 is longer than that of b12 and extends beneath the gp120 ß20-ß21 region and the V2 apex (**Fig. S5H**).

Together, these structural analyses reveal multiple vaccine-elicited pathways to CD4bs recognition. Whereas DH1595.2 and DH1786.1 preferentially targeted variable Env elements and may be less likely to mature toward breadth, DH1530 and DH1777 recapitulated key structural features of the CH103- and b12-type bnAb pathways, respectively. These findings demonstrate that CH505 TF chTrimer vaccination can engage diverse CDRH3-dominant lineages with distinct molecular trajectories toward HIV-1 neutralization.

### Predominant N332-independent V3-directed responses targeted the conserved GDIR motif

Although HVTN 300 was designed to elicit CDRH3-dominant CD4bs bnAb precursor lineages, Nojima culture analysis revealed neutralizing antibodies targeting two additional Env epitopes: a dominant N332-independent V1/V3-directed response and a gp120/gp41 interface-directed response (**Figs 1F−G**, **4, and S6−S7**).

**Figure 4.**
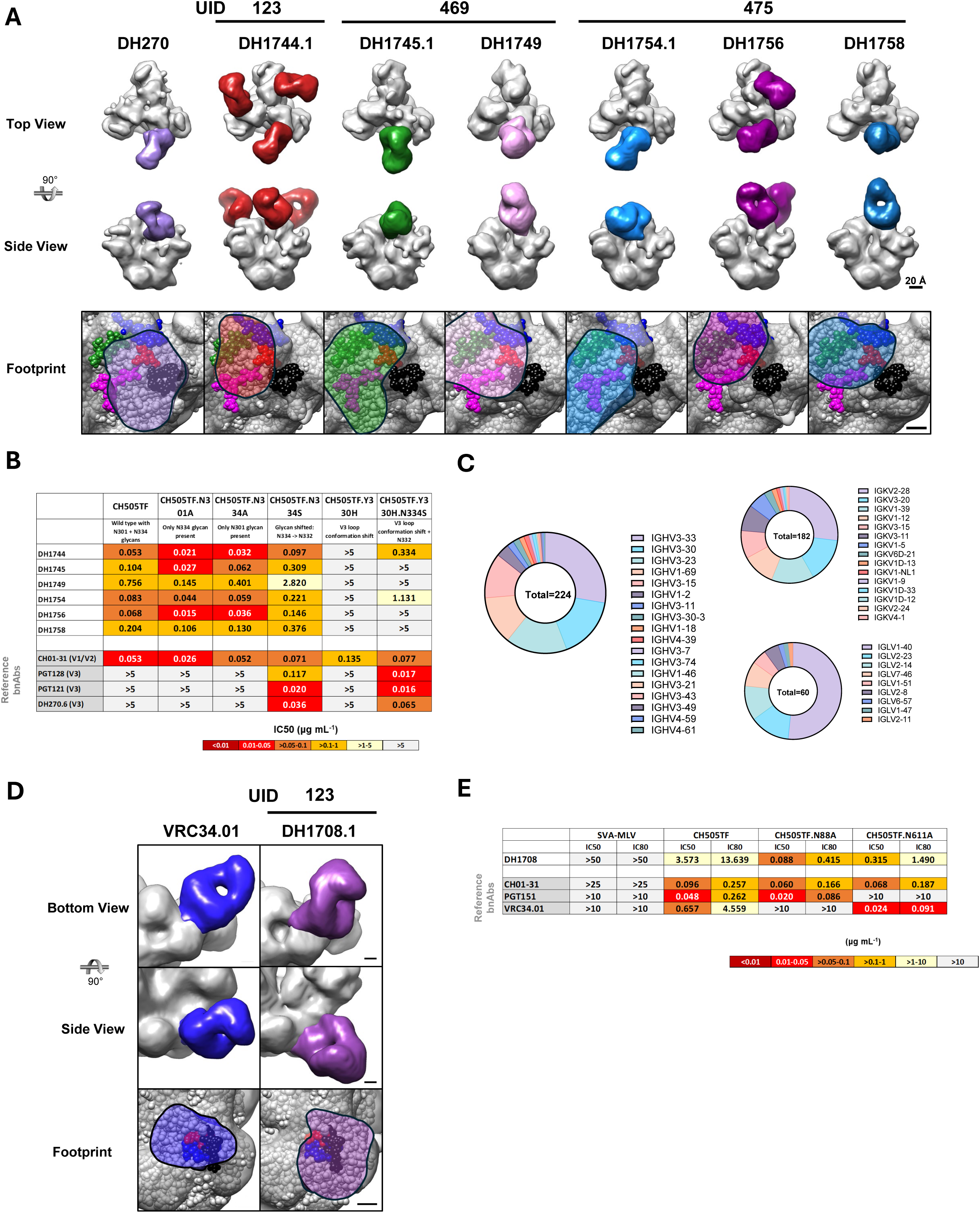
Neutralizing B cell lineages targeting the V3-glycan supersite and the fusion-peptide proximal region elicited by CH505 TF chTrimer immunization. **(A) NSEM reconstructions and epitope footprints of V1/V3-directed mAb–Env trimer complexes.** NSEM reconstructions of a reference V3-glycan bnAb DH270 mAb–Env complex are shown alongside representative HVTN300 V1/V3-directed autologous neutralizing mAbs (DH1744.1 from participant UID123; DH1745.1 and DH1749 from UID469; and DH1754.1, DH1756, and DH1758 from UID475) in complex with Env. For each complex, two orthogonal views are shown with Env rendered in gray and antibody densities colored. The bottom row shows zoomed-in footprints, with NSEM density displayed as a transparent surface and a fitted SOSIP model (PDB 5FYK) shown as space-filling spheres. Structural landmarks are colored as indicated: V1 loop (blue) and the V3-base GDIR motif (red); glycans N156 (green), N301 (magenta), and N332 (black). The fitted model (PDB 5FYK; JR-FL) contains an N332 glycan as displayed; in contrast, the NSEM maps were generated using CH505 Env, which lacks N332 and instead carries an N334 glycan (not shown; positioned ∼7 Å to the right in this view). Scale bars, 20 Å (NSEM maps) and 10 Å (footprints). **(B) Neutralization of representative V1/V3-directed mAbs against CH505 TF Env variants that perturb the V3 loop and surrounding glycan environment.** Heatmap summarizes TZM-bl neutralization potency (IC₅₀, μg mL⁻1; color-coded as indicated) of representative V1/V3-directed mAbs against CH505 TF wild-type (WT) and a panel of autologous Env variants engineered to alter V3 conformation and/or local glycan shielding. Variants include (i) sequon changes at N301 to remove or restore the N301 glycan, (ii) modulation of the N332/N334 glycan-site context (CH505 TF lacks N332 and instead carries an N334 glycan), and (iii) Y330H to perturb V3-loop conformation, as well as the compensatory Y330H+N334S combination, which introduces the N332 while removing N334 glycan. Reference antibodies (CH01-31, PGT128, PGT121, and DH270.6) are shown for comparison. **(C) Germline V-gene usage of V1/V3-directed autologous neutralizing antibodies.** Donut plots summarize immunoglobulin gene usage among V1/V3-directed autologous neutralizing antibodies identified from HVTN300. Distributions are shown for heavy-chain (VH; left) and light-chain V genes (kappa and lambda; top and bottom right, respectively). Each segment represents the fraction of sequences assigned to the indicated V gene. **(D) NSEM reconstructions and epitope footprints of gp120/gp41 interface-directed DH1708.1 mAb–Env complexes.** NSEM reconstructions of the gp120/gp41 interface-directed mAb DH1708.1 from UID123 in complex with CH505 TF SOSIP are shown alongside the prototype fusion-peptide antibody VRC34.01 (left) for reference. For each complex, two orthogonal views are shown with Env rendered in gray and antibody densities colored. The bottom row shows zoomed-in footprints, with NSEM density displayed as a transparent surface and a fitted SOSIP model (PDB 5FYK) shown as space-filling spheres. Structural landmarks and selected glycans are colored as indicated: Red FP (residues 521-528), blue FPPR (residues 529-540), black N88 glycan. Scale bars, 20 Å (NSEM maps) and 10 Å (footprints). **(E) Neutralization of DH1708.1 against CH505 TF and glycan-site mutants.** Heatmap summarizes TZM-bl neutralization potency (IC₅₀ and IC₈₀, μg mL⁻1; color-coded as indicated) of DH1708.1 against CH505 TF and matched glycan-site variants (CH505TF.N88A and CH505TF.N611A), with SVA-MLV included as a negative-control virus. Reference mAbs (CH01-31, PGT151, and VRC34.01) are shown for comparison.

N332-independent V3-glycan-targeted Abs have been demonstrated to evolve to mediate heterologous neutralization ^31–33^. Across the 11 HVTN300 participants, we identified 224 V1/V3-targeted monoclonal cultures comprising 79 distinct clonal lineages and 52.2% of all recovered 429 tier 2 autologous neutralizing B cells (**Data S1**).

By NSEM, the vaccine-induced V1/V3-directed mAbs displayed varying degrees of overlap with the V1 loop, and the conserved V3 base GDIR motif (residues 324-327) that is also engaged by canonical V3-glycan bnAbs (**Fig. 4A**). CH505 TF lacks the N332 glycan but is glycosylated at N334. Removal of the N301 and N334 glycans enhanced autologous virus neutralization by the V1/V3-directed mAbs (**Fig. 4B**) likely by increasing accessibility of the underlying protein epitope and/or permitting more favorable interactions with neighboring glycans ^42^. The Env Y330H substitution abrogated neutralization by the V1/V3-directed mAbs, whereas the Y330H + N334S combination, which removes the N334 glycan and adds an N-glycosylation sequon at N332, partially rescued activity ^24^.

Genetic analysis of the 224 V3-directed B cells revealed biased germline usage, dominated by the VH3 family (79.5%) and Vλ1-40 (51.7%) (**Fig. 4C; Data S1**). Enrichment of VH3-33 and VH3-30, and an average CDRH3 length of 17 amino acids, highlighted a requirement for specific heavy chains to target the conserved GDIR motif. The strong enrichment of Vλ1-40 suggests that this light chain confers a selective advantage for recognition of the N332-independent V1/V3 epitope, enabling maintenance of neutralizing activity despite glycan heterogeneity at positions N301 and N334/N332.

To define the molecular basis of N332-independent V1/V3 recognition, we determined cryo-EM structures of the representative antibodies DH1744.1 and DH1745.1(**Figs. 5A−E, S6A−B**). Cryo-EM structures of CH505 TF chTrimer in complex with DH1744.1 Fab or DH1745.1 Fab were solved at 4.4Å resolution (**Fig. S6A−D**). Both Fabs targeted the V3-glycan supersite but with opposite relative orientations of the heavy and light chains (**Figs. 5A−B and S6E**).

**Figure 5.**
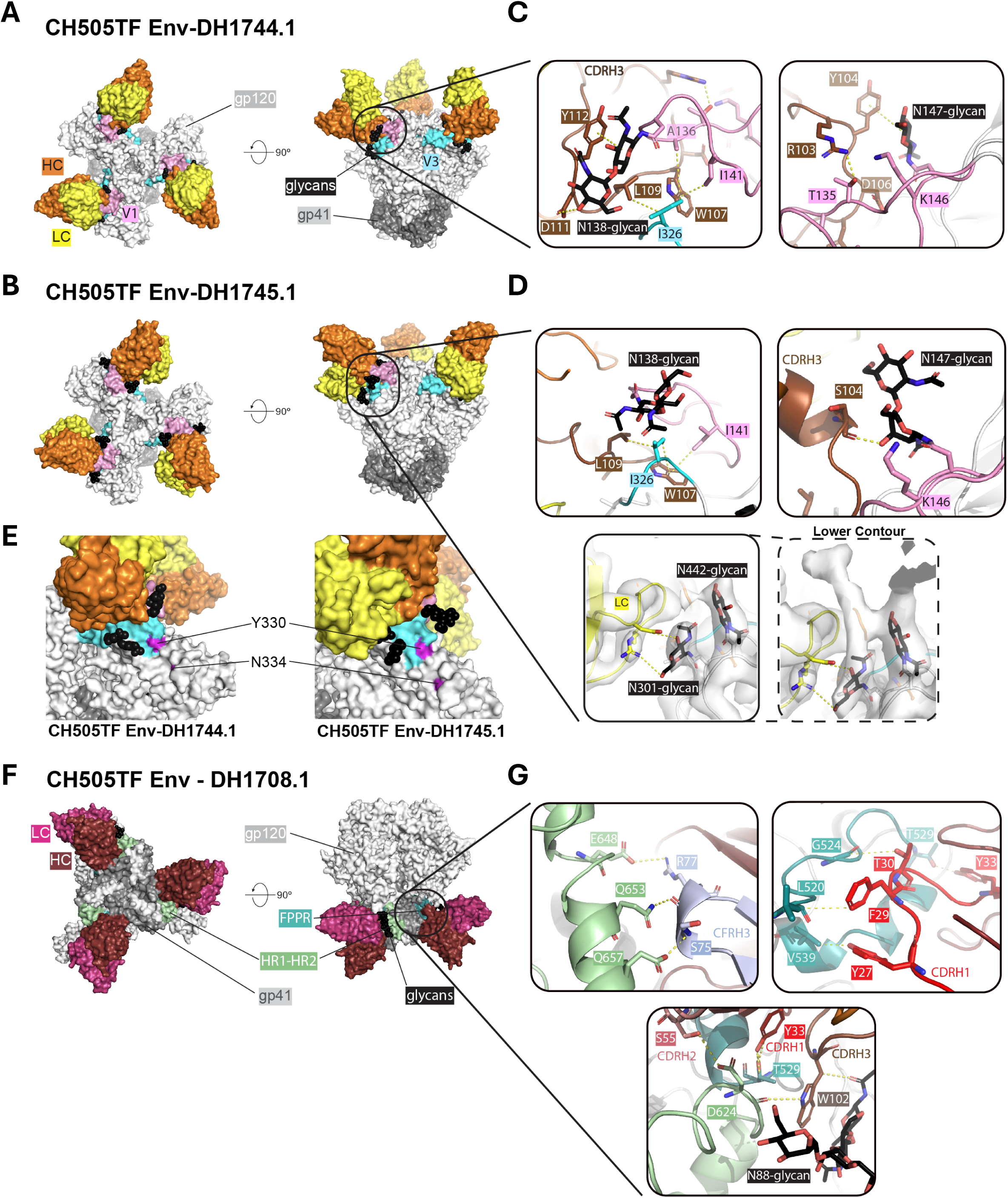
Structural definition of antibodies elicited in the HVTN 300 clinical trial that target epitopes outside the CD4 binding site. (A) Cryo-EM structure of CH505 TF Env bound to DH1744.1 Fab. Overall view of the DH1744.1-CH505 TF Env complex shown in surface representation. The heavy and light chains are colored orange and yellow, respectively. The gp120 and gp41 subunits are colored light gray and dark gray, respectively. The V1 loop is colored pink, V3 GDIR motif is colored cyan. **(B) Cryo-EM structure of CH505 TF Env bound to DH1745.1 Fab.** Overall view of the DH1745.1-CH505 TF complex shown in surface representation. DH1745.1 targets the same general region as DH1744.1 but adopts a distinct relative orientation of the heavy and light chains. A similar color scheme is used as in panel A. **(C) Close-up view of the DH1744.1 interface.** V1-loop glycans at N138 and N147 surround the Fab and contact CDRH3 residues Y104, D111, and Y112 through C-H-π interactions and hydrogen bonds. W107 from the long CDRH3 inserts beneath the V1 loop and forms hydrophobic interactions with V1 residue I141 and V3 residue I326; together with L109, these residues form a hydrophobic core at the interface. **(D) Close-up view of the DH1745.1 interface.** Most contacts are similar to those observed for DH1744.1, except that the N138 glycan is displaced away from the Fab. The DH1745.1 light chain is in close contact with the V3 N301 glycan. **(E) Location of residues Y330 and N334 on the DH1744.1- and DH1745.1-bound structures.** Residues Y330 and N334 are shown in magenta on the (left) DH1744.1-bound and (right) DH1745.1-bound structures. **(F) Cryo-EM structure of CH505 TF Env bound to DH1708.1 Fab.** Overall view of the DH1708.1-CH505 TF complex shown in surface representation. DH1708.1 engages the gp41 fusion peptide proximal region (FPPR) using its heavy chain, with no light-chain contacts observed. The heavy and light chains are colored ruby and warm pink, respectively. The gp120 and gp41 subunits are colored light and dark gray, respectively. The FPPR is colored deep teal, and the region between HR1 and HR2, spanning residues 617-629, is colored pale green. **(G) Close-up view of the DH1708.1 interface.** S75 and R77 form electrostatic interactions with Q653 and E648 at the C-terminal end of the HR2 regions (HRC2) of an adjacent gp41 protomer. CDRH1 forms extensive hydrophobic and hydrogen-bonding contacts with FPPR residues, with an additional interaction between S55 in CDRH2 and D624. A single glycan contact is observed through W102 in CDRH3 and the first N-acetylglucosamine of the N88 glycan.

In DH1744.1, V1 loop glycans at N138 and N147 surrounded the Fab, forming C-H-π interactions and hydrogen bonds with CDRH3 residues Y104, D111, and Y112 **(Fig. 5C).** CDRH3 residue W107 inserted under the V1 loop, establishing hydrophobic contacts with V1 residue I141 and residue I326 in the conserved V3 GDIR/K motif (I326 is conserved in 97.3% of 8,735 global HIV-1 Envs in the Los Alamos HIV Database); together with CDRH3 residue L109, these residues formed a hydrophobic core anchoring the antibody to the V3 base. In DH1745.1, most of these interactions were conserved; however, the N138 glycan was displaced away from the Fab due to differences in CDRH3 geometry, with the light chain instead forming close contact with the N301 glycan on the V3 loop, compensating for the loss of N138 engagement (**Figs. 5D and S6E**). These differences demonstrate that diverse structural solutions can accommodate variable glycan interactions while converging on recognition of the conserved GDIR peptide backbone ^24^.

DH1744.1 and DH1745.1 shared structural features with N332-independent V3-glycan antibodies elicited in nonhuman primates by both CH505 TF chTrimer (DH1024/DH1025.1) ^24^ and the WIN332 immunogen (Ab1983/Ab1999) ^31^, suggesting convergent targeting of this Env vulnerability across immunogens and host species. Like DH1025.1, DH1744.1 and DH1745.1 contacted the V1/V3 interface and engaged the GDIR motif while remaining distant from the N332/N334 glycan site (**Fig. 5E**); similarly, their hydrophobic core formation resembled that of Ab1999 ^31^. However, a notable distinction within this antibody class is the degree of V1 loop engagement. Whereas DH1025.1, Ab1983, and Ab1999 formed more extensive contacts with the hypervariable V1 loop, DH1744.1 and DH1745.1 maintained a greater physical distance from V1, with correspondingly reduced V1-dependent interactions (**Fig. S6E**). This reduced V1 dependence may confer a recognition profile less susceptible to viral escape, since the V1 loop varies dramatically in sequence and length across global HIV-1 Envs ^43^. Moreover, residual V1 contacts suggest that further maturation away from V1 engagement may be required to fully achieve cross-clade Env recognition. Although residue Y330 was outside the Env binding interfaces of DH1744.1 and DH1745.1 (**Fig. 5E**), neutralization was completely abrogated by the Y330H mutation (**Fig. 4B**). The N334S substitution, which restored the N332 glycan, partially rescued activity in DH1745.1 but not in DH1744.1.

Together, these data demonstrate that DH1744.1 and DH1745.1 recognize the V3 base through a conserved GDIR-centered, N332-independent mechanism. Their reduced dependence on the highly variable V1 loop, together with convergent structural features shared with previously described N332-independent V3-directed antibodies, suggests a potential pathway toward broader Env recognition and establishes the conserved GDIR peptide backbone as a target of vaccine-elicited neutralizing antibody responses in humans.

### A vaccine-elicited gp120/gp41 interface lineage targets the fusion peptide-proximal region

Beyond the dominant V3-directed response, our unbiased functional screen identified a neutralizing B cell lineage, designated DH1708, that targeted the gp120/gp41 interface. Immunogenetic analysis showed that the DH1708 lineage utilizes a distinct VH1-2 heavy chain paired with a Vκ1D-13 light chain and encoded a 12-amino-acid CDRH3.

NSEM epitope mapping localized DH1708.1 to the gp120/gp41 junction region, with density consistent with proximity to the trimer base (**Fig. 4D**). Functionally, DH1708.1 neutralized CH505 TF and early autologous variants but was insensitive to canonical fusion-peptide mutations, supporting a mode of recognition distinct from prototype fusion-peptide antibodies such as VRC34.01 (**Fig. 4E**) ^30,44,45^. Although removal of the N88 or N611 glycans modulated DH1708.1 neutralization, the antibody retained activity against CH505 variants lacking these sites, unlike prototype FP/gp41-interface antibodies VRC34.01 and PGT151 (**Fig. 4E**).

Cryo-EM analysis of DH1708.1(**Figs. 5F−I and S7**) identified a gp120/gp41 interface epitope centered on the fusion peptide-proximal region (FPPR) helix (residues 529-532), immediately C-terminal to the fusion peptide (residues 512-520), consistent with the NSEM and neutralization data (**Figs. 4D−E**). DH1708.1 approached the Env trimer base at a shallow angle distinct from classical FP-directed bnAbs such as VRC34.01, engaging the FPPR of one gp41 protomer exclusively through heavy-chain interactions, with all three heavy-chain CDRs contributing to binding and the light chain making no detectable contacts (**Fig. 5F**) ^30^. At the FPPR interface, CDRH1 formed extensive hydrophobic and hydrogen-bonding interactions with FPPR residues, described here as Ab-FPPR pairs, including Y27-L520, F29-L520, T30-A525, and Y33-T529, while S55 in CDRH2 interacted with D624 (**Fig. 5G**). Beyond FPPR contacts, framework region 3 of the heavy chain (CFRH3; residues 67-98) interacted with residues 648-653 in the heptad repeat region C2 (HRC2) of an adjacent gp41 protomer (**Fig. 5G**), with Ab residues S75 and R77 forming electrostatic interactions with gp41 residues Q653 and E648, respectively (**Fig. 5H**). In contrast to VRC34.01, which sits over the N88 glycan, DH1708.1 made only a single glycan contact via W102 in the CDRH3 with the first NAG moiety of N88 (**Fig. 5G**).

The DH1708.1 epitope overlapped the gp120/gp41 junction and encompassed the FPPR, the HR1-HR2 intervening region (residues 617–629), and HR2 residues of an adjacent gp41 protomer, similar to the previously described antibody 3BC315 (PDB: 9ogl)^46^, but with a key difference in approach angle (**Figs. 5F−G**). Whereas 3BC315 approached gp41 nearly parallel to the FPPR, enabling contributions from both heavy and light chain, DH1708.1 approached at a shallower angle that restricted Env engagement to the heavy chain, resulting in a more compact interaction footprint. This mode of base-proximal gp41 recognition highlighted the structural plasticity of vaccine-elicited neutralizing responses and informs efforts to engineer membrane-proximal Env presentation platforms for immunogen design ^47^.

Together, the cryo-EM structures define the molecular basis of two distinct non-CD4bs neutralizing specificities elicited by CH505 TF chTrimer vaccination. DH1744.1 and DH1745.1 recognized the V3 base through a conserved GDIR-centered, N332-independent mechanism with reduced dependence on the hypervariable V1 loop. In contrast, DH1708.1 targeted the gp120/gp41 interface through a previously uncharacterized FPPR-focused mode of recognition mediated exclusively by the heavy chain. Notably, engineering of VRC34 variants to expand recognition beyond the canonical fusion peptide epitope has been shown to enhance neutralization breadth ^48^, suggesting that the FPPR vulnerability identified here may represent an additional target for next-generation immunogen design.

### Single-cell multiomics identified transcriptomic correlates of CDRH3-dominant CD4bs response

Autologous tier 2 neutralizing Bmem cell frequencies varied substantially across participants (statistical framework detailed in **Methods**), with UIDs 123, 469, 185, 332, and 283 exhibiting the highest neutralizing antibody (nAb) responses. However, epitope-level analysis revealed distinct patterns of neutralization specificity. UIDs 123, 469, and 185 emerged as the strongest CD4bs responders based on both CD4bs neutralization frequencies and S365P-sensitive, CDRH3-dominant signatures, whereas UIDs 332 and 283 were dominated by V1/V3-directed responses. Notably, the exceptionally strong V1/V3-directed response in UID 332 (P*_adj_* = 4.2 x 10^-31^) likely masked a subdominant CDRH3-dominant component, such that the S365P-sensitive signature reached only marginal significance after multiple-testing correction (P*_adj_* = 0.0509; **Fig. 6A**). Nevertheless, the magnitude of the underlying CDRH3-dominant response supported inclusion of UID 332 among the high responders for subsequent cellular analyses. Among all participants, UID 123 exhibited the strongest enrichment of candidate CDRH3-dominant CD4bs precursor lineages, with an approximately 5-fold excess of S365P-sensitive CDRH3-dominant precursors relative to the cohort average. In addition, UID 123 generated N332-independent V1/V3-directed neutralizing antibodies and was the only participant in whom the gp120/gp41 interface-targeting lineage DH1708 was identified (**Figs. 1F and 4D**). The elicitation of these three types of nAbs in participant UID 123 motivated further longitudinal characterization of this participant’s vaccine-induced B-cell response.

**Figure 6.**
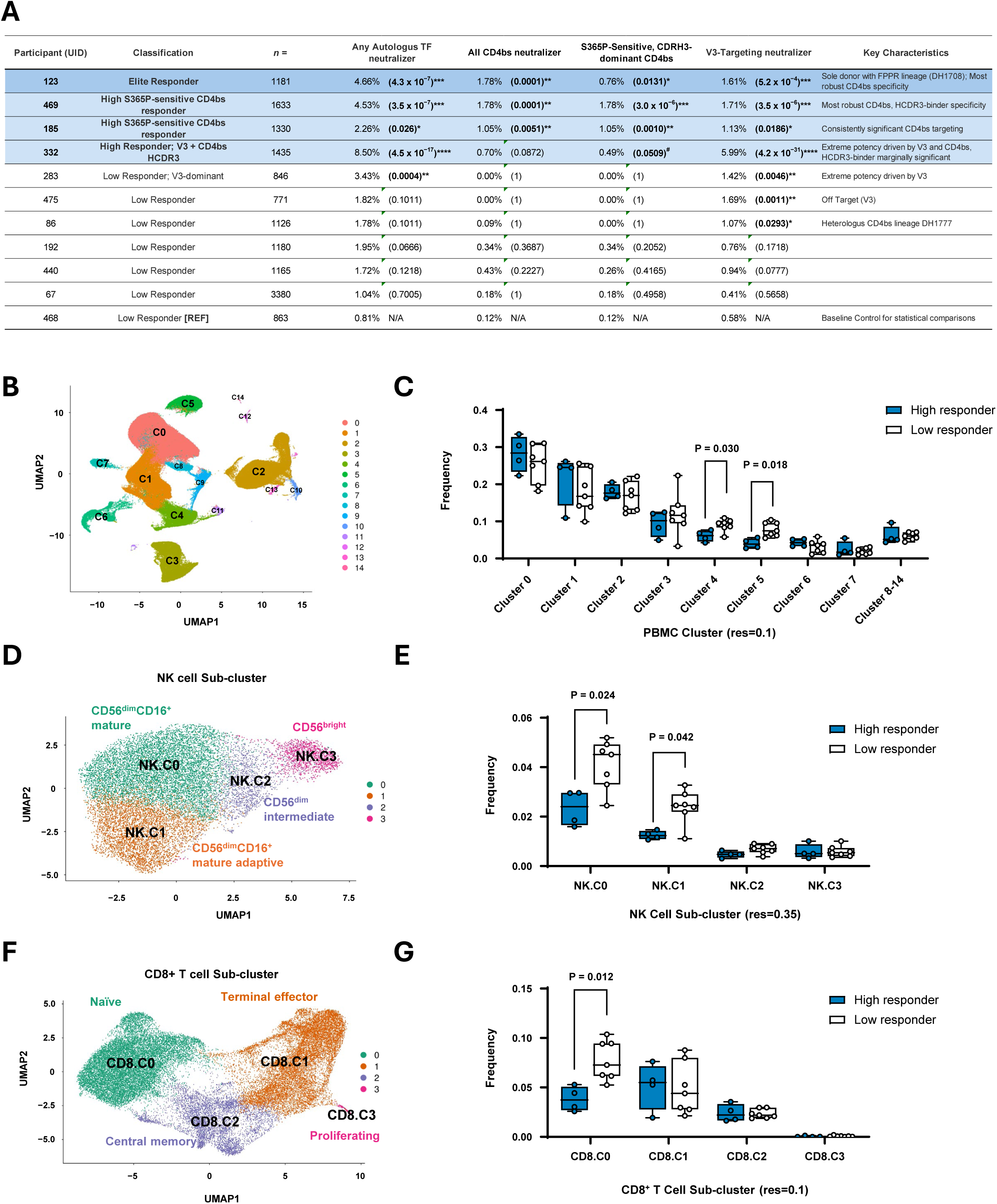
Participant-level autologous neutralizing antibody responses and PBMC/NK-cell features associated with S365P-sensitive CD4bs responder status. **(A) Participant-level classification of autologous neutralizing antibody responses recovered from CH505-reactive memory B cell cultures.** n denotes the total number of CH505-reactive memory B cell cultures analyzed per participant. Frequencies indicate the proportion of cultures within the CH505-reactive memory B cell compartment with the indicated neutralization or epitope-specific activity, including any autologous tier-2 CH505 transmitted/founder neutralization, total CD4bs-directed neutralization, S365P-sensitive CDRH3-dominant CD4bs neutralization, and V3-targeting neutralization. Adjusted P values were calculated by pairwise Fisher’s exact tests relative to the lowest responder/reference participant, followed by Benjamini–Hochberg correction for multiple comparisons. Blue shading highlights participants enriched for S365P-sensitive CDRH3-dominant CD4bs neutralizing activity. **(B) UMAP visualization of total leukapheresis-derived PBMCs profiled by single-cell transcriptomics across post-fourth immunization participant samples.** Unsupervised clustering identified major immune-cell populations at resolution 0.1. PBMC cluster identities were assigned using SingleR annotation based on the Monaco immune cell reference (main labels, see Methods and **Fig. S9A-B**). **(C) Frequencies of PBMC clusters in S365P-sensitive CDRH3-dominant CD4bs high versus low responders.** Box plots with overlaid individual participant values show each cluster as a proportion of total PBMCs. Clusters 8-14, for which frequencies did not exceed 0.05, are grouped. High responders are shown in blue and low responders in white. Two-sided Wilcoxon rank-sum tests were used for group comparisons; only significant p-values (<0.05) are shown. Low responders showed increased representation of selected PBMC clusters, including clusters corresponding to NK and CD8^+^ lymphocyte populations. **(D) UMAP sub-clustering of NK cells from total PBMCs.** NK cells were re-clustered at higher resolution, revealing four NK-cell subclusters: NK.C0: CD56^dim^CD16^+^ mature, NK.C1: CD56^dim^CD16^+^ mature adaptive, NK.C2: CD56^dim^ intermediate, and NK.C3: CD56^bright^ NK cells. NK subcluster annotations were supported by marker-gene expression profiles shown in **Fig. S9C**. **(E) Frequencies of NK-cell subclusters in S365P-sensitive CDRH3-dominant CD4bs high versus low responders, quantified as a proportion of total PBMCs.** Box plots with overlaid individual participant values show high responders in blue and low responders in white. Two-sided Wilcoxon rank-sum tests were used for group comparisons; only significant p-values (<0.05) are shown. Low responders showed a higher representation of CD56^dim^CD16^+^ NK cell subclusters, suggesting that the global NK cell enrichment observed in low responders may reflect increased expansion and/or survival of mature NK cell subpopulations. **(F) UMAP sub-clustering of CD8^+^ T cells from total PBMCs.** CD8^+^ T cells were re-clustered at higher resolution, revealing four transcriptionally and phenotypically distinct subclusters: CD8.C0: naïve, CD8.C1: Terminal effector, CD8.C2: Central memory, and CD8.C3: Proliferating CD8+ T cells. CD8^+^ T cell subcluster annotations were supported by marker-gene expression profiles and SingleR annotation based on the Monaco immune cell reference (fine labels, see Methods and **Fig. S9D-E**). **(G) Frequencies of CD8+ T-cell subclusters in S365P-sensitive CDRH3-dominant CD4bs high versus low responders, quantified as a proportion of total PBMCs.** Box plots with overlaid individual participant values show high responders in blue and low responders in white. Two-sided Wilcoxon rank-sum tests were used for group comparisons; only significant p-values (<0.05) are shown.

To define the full clonal architecture of the UID 123 neutralizing antibody response, we applied 10x Genomics BEAM to paired heavy- and light-chain BCR repertoires together with single-cell transcriptome profiling of the post-fourth immunization leukapheresis sample (**Fig. S8A−F**). BEAM-based reconstruction confirmed all neutralizing lineages identified by Nojima culture analysis and additionally resolved clonal members not captured in the primary culture screen, including 7 additional members of the DH1708 gp120/gp41 interface-targeting lineage and 30 additional members of the DH1595 CD4bs-directed lineage (**Figs. S8G−H**). These expanded lineage reconstructions provided a richer framework for defining maturation trajectories associated with vaccine-induced neutralization.

We next asked whether participants who mounted stronger vaccine-elicited CDRH3-dominant CD4bs neutralizing antibody responses exhibited distinct peripheral immune-cell states. We compared single-cell transcriptomic profiles of PBMCs from high responders (UIDs 123, 469, 185, and 332; **Fig. 6A**) and the remaining low responders. Unsupervised clustering identified responder-associated differences in two PBMC clusters, clusters 4 and 5, with frequencies that differed significantly (p<0.05, two-sided Wilcoxon rank-sum tests) between high and low responders (**Figs. 6B−C**). Based on annotated immune-cell identities, cluster 4 corresponded primarily to NK cells, whereas cluster 5 corresponded primarily to CD8^+^ T cells (**Figs. S9A−B**).

To further define the responder-associated NK-cell states, we subclustered the NK-cell compartment. Low responders were enriched for NK clusters C0 and C1 (**Figs. 6D−E**), corresponding to CD56^dim^CD16^+^ mature NK-cell states characterized by high expression of transcripts encoding cytolytic effector proteins, including perforin and granzyme B, with cluster C1 additionally exhibiting features of adaptive NK cells (**Fig. S9C**). Thus, stronger CDRH3-dominant CD4bs responses were associated with reduced representation of mature cytolytic NK-cell states in the peripheral blood.

Subclustering of the CD8⁺ T-cell compartment showed that the overall difference in CD8⁺ T-cell frequency was driven primarily by enrichment of a naïve CD8⁺ T-cell state in low responders (**Figs. 6E−F, S9D−E**), rather than by expansion of activated or cytotoxic effector CD8⁺ T-cell populations. The CD8⁺ T-cell association therefore primarily reflected differences in lymphocyte composition rather than a differential cytotoxic effector signature.

Together, these analyses identified distinct peripheral cellular correlates of vaccine-elicited CDRH3-dominant CD4bs responses. Stronger responders exhibited reduced representation of mature cytolytic NK-cell states and naïve CD8⁺ T cells, defining an immune landscape associated with more successful induction of CDRH3-dominant CD4bs neutralizing antibody responses.

## DISCUSSION

Here, we demonstrate that immunization with the CH505 TF chTrimer can simultaneously prime multiple neutralizing antibody pathways in humans, including responses targeting the CD4bs, the V1/V3 region, and the gp120/gp41 interface. This breadth of vaccine-elicited neutralizing specificities was revealed through unbiased functional screening of single memory B-cell cultures ^27,28^.

Recent clinical studies evaluating MPER peptide liposomes ^14,15^, multimeric engineered gp120 outer-domain (eOD-GT8) nanoparticles ^7,9^ and the Env trimer-based BG505GT1.1 ^8^, have established proof of concept that naïve B-cell-targeting immunogens can expand rare bnAb precursor populations in humans. Extending these observations, we found that a transmitted/founder Env immunogen derived from the virus that initiated the CH103 lineage simultaneously primed multiple neutralizing antibody classes, including candidate CDRH3-dominant CD4bs bnAb precursors, N332-independent V3-GDIR-directed antibodies, and a gp120/gp41 interface lineage. While heterologous breadth remained limited in this priming-only study, these findings demonstrate that a single Env immunogen can engage multiple bnAb-relevant developmental pathways in parallel.

The ability of CH505 TF chTrimer to induce multiple candidate bnAb precursor classes extends prior studies in which vaccine-elicited responses were primarily identified through serum epitope mapping or antigen-binding characteristics. In a clinical trial of native-like BG505 Env trimers, V1/V2/V3-directed serum antibody responses were detected in 5 of 17 participants by electron microscopy-based polyclonal epitope mapping, and two autologous neutralizing antibodies with similar Env mapping were isolated ^34^. Similarly, BG505 GT1.1, engineered to bind naïve B-cell receptors targeting the CD4bs and V2 apex, induced memory B cells with binding characteristics consistent with both specificities ^8,49^. In contrast, unbiased functional screening in HVTN 300 enabled direct isolation and characterization of multiple concurrent vaccine-elicited neutralizing lineages through binding, blocking, and structural analyses. Serum neutralization analysis further demonstrated CH505 tier 2 autologous neutralizing responses in 8 of 11 vaccinees ^26^. Together, these results demonstrate that CH505 TF chTrimer can template a polyepitope neutralizing antibody response in humans.

The identification of this multi-epitope neutralizing response was enabled by our high-throughput implementation of the Nojima limiting-dilution culture platform ^27,28^. Conventional single-cell antibody discovery approaches typically enrich antigen-binding B cells and, because downstream recovery, expression, and functional testing of individual antibodies are resource intensive, often further prioritize cells with predefined epitope-binding phenotypes or sequence features. Such strategies are highly effective for interrogating a specific antibody class but may under sample neutralizing responses outside the intended target. In contrast, the Nojima workflow broadly samples the vaccine-responding B-cell repertoire and directly screens antibodies secreted by individual B cells for neutralization activity before assigning epitope or antibody class. Neutralizing cultures can then be resolved by immunogenetic, binding, and structural analyses, allowing vaccine-elicited lineages to be classified according to function rather than selected a priori by presumed specificity. This function-first strategy enabled us to reconstruct the neutralizing antibody repertoire across multiple Env epitopes and to identify both anticipated and unanticipated vaccine-induced neutralizing lineages.

This approach allowed us to quantify the frequency of each neutralizing B-cell lineage and led to the unexpected identification of the VH1-2/Vκ1D-13 lineage DH1708, which targets the gp120/gp41 interface, in addition to the dominant N332-independent V3-GDIR-directed neutralizing response that comprised more than half of the neutralizing memory B-cell repertoire (1 in 9,716 total Bmem cells). Notably, the absence of the N332 glycan on CH505 TF may have redirected immunodominance toward N332-independent modes of V3 recognition, thereby revealing an alternative vaccine-accessible pathway to V3-directed neutralization.

The translational goal of the HVTN 300 clinical trial was to determine whether vaccination with a CH505 TF chTrimer, which originally initiated the prototype CDRH3-dominant CH103 lineage in a person living with HIV (PLWH) ^18^, could induce a polyclonal CD4bs bnAb candidate precursor response. Although antibodies recapitulating the CH103-like binding mode were elicited, the paucity of antibodies using the prototype VH4-59/Vλ3-1 pairing was consistent with this precise genetic solution being disfavored, as predicted ^18,23^. Instead, multiple genetically distinct lineages converged on CDRH3-dominant CD4bs recognition.

The DH1777 lineage further highlighted the structural diversity of vaccine-induced CD4bs responses. DH1777.1 was not S365P-sensitive, adopted a binding orientation distinct from CH103 and more closely resembling the CDRH3-dominant CD4bs bnAb b12 ^38^, and exhibited detectable heterologous neutralization, neutralizing 8 of 134 (6%) globally diverse HIV isolates. Its recovery illustrates that vaccination can access multiple structural solutions for CD4bs recognition and underscores the value of function-first repertoire analysis for capturing neutralizing responses that may fall outside predefined molecular signatures of the intended bnAb class.

The ability of an immunogen designed from a single bnAb lineage UCA to elicit a polyclonal response targeting the same structural class suggests that lineage-informed immunogen design can be generalized beyond an individual clonotype. Additional evidence for this generalizability comes from the observation that HIV Envs isolated from PLWH who developed bnAbs, when used to construct SHIVs, reliably induce polyclonal bnAb responses in SHIV-infected macaques targeting the same bnAb epitope identified in the original human donor ^50–53^. Thus, CDRH3-dominant putative bnAb precursors can be induced in humans, providing a starting point for the design of sequential boost immunogens to drive affinity maturation toward increased neutralization breadth and potency.

This study has provided data for iterative improvement of HIV Vaccine Trials Network (HVTN) HIV vaccine discovery medicine trials. The relatively low frequency of S365P-sensitive, CDRH3-dominant neutralizing antibody precursors (1 in 38,612 memory B cells) indicates that either priming immunogens with enhanced activation capacity or highly optimized sequential Envs for efficient boosting will be required to induce a reproducible, serum-dominant CDRH3-directed bnAb response. In this regard, studies of the CH505 PLWH identified Env variants that reacted with later members of the CDRH3-dominant bnAb lineage and may serve as boosting candidates ^20^; these variants are now being tested for their ability to boost CH103-like intermediate bnAbs in animal models. Moreover, in the new HVTN 312 clinical trial (NCT06557785), an mRNA-encoded CH505 TF Env is being paired with immunogens designed to target CD4-mimicking, CDRH2-dominant CD4bs bnAb lineages to simultaneously induce both CD4bs bnAb lineage types.

Current efforts are also focused on designing sequential boosting immunogens to increase the frequency of gp120/gp41 interface-directed precursor lineages and guide their subsequent affinity maturation. In parallel, the V1/V3-directed antibodies isolated in this trial define a structurally and genetically distinct class of neutralizing antibodies. These antibodies bind adjacent to the V1 loop and depend on the conserved Y330 residue and GDIR motif, rather than on the N301, N332, or N334 glycans typically required by canonical V3-glycan bnAb lineages. Although promiscuous glycan engagement may explain reduced dependence on individual glycans despite glycan contacts within the epitope ^42^, the antibodies identified here were characterized by dominant VH3/Vλ1-40 usage and N332-independent GDIR recognition. Importantly, N332-independent autologous neutralizing antibody lineages have previously been shown to evolve toward broad neutralizing activity ^31–33^. Thus, these immunodominant V3 base-targeting antibodies may represent a distinct vaccine-accessible pathway toward neutralization breadth, and ongoing studies are focused on defining immunogens capable of promoting their continued affinity maturation.

Finally, although heterogeneity in human vaccine responses has often been attributed to germline-encoded differences in the responding B cell repertoire, including Ig VH allelic variation ^8,14^, our current findings identify an additional host cellular correlate of vaccine responsiveness. Low responders in HVTN 300 exhibited higher frequencies of circulating NK cells, with enrichment of mature CD56^dim^CD16^+^ NK cells in the NK compartment, together with a CD8^+^ T cell compartment containing a higher proportion of naïve T cells. These features identify a host cellular state associated with weaker vaccine-elicited HIV CD4bs neutralizing antibody responses in humans. We have previously linked dysfunctional NK cells to enhanced bnAb development in PLWH ^54^. Here we showed that mature NK cells expressing high levels of transcripts encoding cytolytic effector proteins are associated with a low responder state of bnAb precursor expansion. This observation is consistent with recent systems vaccinology findings in older adults, where a baseline cytotoxicity-associated signature and increased CD16^+^ NK cell frequency were associated with weaker antibody responses to the T cell-dependent conjugate pneumococcal vaccine PCV13, but not to the T cell-independent polysaccharide vaccine PPSV23 ^55^. Together with experimental studies demonstrating that NK cells can limit Tfh and GC B-cell responses, including through perforin-dependent regulation of activated CD4^+^ T cells ^56,57^, these human data support a model in which excessive NK-cell cytotoxicity can constrain T-cell help and thereby disfavor vaccine-induced B-cell lineage expansion and maturation. These findings suggest that eliciting robust CD4bs neutralizing antibodies across individuals may require not only precise engagement of appropriate precursor B cells, but also immunization regimens that foster sufficient Tfh help to support their sustained expansion and maturation.

In summary, CH505 TF chTrimer vaccination primed multiple neutralizing antibody developmental pathways in humans, including CDRH3-dominant CD4bs, N332-independent V3-GDIR, and gp120/gp41 interface-directed lineages. These findings provide proof of principle that lineage-informed immunogen design can engage a broader spectrum of bnAb-relevant precursor populations than previously appreciated and establish multiple vaccine-accessible starting points for sequential immunization strategies aimed at eliciting broadly neutralizing HIV-1 antibodies.

### Limitations of the Study

This study has several limitations. First, the analysis was constrained by the relatively small number of participants with samples available for comprehensive B-cell repertoire analysis. Although 13 participants were initially enrolled, 11 completed sufficient immunizations to undergo leukapheresis, and only 9 received all four immunizations scheduled before leukapheresis. Second, HVTN 300 was designed as a priming-only study and did not include sequential heterologous boosting immunogens. We therefore could not directly determine whether the vaccine-elicited precursor lineages identified here would undergo further affinity maturation and acquire broad neutralization following additional immunization. Instead, their developmental potential was inferred from structural, genetic, and antigen-recognition features shared with established bnAbs or their inferred precursors and from previously defined maturation pathways associated with the acquisition of neutralization breadth ^18,58,59^.

## Competing Interests

SRW has received institutional grants or contracts from Sanofi Pasteur, Janssen Vaccines/Johnson & Johnson, Moderna Tx, Hookipa, Pfizer, AbbVie, Vir Biotechnology, and Worcester HIV Vaccine; has participated on data safety monitoring or advisory boards for Janssen Vaccines/Johnson & Johnson, Cyanvac, CSL Behring, and BioNTech; and his spouse holds stock/stock options in Regeneron Pharmaceuticals. EBW has received research funding from Pfizer, Moderna, Seqirus, Najit Technologies, and Clinetic for the conduct of clinical research studies. He has also received support as an advisor to Vaxcyte, Seqirus, and Pfizer, consultant to ILiAD Biotechnologies, and DSMB member for Shionogi and Emmes. BFH and KOS have patents on the CH505 Env immunogen in this study. Disclaimer: This article was written by some authors [LP] in their capacity as employees of the National Institutes of Health (NIH), but the views expressed herein do not necessarily represent those of the NIH.

## Data Availability

All data produced in the present study are available upon reasonable request to the authors

## Acknowledgment

This work was supported by the Department of Health and Human Services (HHS), National Institute of Allergy and Infectious Diseases (NIAID), and the National Center for Advancing Translational Sciences (NCATS) through grants UM1 AI068614 (to the HVTN LOC), UM1 AI068635 (to the HVTN SDMC), UM1 AI068618 (to the HVTN LC), UM1 AI069412 and UL1 RR025758 (to Harvard), U54 AI170752 (to the Duke Center for HIV Structural Biology [DCHSB]; PA), R01 AI145687 (to PA), UM1 AI144371 (to the Consortium for HIV/AIDS Vaccine Development [CHAVD]), and UM1 AI100645 (to the HHS, NIAID, DAIDS Center for HIV/AIDS Vaccine Immunology–Immunogen Design [CHAVI-ID]). Cryo-EM data were collected at the Duke Krios at the Duke University Shared Materials Instrumentation Facility (SMIF), a member of the North Carolina Research Triangle Nanotechnology Network (RTNN), which is supported by the National Science Foundation (award number ECCS-2025064) as part of the National Nanotechnology Coordinated Infrastructure (NNCI). We thank Dr. Nilakshee Bhattacharya for assistance with microscope alignments. This study utilized the computational resources offered by Duke Research Computing (http://rc.duke.edu; NIH 1S10OD018164-01) at Duke University. The authors thank Sommer Holmes, Benae Clardy, and Gabby Miner for technical assistance, Hsuan Su for illustration assistance and Whitney Beck for editorial assistance. The authors thank Duke School of Medicine for the use of the Sequencing and Genomic Technologies Core Facility for PacBio-based BCR sequencing. Purchase of the PacBio Revio system was funded by the NIH (1S10OD034222-01).

## Author contributions

Conceptualization: CHY, SRW, KOS, KW, DM, RJE, PA, LB, BFH

Investigation: CHY, RJP, QEZ, BT, KS, KL, HFK, EI, MC, LL, HIH, JL, AQ, YX, YC, RT, KM,TNS, HAL, RP, MB, RS, NL, AE, SS, KT, VI, GJ, PW, DC, AA, ED, IZ, MB, SV, JSM, KW, MSS

Methodology: SS, GHK, DC

Project administration: CHY, SRW, EBW, MJM, LC, KM, LB, BFH

Resources: SRW, WOH, OH, PCY, LLP, IT, CY, RB, JA, MJM, LC, LB

Supervision: MJM, LC, KM, WW, DM, LB, BFH

Visualization: CHY, RJP, QEZ, BT, KS, HFK, EI, MC, HIH, RP, MB, RJE

Writing – original draft: CHY, PA, BFH

Writing – review & editing: SRW, WOH, PB, WW, KOS, KW, RJE, LB

## KEY RESOURCES TABLE

## RESOURCE AVAILABILITY

### Lead contact

Further information and requests for resources and reagents should be directed to, and will be fulfilled by, the lead contact, Chen-Hao Yeh.

### Materials availability

All data supporting the findings of this study are available within the main article and the supplementary materials. Cryo-EM reconstructions of DH1530.3, DH1541, DH1708, DH1777.1, and DH1783 bound to the full-length CH505 TF HIV Env have been deposited in the Electron Microscopy Data Bank (EMDB) under accession numbers EMD-78071, 78074, 78496, 77833, 78500, 72878, 72829 and 77813. X-ray crystal structures have been deposited in the Protein Data Bank under accession codes PDB: 37CF, 37CI, 36TE, 37UR, 9YF6, 9YF7 and 36SE. Additional data relevant to this study will be made available upon request to the lead contact.

### Data and code availability

Results from additional data analyses have been deposited in Mendeley Data (repository link for review: https://data.mendeley.com/preview/94gp8439bz?a=4a4c7945-e5ab-49ad-9244-5b6811f94b52). Custom scripts used for single B-cell V(D)J sequencing analysis have been deposited in Zenodo (links in individual sections). Additional software and computational tools used for data processing and analyses are described in the Methods. Any further information required to reanalyze the data reported in this paper will be provided by the lead contact upon reasonable request.

## EXPERIMENTAL MODEL AND STUDY PARTICIPANT DETAILS

### Vaccine Participants

HVTN 300 (ClinicalTrials.gov: NCT04915768) is an open-label, phase 1 clinical trial. Part A was designed to enroll at least 12 healthy adult participants; Part B will be reported separately. Participants received 300 µg CH505 TF chTrimer administered intramuscularly, formulated with 5 µg 3M-052-AF and 500 µg alum (Access to Advanced Health Institute, Seattle WA) in a total volume of 1 mL. Vaccinations were administered on study day 0 (month 0), day 56 (month 2), day 112 (month 4), day 224 (month 8), and day 364 (month 12). Each dose was delivered as bilateral intramuscular injections (0.5 mL per deltoid) using a needle and syringe. Key eligibility criteria included age 18-55 years, good general health, HIV-uninfected status, low assessed risk for HIV acquisition, and willingness/ability to provide written informed consent. The study protocol was approved by the institutional review board at each participating site, and all participants provided written informed consent. A CONSORT diagram for HVTN 300 is provided in a companion manuscript ^26^.

### Ethical oversight

Ethical oversight and trial conduct were implemented in accordance with the standard operating procedures of the HVTN and the study sponsor (DAIDS). The protocol was reviewed and approved by the institutional review boards at all participating sites, and each participant provided written informed consent prior to enrollment. Following trial halting, the event underwent formal review by the Protocol Safety Review Team including the protocol chairs, protocol leadership, and clinical safety specialists—and by the HVTN Data and Safety Monitoring Board.

### Sample/participant identifiers

Numbers reported in this publication do not represent sample or participant identifiers. Instead, they correspond to HVTN-assigned Universal Identification (UID) codes, which provide an additional layer of de-identification beyond internal PTID and PUBID numbers. UID codes are specifically intended for use in presentations and publications.

### HVTN 300 Primary and Secondary Outcomes

#### Primary objectives and endpoints

*Primary objective 1:*

To evaluate the ability of the vaccine regimen to elicit CD4 binding-site, V2 apex and V3 glycan (bnAb region at the base of the V3 loop), and CH505TF-specific Bmem cells

*Primary endpoint 1:*

Flow cytometry analysis of the frequency of the CD4 binding-site, V2 apex and V3 glycan (bnAb region at the base of the V3 loop), and CH505TF-specific IgG+ B cells

*Primary objective 2*

To evaluate the ability of the vaccine regimen to elicit autologous tier 2 virus neutralizing antibodies

*Primary endpoint 2:*

Response rate and magnitude of serum antibody neutralization of vaccine-matched tier 2 HIV-1 strains as measured by TZM-bl assay

*Primary objective 3:*

To evaluate the safety and tolerability of a 300 mg dose of CH505 TF chTrimer admixed with different adjuvant regimens

*Primary endpoint 3:*

Local and systemic reactogenicity signs and symptoms will be collected for a minimum of seven days following receipt of any study vaccine.

#### Secondary objectives and endpoints

*Secondary objective 1:*

To evaluate HIV-1 binding antibody responses elicited by CH505 TF chTrimer admixed in 3M-052-AF + Alum or 3M-052-AF (without alum)

*Secondary endpoint 1:*

Response rate and magnitude of serum IgG binding antibodies as assessed by binding Ab multiplex assay (BAMA)

*Secondary objective 2:*

To evaluate the ability of the vaccine regimen to elicit HIV-1 specific heterologous tier 2 neutralizing antibodies

*Secondary endpoints 2:*

Response rate and magnitude of serum antibody neutralization of heterologous HIV-1 strains as measured by TZM-bl assay

## METHOD DETAILS

### Flow Cytometry Sorting of CH505.TF trimer-specific B Cells

Sorting of immunogen-specific Bmem cells was performed as previously described ^14^. Cryopreserved leukapheresis PBMCs were thawed and enriched for B cells using the EasySep™ Human Pan-B Cell Enrichment Kit (STEMCELL Technologies; Cat#19554) according to the manufacturer’s instructions. Enriched cells were stained with optimized concentrations of fluorochrome-conjugated monoclonal antibodies against human IgM (BV711; clone G20-127; BD Biosciences), IgD (BUV496; clone IA6-2; BD Biosciences), CD10 (PE-CF594; clone HI10A; BD Biosciences), CD3 (PE-Cy5; clone HIT3a; BD Biosciences), CD235a (PE-Cy5; clone GA-R2; BD Biosciences), CD16 (PE-Cy5; clone 3G8; BioLegend), CD14 (PE-Cy5; clone M5E2; BioLegend), CD27 (PE-Cy7; clone O323; Thermo Fisher), CD38 (Alexa Fluor 700; clone LS198-4-3; Beckman Coulter), CD19 (APC-Cy7; clone SJ25C1; BD Biosciences), IgG (BV605; clone G18-145; BD Biosciences), CD21 (BV650; clone 1048; BD Biosciences), CD11c (BUV396; clone B-Ly6; BD Biosciences), CXCR5 (BUV563; clone RF8B2; BD Biosciences), CD24 (BUV737; clone ML5; BD Biosciences), and CD20 (BUV805; clone 2H7; BD Biosciences).

Antigen baits were generated as 4:1 molar mixture of biotinylated CH505 TF chTrimer immunogen with Streptavidin-VioBright 515 (Miltenyi Biotec) or Streptavidin-Alexa Fluor 647 (Thermo Fisher). Dead cells were excluded using 7-AAD (BD Biosciences). Bmem cells were defined as the union of two populations after exclusion of dead cells and non-B lineages: (i) 7-AAD⁻CD14⁻CD16⁻CD3⁻CD235a⁻CD20⁺CD27⁺ and (ii) 7-AAD⁻CD14⁻CD16⁻CD3⁻CD235a⁻CD20⁺CD27⁻IgD⁻. Within this combined Bmem cell gate, cells binding both VB515-CH505 TF and AF647-CH505 TF baits were single-cell sorted on a BD FACSymphony S6 (BD Biosciences) using FACSDiva software version 8. Data were analyzed using FlowJo version 10 (BD Biosciences).

### Human Nojima Single B-cell Cultures

Nojima single-cell B cell cultures were performed as previously described, with minor modifications ^28^. MEC-147 feeder cells were maintained in IMDM-10 (IMDM; Thermo Fisher, Cat#12440053) supplemented with 10% FBS (Cytiva, Cat#SH30071.03E), 55 µM 2-mercaptoethanol (Thermo Fisher, Cat#21985023), penicillin (100 U/mL), and streptomycin (100 µg/mL) (Thermo Fisher, Cat#15140122). Twenty-four hours prior to single-cell sorting, feeder cells were detached with 0.05% Trypsin-EDTA (Thermo Fisher, Cat#15400054), resuspended in pre-warmed B cell culture medium (BCM; RPMI-1640 (Thermo Fisher, Cat#11875119) supplemented with 10% FBS (Cytiva, Cat#SH30071.03E), 10 mM HEPES (Thermo Fisher, Cat#15630080), 1 mM sodium pyruvate (Thermo Fisher, Cat#11360070), 1x MEM non-essential amino acids (Thermo Fisher, Cat#11140050), 55 µM 2-mercaptoethanol (Thermo Fisher, Cat#21985023), penicillin (100 U/mL), and streptomycin (100 µg/mL) (Thermo Fisher, Cat#15140122)), and seeded into 96-well flat-bottom plates (Corning, Cat#3595) at 3,000 cells per well in 200 µL. CH505 TF-double positive Bmem cells were single-cell sorted by flow cytometry directly into feeder cell plates at one cell per well. On day 6, 100 µL of BCM was added to each well (total volume 300 µL thereafter). Medium was replaced on days 12, 17, and 21. On day 21, 200 µL of culture supernatant was collected for indirect binding ELISA. On day 25, supernatants were harvested for total IgG ELISA and TZM-bl neutralization assays. Following supernatant collection, plates containing clonally expanded B cells were frozen at −80°C for subsequent RNA extraction and V(D)J sequencing.

### ELISA Screening of IgG-Producing Clonal B Cell Cultures

Standard sandwich ELISAs were performed to identify IgG-producing B cell clones from single-cell culture supernatants. High-binding 384-well plates (Corning, Cat#3700) were coated overnight at 4°C with goat anti-human Igκ (2 µg/mL; polyclonal; SouthernBiotech, Cat#2061-01) and goat anti-human Igλ (2 µg/mL; polyclonal; SouthernBiotech, Cat#2071-01) diluted in carbonate coating buffer (100 mM NaHCO₃, 2.5 mM Na₂CO₃). Plates were washed three times with PBS containing 0.1% Tween-20 using a BioTek ELx405 plate washer and blocked with 100 µL/well blocking buffer (PBS, 0.5% BSA) for 2 h at room temperature or overnight at 4°C. After blocking, 55 µL of blocking buffer was removed and 5 µL of culture supernatant was added per well (final dilution 1:10). Plates were incubated for 2 h at room temperature or overnight at 4°C, washed three times, and incubated with goat anti-human IgG-HRP (1:5,000; polyclonal; SouthernBiotech, Cat#2040-05) for 1 h at room temperature in the dark. Plates were washed three times, developed with TMB substrate (BioLegend, Cat#421101) for 5 min at room temperature in the dark, and stopped with 1 M H₃PO₄ (Sigma-Aldrich). Absorbance was measured at 450 nm. Supernatants from feeder-only wells (no sorted B cells) served as negative controls. Wells were scored as IgG-positive when OD₄₅₀ values exceeded the negative-control mean plus six standard deviations (mean + 6xSD). IgG-positive culture supernatants were transferred to master plates for downstream binding and functional assays.

### Antigen-Specific Antibody Indirect Binding ELISAs

Antibody binding of culture supernatants and recombinantly expressed monoclonal antibodies was measured by ELISA in 384-well high-binding plates (Costar, Cat#3700) coated overnight at 4°C with either streptavidin (2 µg/mL; Thermo Fisher Scientific, Cat#S-888) or protein antigen diluted in 0.1 M sodium bicarbonate. Plates were washed with PBS containing 0.1% Tween-20 (PBS-T) and blocked for 1 h with assay diluent (PBS supplemented with 4% (w/v) whey protein, 15% normal goat serum, 0.5% Tween-20, and 0.05% sodium azide). For SOSIP gp140 ELISAs, streptavidin-coated plates were incubated with biotinylated SOSIP gp140 (2 µg/mL in assay diluent) for 1 h, followed by washing.

Culture supernatants or monoclonal antibodies were added in 10 µL volumes and incubated for 1 h at room temperature. Supernatants were added to a single well at the original concentration while monoclonals were added in 3-fold serial dilutions starting at 100 µg/mL. Plates were washed and incubated for 1 h with HRP-conjugated goat anti-human secondary antibody (Jackson ImmunoResearch, Cat#109-035-098) diluted 1:15,000 in assay diluent lacking sodium azide. Plates were washed and developed with SureBlue Reserve TMB substrate (SeraCare, Cat#5120-0083) (20 µL/well) for 15 min, and reactions were stopped with 20 µL HCl stop solution. Absorbance was read at 450 nm on a SpectraMax Plus 384 plate reader. Data were acquired using SoftMax Pro (v5.3) and collated/visualized in Microsoft Excel and GraphPad Prism (v9).

### TZM-bl neutralization assays

Neutralization assays were performed using TZM-bl cells as previously described ^60,61^. Neutralizing activity of culture supernatants and mAbs was tested against Env-pseudo-typed viruses expressing autologous CH505 Env variants, including the transmitted-founder CH505 TF, engineered glycan-modified variants (CH505TF.gly4, CH505TF.gly4.S365P.2, CH505TF.G458Y.N279K/GnT1⁻, and CH505TF.G458Y.N279K.N280D/GnT1⁻), and an early longitudinal variant (CH505.w4.3).

S365P-sensitive, CDRH3-dominant activity was defined by a >3-fold shift in IC50 between CH505.gly4 and CH505.gly4.S365P.2 ^35^. Pseudoviruses were generated by transfection of 293T/17 or 293S/GnT1⁻ cells ^60^. Briefly, 90 µL of culture supernatant or mAb dilution was incubated with 45 µL of pre-titrated virus for 1 h at 37°C. Freshly trypsinized TZM-bl cells (110 µL) containing an optimized concentration of DEAE-dextran were added, and plates were incubated for 48 h at 37°C. Infection was determined by measuring Tat-regulated firefly luciferase activity ^60^. Neutralization titers were calculated as the percent reduction in relative luminescence units (RLUs) compared to virus-only controls after subtraction of background RLUs from cell-only controls. Assay specificity was confirmed using the SVA-MLV negative control. Heterologous neutralization breadth was assessed for DH1530 and DH1541 lineages, as well as DH1777.1 and DH1708.1 mAbs, using the global panel ^62^. Additionally, DH1777.1 was tested against a multi-clade panel of 119 HIV-1 Envs ^63^.

### Comparison of Nojima Cultures with Conventional Single-Cell Sorting for BnAb Precursor Isolation

To benchmark the Nojima culture platform against conventional approaches, we directly compared the two methods on paired post-fourth-immunization (week 34) leukapheresis samples from four HVTN 300 participants (UID 123, 185, 469, 475) with serum CD4bs-directed neutralization signatures ^14,26^. The conventional workflow comprised fluorophore-labeled Env single-cell sorting followed by single-cell RT-PCR recovery and recombinant IgG expression. Given the limited throughput of this approach, it was implemented as a targeted CD4bs-enrichment strategy using differential binding to CH505.M11 versus the CDRH3-dominant knock-out variant CH505.M11.STG (STGKO), rather than broadly sampling the full CH505 TF-reactive Bmem repertoire (**Figs. S1F−H**) ^58^. Of 1,922 single B cells sorted by the conventional approach, 1,068 VH/VL pairs were recovered, yielding 41 CD4bs-directed mAbs and 11 autologous tier 2 CH505 TF neutralizers (**Figs. S1F−H**). Within the CDRH3-dominant-enriched compartment, Nojima cultures recovered all three CD4bs-directed neutralizing lineages identified by conventional sorting and identified seven additional lineages not captured by the targeted workflow, representing a more than three-fold increase in lineage recovery. Critically, because conventional sorting was prospectively restricted to the CD4bs compartment, it necessarily missed capturing the V3-glycan- or gp120/gp41 interface-directed neutralizing lineages that constituted the majority of the CH505 TF-elicited responses. Together, these results establish the Nojima culture platform as a preferred approach for unbiased, epitope-unrestricted profiling of vaccine-elicited neutralizing repertoires, enabling both deeper sampling within a targeted specificity and comprehensive recovery across the full spectrum of Env-directed responses.

### RNA extraction for BCR V(D)J sequencing

For IgG-positive single-B cell culture wells, RNA was isolated from frozen cell pellets retained in the corresponding original culture wells. High-throughput extraction was performed using the Quick-RNA 96 Kit (Zymo Research, Cat. #R1053).

### PacBio BCR V(D)J sequencing of Nojima culture-derived B cells

RNA extracted from cultured clonal B cells was reverse-transcribed to generate full-length immunoglobulin V(D)J cDNA using SMART template-switching technology to enable unbiased 5’ capture. Briefly, RNA was combined with dNTPs (NEB, Cat. #N0447L), a template-switching oligonucleotide (TSO; containing 5’ padding and a plate-level barcode), and constant-region reverse primers specific for IgG, Igκ, and Igλ. Reactions were denatured at 72°C for 3 min, chilled on ice for 2 min, and supplemented with an RT mix containing 5x RT buffer (final 1x), 20 mM DTT (final 2 mM), RNaseOUT™ RNase Inhibitor (Thermo Fisher Scientific, Cat. #10777-019; final 1x), and SMARTScribe™ Reverse Transcriptase (Takara, Cat. #639538). Reverse transcription was performed at 42°C for 90 min followed by heat inactivation at 70°C for 15 min. Final concentrations were 0.5 mM each dNTP, 1.0 µM TSO, and 0.2 µM each constant-region reverse primer.

Full-length V(D)J amplicons were generated by two rounds of semi-nested PCR. In PCR1, a common forward primer complementary to the TSO was paired with a constant-region reverse primer (IgG, Igκ, or Igλ) in separate reactions. In PCR2, products were re-amplified using (i) a set of twelve forward primers targeting the PCR1 forward priming site and incorporating 5’ padding plus a column barcode, and (ii) class-specific semi-nested reverse primer sets (eight primers per class) binding upstream of the corresponding PCR1 reverse priming sites and incorporating 5’ padding plus a row barcode. This 12 x 8 dual-indexing scheme uniquely barcoded each well of a 96-well plate.

PCR1 and PCR2 were performed either with Herculase II Fusion DNA Polymerase (Agilent, Cat. #600679) in 1x Herculase Reaction Buffer containing 0.2 mM each dNTP and 0.2 µM each primer, using template at 1/10 reaction volume and polymerase at 1/50 reaction volume, or with PrimeSTAR GXL DNA Polymerase (Takara, Cat. #R050B) in 1x PrimeSTAR Reaction Buffer containing 0.1 mM each dNTP and 0.3 µM each primer, using template at 1/10 reaction volume and polymerase at 1/50 reaction volume. For the Herculase condition, cycling parameters for PCR1 (and human PCR1) were 95°C for 4 min; 30 cycles of 95°C for 30 s, 55°C for 20 s, and 72°C for 30 s; followed by 72°C for 10 min; for human PCR2, the annealing temperature was increased to 59°C (all other parameters unchanged). For the PrimeSTAR GXL condition, cycling parameters for PCR1 (and human PCR1) were 95°C for 1 min; 30 cycles of 98°C for 10 s, 55°C for 15 s, and 68°C for 60 s; followed by 72°C for 10 min; for human PCR2, the annealing temperature was increased to 60°C (all other parameters unchanged).

PCR2 products were pooled per original 96-well RNA plate, combining IgG, Igκ, and Igλ amplicons. Pooled libraries were size-selected/gel-purified using a BluePippin system (Sage Science, Cat. #BDF1510), ethanol-precipitated, and eluted in 10 mM Tris-HCl (pH 8.0). Libraries were quality controlled, converted to SMRTbell® templates using the SMRTbell® Prep Kit 3.0 (PacBio, Cat. #102-182-700), indexed with the SMRTbell® Adapter Index Plate 96A (PacBio, Cat. #102-009-200) and sequenced on the PacBio Revio platform.

### Immunogenetic Analysis of PacBio V(D)J Sequencing of Nojima culture-derived B cells

PacBio circular consensus sequencing (CCS) reads were demultiplexed, processed, and immunogenetically annotated using Abulous (https://github.com/WieheLab/Abulous; DOI:5281/zenodo.21828611), a custom Python program developed as an updated version of our previously described Absolute software ^28^. The Abulous workflow consists of the following steps: (1) reads are preprocessed for quality control by removing reads outside the expected amplicon length range of 400-800 nucleotides. (2) Sequencing reads are assigned to individual wells from single B cell culture plates by matching subsequences to the corresponding row and column barcodes and to chain type by matching to chain-specific primers (3) For each well and chain type pair, sequences are clustered based on a 210 nucleotide region starting 30 nucleotide upstream (5′) of the chain barcode which includes the CDR3, with group membership defined by a 90% sequence identity threshold and sequences below this threshold initiating new clusters; (4) clusters containing at least 5 sequences are then aligned with MAFFT ^64^ to generate a secondary consensus sequence, defined by the most frequent nucleotide at each position; (5) secondary consensus sequences were then aligned to each other, and those sharing ≥90% identity were merged and re-aligned to generate a final refined consensus sequence, following the procedure described in step 4.

Final refined consensus sequences were then grouped by participant and chain, annotated with their immunogenetic information, and partitioned into clonal families using the immunogenetic analysis software package Partis ^65,66^ with a custom pre-trained parameter directory based on the OGRDB human germline reference ^67^(version 9; DOI:10.5281/zenodo.13923288). If more than one sequence for a given chain type was annotated within a well, sequences supported by fewer than 10 CCS reads were discarded. If multiple sequences remained, indicating the presence of more than one heavy or light chain in the well, additional filtering was applied as follows: if all remaining sequences shared the same V allele, only the sequence with the highest number of supporting reads was retained; otherwise, all productive and functional sequences were retained. Sequences were classified as productive if they lacked premature stop codons, and as functional if they contained an in-frame CDR3, expected invariant residues were present, and any insertions or deletions occurred in multiples of three nucleotides. Clonal partitioning was performed using heavy chain sequences. For specific clones of interest in which two different light chains were detected, we re-sequenced these wells to verify the correct light chain pairing.

For comparisons between the pre-vaccination V(D)J repertoire generated by 10x Genomics sequencing and Nojima culture-derived V(D)J sequences generated by PacBio sequencing, the two datasets were integrated on a per-donor basis, immunogenetically annotated, and clonally partitioned. For Nojima-derived clonal groups in which multiple light-chain V genes were detected, only sequences using the predominant light-chain V gene within that clone were retained. For analyses normalized at the clonal level, one representative sequence was selected from each clone. Code used for annotation of the Nojima culture-derived sequences and generation of the heatmaps is available at Zenodo (DOI:10.5281/zenodo.21828021).

### PCR Isolation of Antibodies from Antigen-Specific B Cells

Single antigen-specific B cells were sorted into 96-well plates containing lysis buffer and stored at −80°C as previously described ^68,69^. Immunoglobulin heavy and light chain gene segments were recovered by single-cell PCR ^68^. Sequence data were processed using a custom bioinformatics pipeline for base-calling, contig assembly, quality trimming, and immunogenetic annotation using Cloanalyst (https://www.bu.edu/computationalimmunology/research/software/). Functional V(D)J sequences were identified and isotyped as described ^70^. Validated heavy and light chain variable region sequences of antibodies have been deposited in Mendeley Data.

### Single-cell BCR repertoire library construction and 10x Genomics sequencing of pre-vaccination samples

For pre-vaccination BCR repertoire analysis, B cells were enriched from pre-vaccination HVTN 300 leukapheresis PBMC samples using a commercially available negative-selection B-cell enrichment kit (STEMCELL Technologies). Single-cell VDJ libraries were generated using the 10x Genomics Chromium Controller (PN-1000204), Chromium Next GEM Single Cell 5’ Kit v2 (PN-1000263), and BCR Amplification Kit (PN-1000253) according to the manufacturer’s protocol. Briefly, 16,000–18,000 enriched B cells were loaded per sample to target recovery of approximately 10,000 cells. Libraries were quantified using TapeStation (Agilent) and Qubit 3 Fluorometer (Thermo Fisher) and sequenced on an Illumina NovaSeq 6000 using a NovaSeq S4 200-cycle reagent kit with 26 cycles for Read 1, 10 cycles for i7 index, 10 cycles for i5 index, and 150 cycles for Read 2. VDJ libraries were sequenced to a target depth of 5,000 reads per cell.

### Single B-cell barcode-enabled antigen mapping (BEAM) by 10x Genomics sequencing

To isolate antigen-reactive B cells, antigen assemblies were generated comprised of antigen and a BEAM Conjugate that contains Biolegend TotalSeq-C barcoded streptavidin molecule linked with a PE or APC fluorescence marker. The BEAM Assemblies served as B cell hooks to capture antigen-reactive B cells, and were produced according to the 10X Genomics Barcode Enabled Antigen Mapping Workbook for BEAM-Ab Assembly (Doc CG000597 Rev A). The biotinylated proteins used to make BEAM Conjugates were HIV-1 Env antigens: CH505TF SOSIP (TotalSeq-C0961 PE; Cat #: 405155, and TotalSeq-C0956 APC; Cat #: 405283), CH505.M11 SOSIP (TotalSeq-C0962 PE; Cat #: 405153, and TotalSeq-C0957 APC; Cat #: 405285), CH505.M11-STG SOSIP (TotalSeq-C0964 PE; Cat #: 405297, and TotalSeq-C0958 APC; Cat #: 405293) CH505.M5.G458Y SOSIP (TotalSeq-C0965 PE; Cat #: 405295, and TotalSeq-C0959 APC; Cat #: 405159), and CH505.M5.G458Y.K282D-STG SOSIP (TotalSeq-C0966 PE; Cat #: 405161, and TotalSeq-C0960 APC; Cat #: 405157). A non-relevant HIV-1 antigen Human Serum Albumin assembly (TotalSeq-C0967 PE; Cat #: 405163, and TotalSeq-C0976 APC; Cat #: 405195) was prepared as a negative control for background subtraction (see BEAM scoring). Two BEAM Assemblies (PE and APC) were produced for each antigen using unique barcodes. Antibody-coated beads (negative bead control Influenza HA-reactive mAb CH65; positive control beads: HIV-1 Env reactive mAbs PGT151) were used to confirm expected binding and fluorescent profiles of BEAM assemblies by flow cytometry.

Cryopreserved leukapheresis vials (25 vials at 2×10^7^ cells per vial) from UID 123 were thawed at 37°C and slowly diluted in 1mL RPMI with 10% FBS (complete RPMI, cRPMI) supplemented with Benzonase at 25 U/mL. After 1 minute, the cell suspension was further diluted with 13mL cRPMI. Cells were washed via centrifugation for 5 minutes at 400rcf, then resuspended in cRPMI with 5 μM Chk2 inhibitor (Millipore Sigma; Catalog #220486) at 2X10^6^ cells/mL final concentration based on the frozen cell count. After cell resuspension, an aliquot (10 µL) was taken for cell count verification using the Countess 3 Automated Cell Counter (Thermo Fisher). Following centrifugation, the cells were resuspended in FACS buffer (1% BSA in 1X PBS) with 5μM Chk2 inhibitor and CD4 blocking antibody, Clone SK3 (Biolegend; Cat #: 344602; 1:100 dilution) on ice for 30 minutes at a final concentration of 1X10^6^ cells/100µL. An aliquot of 0.4-0.5×10^6^ cells was set aside as a negative control without BEAM antigens to determine antigen-specific sort gating, but stained with the rest of the panel. The remaining cells were stained with surface antibodies: CD3 PE-Cy5 (clone HIT3a; BD Biosciences Cat #: 555341; 1:40), CD235a PE-Cy5 (clone GA-R2; BD Biosciences Cat #: 559944; 1:150), CD16 PE-Cy5 (clone 3G8; Biolegend Cat #: 302010; 1:200), CD14 PE-Cy5 (clone M5E2; Biolegend Cat #: 301864; 1:80), CD27 PE-Cy7 (clone O323; eBiosciences Cat #: 25-0279; 1:20), CD38 APC-AF700 (clone LS198-4-3; Beckman Coulter Cat #: 6699531; 1:100), CD19 APC-Cy7 (clone SJ25C1; BD Biosciences Cat #: 557791; 1:40), IgG BV605 (clone G18-145; BD Biosciences Cat #: 563246; 1:80), and IgM BV711 (clone G20-127; BD Biosciences Cat #: 740795; 1:100) for flow sorting and BEAM Assemblies for 30 minutes on ice. Cells were stained with the antibody staining panel in FACS buffer at a final concentration of 1X10^6^ cells/100µL. Following cell staining, cells were washed with 1X PBS, then resuspended in 1X PBS with 5 μM Chk2 inhibitor and Aqua vital dye (Thermo Fisher; Cat #: L34957; 1:1000) for 30 minutes on ice at a final concentration of 1X10^6^ cells/100µL. Cells were then washed with FACS buffer and, finally, resuspended in 100µL per 1X10^6^ cells in FACS buffer with 5 μM Chk2 inhibitor and kept on ice. Live CD19^+^IgG^+^ antigen-reactive B cells were sorted into 500µL cRPMI with 5 μM Chk2 inhibitor in a FACS tube using a 100 μm nozzle on a BD FACSymphony S6 (DHVI Flow Cytometry core). Sorted cells were kept on ice for no more than 1 hour and then centrifuged at 400 xg for 5 minutes. The supernatant was slowly aspirated and discarded while leaving behind ∼50-200µL of residual cells in buffer to target 400-2000 cells/μL as per 10X Genomics recommendations.

GEM derived cDNA from sorted antigen-reactive B cells populations from UID 123 were used to generate VDJ, gene expression, and BEAM barcode libraries using Chromium GEM-X single Cell 5’ v3 Reagent Kits (10X Genomics; Catalog #PN-1000695) with Feature Barcode technology for BEAM per the manufacturer’s recommendations (10X Genomics, Document #CG000591 Rev A). For single cell encapsulation, we input all the cells recovered as quantified above into 2 lanes of the 10X Genomics CHIP, thus allowing us to target 20,000 cells per lane. After preparation of the gene libraries, library QC was performed using TapeStation 4200 with the high sensitivity D1000 or D5000 ScreenTape (Agilent Technologies, Cat #: 5067-5582) and Qubit Flex fluorometer (Thermo Fisher Scientific). The qualified VDJ, gene expression and BEAM libraries were pooled at 1:10:1 ratio, and sequenced with Novaseq X Plus 10B 100 cycles kit (Illumina) to achieve the sequencing depth of ≥5,000 read pairs/cell for VDJ libraries, ≥50,000 read pairs/cell for 5’ gene expression libraries, and ≥5,000 read pairs/cell for BEAM libraries following the manufacturer’s protocol (Illumina, Document #200027171 v02). For Illumina NGS, we loaded concentration of libraries at 150pM and 1% Phix (Illumina, Cat #: FC-110-3001).

### Processing of 10x Genomics V(D)J, Feature Barcode, and Gene Expression Sequencing Data

Immune profiling and transcriptomic data were processed using the Cell Ranger Single Cell Software Suite. Samples were demultiplexed, quality filtered, assembled, and aligned to a human VDJ reference with immunoglobulin genes or to the human transcriptome for GEX, followed by UMI counting. Assembled VDJ contigs and chain annotations generated by Cell Ranger were used as input for downstream BCR analysis using the Cloanalyst software package. Immunogenetic annotations were assigned using the default Cloanalyst human immunoglobulin library, and clonal partitioning was performed using a custom analysis pipeline. Paired unmutated common ancestor (UCA) inference and clonal lineage analyses were also conducted using Cloanalyst.

BEAM analysis was performed using the Cell Ranger Software Suite (version 9.0.1) and custom scripts. Demultiplexed FASTQ files were generated with the mkfastq command and processed using the count command, along with a feature reference file specifying the barcodes of oligonucleotide-tagged antigens. The resulting feature barcode count matrix was then used to calculate BEAM scores as described below.

#### BEAM-Ab scoring prediction analysis

The processed BEAM-Ab feature barcode library as described above was used to generate BEAM Scores. These scores provide a quantitative measure of antigen binding and were used to identify HIV-1 Env-reactive BCRs. BEAM Scores were calculated using the count matrices containing unique molecular identifier (UMI) barcode counts for each cell, target antigen, and negative control antigen. Scores represent the likelihood of antigen binding relative to the negative control and were computed using beta distribution modeling, defined as 100 times the probability that a given BCR binds a target antigen at a confidence level of at least 92.5%. Scores range from 0 to 100, where 0 predicts no target antigen binding relative to the negative control and 100 indicates a high likelihood of binding relative to the negative control. To improve reliability of B cell capture and confidence in scoring, two independent BEAM assemblies were generated for each HIV-1-reactive antigen using distinct barcodes, and UMI counts from dual-barcoded antigens were averaged for score calculations.

For downstream analysis, cells with a sum of BEAM scores across all panel antigens greater than 10 were retained as exhibiting appreciable antigen reactivity. Within this antigen-reactive population, STG variants of each immunogen — each incorporating the three gp120 core substitutions S365K, T455E, and G459E that selectively abrogate CD4bs broadly neutralizing antibody recognition ^71^ served as antigen-specificity negative controls. Cells with BEAM scores exceeding 25 against an STG variant were considered to lack the epitope specificity defined by the corresponding native antigen and were excluded from the relevant BnAb-precursor category. This STG-based exclusion threshold of less than 25 is distinct from the non-relevant protein (HSA) exclusion threshold of less than 20 used for general IgG specificity filtering.

BnAb-precursor-class specificities were defined by differential BEAM score thresholds anchored to canonical lineage binding signatures. CH103-like B cells were defined as IgG⁺ cells with a CH505.M11 BEAM score greater than 50 and a CH505.M11-STG BEAM score less than 25. CH235-like B cells were defined as IgG⁺ cells with a CH505.M5.G458Y BEAM score greater than 50 and a CH505.M5.G458Y.K282D-STG BEAM score less than 25. VRC01-like B cells were defined as IgG⁺ cells with a CH505TF BEAM score greater than 50 and a CH505.M5.G458Y BEAM score less than 25. By these criteria, 877 VRC01-like, 304 CH103-like, and 67 CH235-like B cells were identified.

Previous BEAM experiments have informed our working criteria for selection of BCRs from BEAM-Ab for Ab expression. Our criteria for IgG Abs are as follows: heavy/light chain UMIs ≥ 5, BEAM score > 50 for HIV-1 antigens, BEAM score < 20 for non-relevant HSA protein.

### PBMC single-cell gene expression library production and 10x Genomics sequencing

To generate PBMC single-cell transcriptomic profiles from all post-fourth immunization participant samples (n=11), cryopreserved leukapheresis PBMCs were thawed, washed in ice-cold RPMI supplemented with 10% FBS, and counted. Single cells were encapsulated using the 10x Genomics Chromium X (10x Genomics, Pleasanton, CA), and gene expression libraries constructed with the 5’ GEM-X chemistry (PN 1000699). Libraries were quantified using a TapeStation 2200 (Agilent, Santa Clara, CA) and a Qubit Fluorometer (Thermo Fisher Scientific, Waltham, MA). Paired-end sequencing was performed on the Illumina NextSeq 2000 platform (Illumina, San Diego, CA).

### Single-cell gene expression analysis of PBMCs

GEX count matrices from the 11 post-fourth immunization PBMC samples were analyzed in R using Seurat. For each sample, cells were retained if they expressed between 200 and 6,000 genes, and fewer than 15% mitochondrial transcripts, to exclude low-quality cells and likely doublets, resulting in a total of 235,110 cells across the 11 samples. Gene expression counts were log-normalized, scaled and centered prior to PCA. To correct for sample technical variation, canonical correlation analysis (CCA)-based integration was applied using IntegrateData ^72^. Cells were clustered on a shared nearest-neighbor (SNN) graph (FindNeighbors) using the Louvain algorithm (FindClusters, resolution = 0.1) and visualized by UMAP. Automated cell type annotation was also performed using SingleR ^73^ with the Monaco immune cell reference ^74^. Cluster composition was compared between high responders and low responders. NK cell- and CD8+ T-cell annotated clusters showed frequency differences, therefore these populations were analyzed further separately based on the associated SingleR labels. Within each subset (15,025 NK cells, 33,313 CD8+ T cells), clusters expressing markers indicative of a different lineage or low-quality metrics were excluded. Each cleaned subset was processed independently: cells were reclustered (FindClusters, resolution = 0.35 for NK, 0.1 for CD8) and subclusters were annotated by manual inspection of marker genes identified with FindAllMarkers (**Figs. S9C−D**) and cross-referenced against published human NK cell ^75^ CD8^+^ T cell ^76^ and PBMC ^76^ atlases. Data visualization was performed using Seurat and ggplot2.

### Recombinant Antibody Production

PCR amplicons of antibody heavy and light chain genes from single B cells were used to generate linear expression cassettes as described ^24,68^. Cassettes were transfected into Expi293F cells (Thermo Fisher) using ExpiFectamine to produce microgram quantities of IgG. Three days post-transfection, culture supernatants were harvested by centrifugation, 0.22 μm filtered, and screened for HIV-1 envelope binding via ELISA, with a positivity cutoff of OD450>0.5. The variable region of selected heavy and light chain genes were synthesized and cloned into human IgL or IgG1 expression vectors (GenScript). Plasmids were prepared using Midi or Mega Prep kits (Qiagen) and transfected into Expi293F cells using ExpiFectamine 293 reagents (Thermo Fisher) ^59^. Antibody was purified from harvested cell culture supernatants using Sure70 protein A resin and eluted with glacial acetic acid. The pH of the antibody solution was neutralized with Tris-HCl pH8. Purified recombinant antibodies were buffer exchanged into 25mM Citric Acid/125mM NaCl pH6, aliquoted and stored at −80°C. Protein integrity and expression were verified by SDS-PAGE and Western blot analysis. Alternatively, selected monoclonal antibodies and Fab fragments were generated through GenScript recombinant antibody production services. Recombinant antibodies or Fabs were expressed in CHO cells, purified, and subjected to quality-control analyses, including CE-SDS and/or SDS-PAGE to assess purity and integrity, and SEC-HPLC to evaluate monodispersity and detect aggregates.

### F(ab) Preparation from Purified IgG Samples

F(ab) fragments were generated from purified serum IgG or recombinant mAbs using a modified papain digestion protocol. Purified IgGs were dialyzed into 20 mM sodium phosphate, 10 mM EDTA (pH 7.0) and concentrated to 20 mg mL^-1^ using Amicon Ultra 0.5 mL centrifugal filters (10 kDa MWCO; MilliporeSigma, Cat. #UFC501096). IgGs were digested with papain-agarose resin (Thermo Fisher Scientific) in digestion buffer (20 mM sodium phosphate, 10 mM EDTA, 20 mM cysteine, pH 7.4) for 5 h at 37°C. For selecting mAbs, digestion was performed using Lys-C in place of papain-agarose.

Following digestion, F(ab) fragments were separated from undigested IgG and Fc fragments by incubation with rProtein A Sepharose Fast Flow (Cytiva) for 1 h. Cysteine was removed by centrifugal filtration and buffer exchange into 1x PBS (pH 7.4). F(ab) quality was assessed by SDS-PAGE and size-exclusion chromatography (SEC). For SDS-PAGE, 2 µg protein per lane was resolved on 4-15% TGX stain-free gels (Bio-Rad) under reducing and non-reducing conditions at 200 V in Tris-glycine-SDS running buffer. Bands were visualized using a Gel Doc EZ imager (Bio-Rad) and compared to a protein ladder (Bio-Rad). For SEC, 15 µg protein was injected onto a Superdex 200 Increase 10/300 column (Cytiva) using a 100-µL loop and run at 0.5 mL min^-1^ on an ÄKTA Pure system (Cytiva). Chromatograms were analyzed in Unicorn v7.6.0, and apparent molecular weight was estimated by linear regression using a mixture of protein standards (Cytiva) run on the same column.

### Protein Expression and Purification

Stabilized HIV-1 Env soluble trimers were designed as gp41-chimeric SOSIP gp140s ^77–79^, and further stabilized by the addition of E64K and A316W mutations ^77^. CH505 gp140 Env trimers were expressed as previously described ^80^. In brief, Freestyle293 cells (Invitrogen) were diluted to 1.25×10^6^ cells/mL with fresh Freestyle293 media up to 1L total volume. Cell viability was typically >95% on the day of transfection. The plasmids were equilibrated with Optimem for 5 min at room temperature. 293Fectin was equilibrated with Optimem for 5 min at room temperature. The equilibrated DNA and 293Fectin were complexed together for 20 min at room temperature and added to cells. More specifically, the 1L of cells was co-transfected with 650 μg of SOSIP expressing plasmid DNA and 150 μg of furin expressing plasmid DNA complexed with 293Fectin (Invitrogen). The cell culture supernatant was clarified on day 6 by centrifugation at 4000g for 60 min and filtered through a 0.8 μm filter. Cell culture supernatant was concentrated with a VivaFlow200 (Sartorius) approximately 10 fold, and subjected to positive PGT145 antibody affinity chromatography. PGT145 affinity column were prepared by conjugating it to Sepharose Fast Flow (Millipore Sigma) resin via CnBr chemistry per the manufacturer’s protocol at a 10 mg of PGT145 per mL of Sepharose ratio. PGT145 resin was packed into an empty 10/10 column (Cytvia) in PBS/0.02% azide. Concentrated cell culture supernatant was loaded onto the column using an AKTA Pure (Cytvia) and eluted with 5 column volumes of 3M MgCl_2_. Eluted protein was concentrated down to 2 mL for size exclusion chromatography with a Superose6 16/600 column (GE Healthcare) at 1 mL/min in 10 mM Tris pH8, 500 mM NaCl. Fractions containing trimeric HIV-1 Env protein were pooled together, sterile-filtered, snap frozen, and stored at −80°C.

### Antibody Blocking ELISA

Antibody blocking assays were performed in 384-well plates (Costar #3700). Monoclonal antibodies with defined SOSIP binding specificities were conjugated to horseradish peroxidase (HRP) using Lightning-Link® labeling kits (Novus Biologicals #701-0003). Plates were coated overnight at 4°C with streptavidin (Thermo Fisher Scientific #S-888) at 2 µg/ml in 0.1 M sodium bicarbonate buffer, then washed with PBS/0.1% Tween-20 and blocked for 60 minutes with assay diluent (PBS containing 4% (w/v) whey protein, 15% normal goat serum, 0.5% Tween-20, and 0.05% sodium azide). Biotinylated SOSIPs were added at concentrations empirically determined to permit optimal blocking by their cognate unlabeled antibody, as established in prior optimization experiments, and incubated for 60 minutes (10 µl/well). After washing, unlabeled purified human monoclonal antibodies were added in 10 µl/well across a three-fold serial dilution starting at 50 µg/ml and incubated for 60 minutes. Plates were washed and HRP-conjugated antibodies were added at 10 µl/well for 60 minutes at concentrations approximating their EC50, as determined during optimization. Following a final wash, HRP activity was detected by addition of 20 µl SureBlue Reserve TMB substrate (Seracare #5120-0081), and the reaction was stopped with 20 µl HCl stop solution. Absorbance was measured at 450 nm. Percent blocking was calculated as: 100 − [(sample OD / 0% blocking control OD) × 100]. All steps were carried out at room temperature unless otherwise indicated.

### Negative Stain Electron Microscopy (NSEM)

IgG and CH505 TF SOSIP trimer were mixed at a 1.5:1 ratio (IgG:trimer), diluted to 400 µg/ml SOSIP with HBSO_4_-150 buffer (150 mM Na_2_SO_4_, 2.5 mM NaCl, 2.8 Na3N, 20 mM HEPES, pH 7.4), and incubated 1 hr at RT. Then a portion of the mixture was diluted 1:4 in HBSO_4_-150 buffer containing 16 mM glutaraldehyde, fixed 5 min, quenched by adding 1 M Tris (pH 7.4) to give 80 mM final Tris concentration, and incubated for 5 min. Quenched samples were diluted with HBSO_4_-150 to **100** µg/ml with HBSO_4_-150 buffer and applied to a glow-discharged carbon-coated EM grid for 10-12 second, blotted and stained with 2 g/dL uranyl formate for 1 min, then blotted and air-dried. Grids were examined on a Philips EM420 electron microscope operating at 120 kV and nominal magnification of 49,000x, and ∼100 images for each complex were collected on a 76 Mpix CCD camera at 2.4 Å/pixel. Images were analyzed by 2D class averages and 3D reconstructions calculated using standard protocols with Relion 3.0 ^81^.

### Cryo-Electron Microscopy (cryo-EM)

For grid preparation, Env trimer (1.0-2.0 mg mL^-1^) was mixed with a 5-fold molar excess of Fab and incubated at room temperature for 30 min to 1 hour. A 3.0 to 3.5-µL aliquot of the complex was applied to glow-discharged Quantifoil Cu R1.2/1.3 grids (Electron Microscopy Sciences) that had been treated for 15 to 40 s using a PELCO easiGlow system (Ted Pella). Grids were incubated for 30 s at >95% relative humidity, blotted for 2.5 s, and plunge-frozen in liquid ethane. Data were collected on a 300 keV Titan Krios (Thermo Fisher Scientific) equipped with a K3 direct electron detector (Gatan) using a nominal defocus range of between −3.0 and −0.5 µm and a total electron exposure ranging from 51.5-66.0 e⁻ Å^-2^. Movies were processed in cryoSPARC ^82^. Beam-induced motion was corrected using Patch Motion Correction, and CTF parameters were estimated for each micrograph. Particles were autopicked using a blob-based picker and extracted with a 320-pixel extraction box size Fourier cropped to 80 pixels for initial cleanup. After iterative 2D classification to remove junk, selected particles were re-extracted with a 320-pixel box and subjected to heterogeneous refinement followed by non-uniform refinement to generate the final reconstruction(s). For processing DH1595.2/DH1786.1 Fab-bound CH505 TF SOSIP Env complex datasets, cryo-EM maps obtained from processing datasets collected via Tundra Cryo-TEM (100 keV; Thermo Fisher Scientific) were used as volume inputs for heterogeneous refinement. Model fitting and refinement were performed using UCSF Chimera/ChimeraX ^83^, Coot ^84^, and Phenix ^85^. Figures were prepared in ChimeraX ^83^ and PyMOL ^86^.

### Surface Plasmon Resonance (SPR)

SPR experiments were performed on a T-200 Biacore System (Cytiva) operating at 25 °C. HBS-EP+ (10 mM HEPES, pH 74, 150 mM NaCl, 3 mM EDTA and 0.05% surfactant P-20) was used as running buffer.

For assessing the binding of DH1777 lineage Fab, either PGT145 or VRC34.01 IgG was injected at 200 nM (120 s at 5 µL/min) on an anti-human Fc antibody surface prepared on a Series S CM5 chip (Cytiva) using a Human Antibody Capture Kit (Cytiva). This was followed by the injection of 200 nM CH505 TF SOSIP trimer (120 s at 5 µL/s). 250 nM Fab was then injected (240 s at 50 µL/s, followed by a 360s off phase) over the PGT145/VRC34.01-bound CH505 TF SOSIP trimer. The surface was regenerated with three successive 3M MgCl_2_ injections, each lasting 10 seconds at 100 µL/min.

For assessing the binding of DH1530.3 mutant constructs, PGT145 IgG was injected at 200 nM (120 s at 10 µL/min) on an anti-human Fc antibody surface prepared on a Series S CM5 chip using a Human Antibody Capture Kit. This was followed by injection of 200 nM CH505 TF SOSIP trimer (120 s at 510µL/s). 500 nM Fab was then injected (120 s at 50 µL/s, followed by a 360s off phase) over the PGT145-bound CH505 TF SOSIP trimer.

The surface was regenerated with three successive 3M MgCl_2_ injections, each lasting 10 seconds at 100 µL/min.

### Reagent Authentication

Expi293F and TZM-bl cell lines were provided with a certificate of analysis from their sources. Cell identity is verified by morphology or fluorescent markers expressed. Positive control antibodies were used in the TZM-bl neutralization assay to distinguish the cell lines and for comparison between lots when new vials of cells are cultured.

### Quantification and statistical analysis

Frequencies of autologous neutralizing lineages within each phenotypic category were calculated as the quotient of positive events over the total number of antigen-specific Bmem cells (n) assayed. To identify vaccine responders, we implemented a hierarchical statistical framework in which each participant’s frequency was compared against the lowest-responder reference using pairwise two-sided Fisher’s exact tests. This analysis was performed across multiple nested epitope categories: (i) total autologous tier 2 TF neutralizers, (ii) V3-directed neutralizers, (iii) any CD4bs-directed neutralizers, and (iv) S365P-sensitive, CDRH3-dominant-type CD4bs neutralizers. For the repertoire-level analysis, frequencies were additionally normalized to the total Bmem cell compartment.

Raw p-values were adjusted for multiple hypothesis testing using the Benjamini–Hochberg procedure to control the false discovery rate (FDR) at 0.05; this correction was selected to maintain statistical validity given the hierarchical positive regression dependency of the epitope mapping data (e.g., S365P-sensitivity nested within total CD4bs reactivity). Statistical significance was defined as *P_adj_* < 0.05, with marginal significance explicitly noted where relevant. Fold-enrichment for elite responders (e.g., UID 123) was calculated relative to the cohort geometric mean. The hierarchical distribution of antigen-specific B cell–derived antibodies—from total recovered clones through CH505-binding, virus-neutralizing, and autologous TF-neutralizing subsets—was visualized as a Sankey diagram using SankeyMatic (https://sankeymatic.com/).

To identify participants enriched for autologous tier 2 neutralizing activity, we compared neutralizing Bmem cell frequencies across participants using the hierarchical framework described above. Five participants showed significant enrichment of autologous tier 2 neutralizing CH505-positive Bmem cells: UIDs 123, 469, 185, 332, and 283 (**Fig. 6A**; *P_adj_* < 0.05). However, epitope-level analysis revealed that these responses differed in specificity. UID 332 exhibited the strongest enrichment for total TF neutralizers (*P_adj_* = 4.5 × 10^−17^) and V3-targeting neutralizers (*P_adj_* = 4.2 × 10^−31^), whereas enrichment for CD4bs-directed, CDRH3-dominant activity was much weaker and only marginal after correction (**Fig. 6A**; *P_adj_* = 0.0509). UID 283 showed a closely similar pattern, with significant enrichment for total TF neutralizers and V3-targeting neutralizers, but complete absence of detectable CD4bs-directed neutralization. Thus, whereas UID 332 may contain a minor subdominant CD4bs component, both participants were dominated by strain-specific V3-directed responses. We therefore refined the analysis to focus on high-confidence CD4bs-directed responses by examining the frequencies of (i) any CD4bs neutralization and (ii) S365P-sensitive neutralization, a signature of CDRH3-dominant-type CD4bs antibodies. By this more stringent definition, UIDs 123, 469, and 185 emerged as the principal CD4bs responder group, retaining strong statistical significance in both categories (**Fig. 6A**). We next anchored these findings at the repertoire level by quantifying autologous tier 2 neutralizer frequencies across the total Bmem compartment. Under this normalized framework, UID 123 remained significant (*P_adj_* = 0.0052) and exhibited a distinct quantity-plus-quality profile, with an approximately 5-fold enrichment of S365P-sensitive, CDRH3-dominant-type CD4bs precursors relative to the cohort mean (**Fig. 6A**). UID 123 also stood out qualitatively, as it was the only participant to yield the FPPR-targeting neutralizing lineage DH1708 (**Figs. 1F and 4D**). The convergence of a significantly enriched CDRH3-dominant CD4bs precursor reservoir and the unique recovery of both V3- and FPPR-targeting neutralizing lineages nominated UID 123 for deeper investigation.

## Notes

### Competing Interest Statement

Conflicts of Interest: SRW has received institutional grants or contracts from Sanofi Pasteur, Janssen Vaccines/Johnson & Johnson, Moderna Tx, Hookipa, Pfizer, AbbVie, Vir Biotechnology, and Worcester HIV Vaccine; has participated on data safety monitoring or advisory boards for Janssen Vaccines/Johnson & Johnson, Cyanvac, CSL Behring, and BioNTech; and his spouse holds stock/stock options in Regeneron Pharmaceuticals. EBW has received research funding from Pfizer, Moderna, Seqirus, Najit Technologies, and Clinetic for the conduct of clinical research studies. He has also received support as an advisor to Vaxcyte, Seqirus, and Pfizer, consultant to ILiAD Biotechnologies, and DSMB member for Shionogi and Emmes. BFH and KOS have patents on the CH505 Env immunogen in this study. Disclaimer: This article was written by some authors [LP] in their capacity as employees of the National Institutes of Health (NIH), but the views expressed herein do not necessarily represent those of the NIH.

### Clinical Trial

NCT04915768

### Author Declarations

The Partners (MassGeneral Brigham) Human Research Committee gave ethical approval for this work.

### Summary of Updates

In this version, we have reorganized the manuscript structure to improve logical flow and readability, while ensuring all structural data and coordinates have been fully deposited into the appropriate repository and cited in the Data Availability statement.

