## Supplementary material for "Priming of Multiple HIV Neutralizing B Cell Precursors in Humans": Figure S1-S9

**A**

| Participant UID | Vaccination | | | | |
|---|---|---|---|---|---|
| | 1st | 2nd | 3rd | 4th | 5th |
| 67 | v | v | v | v | v |
| 86 | v | v | v | v | v |
| 123 | v | v | v | v | v |
| 185 | v | v | v | v | |
| 192 | v | v | v | v | v |
| 283 | v | v | v | v | v |
| 332 | v | v | v | v | |
| 440 | v | v | | | |
| 468 | v | | | | |
| 469 | v | v | v | v | v |
| 475 | v | v | v | v | v |
| n = | 11 | 10 | 9 | 9 | 7 |

**B**

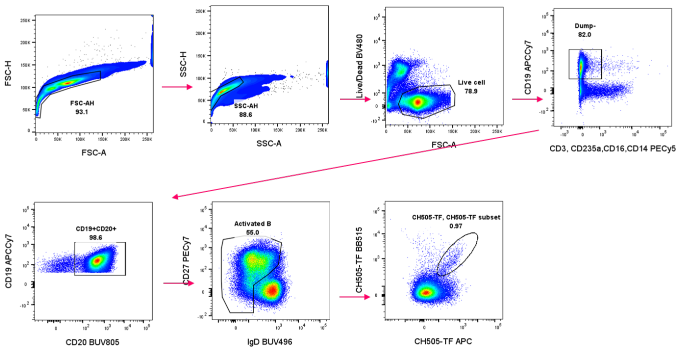


**C**

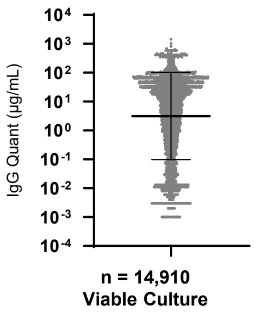


**D**

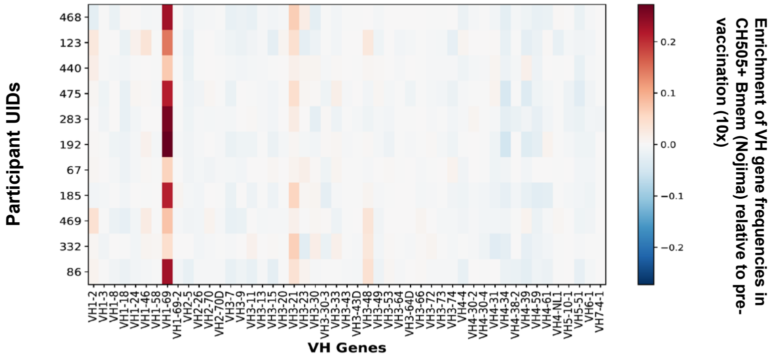


**E**

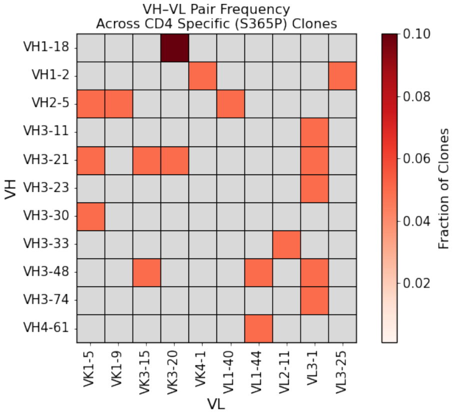


**F**

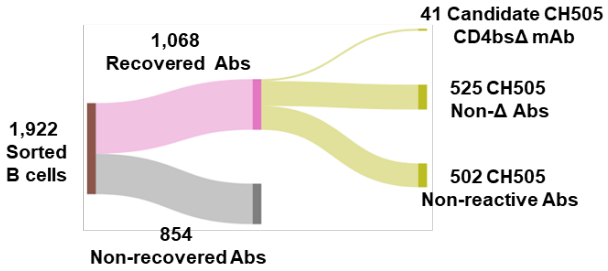


**G**

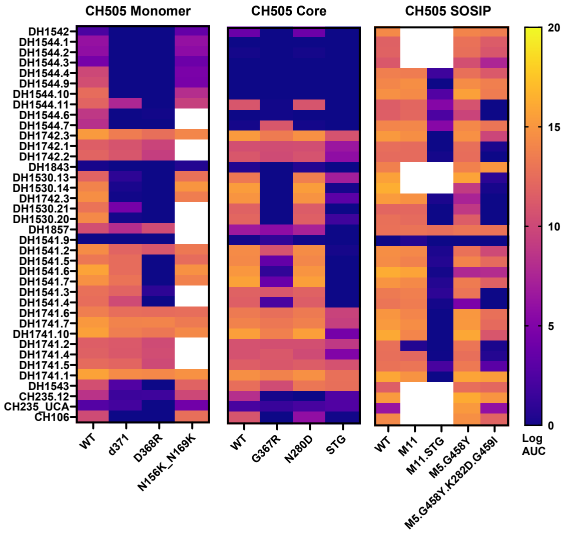


**H**

| HIV-1 Neutralization IC50 (ug/mL) | | | | | |
|---|---|---|---|---|---|
| Antibody ID | SVA-MLV | CH505TF | CH505.w4.3 | CH505TF.gly4 | CH505TF.gly4.S365P |
| DH1544.10 | >50 | 20 | 0.62 | 0.03 | 0.04 |
| DH1544.6 | >50 | 47 | 0.64 | 0.04 | 0.12 |
| DH1530.13 | >50 | 0.21 | 0.02 | 0.001 | 13 |
| DH1530.14 | >50 | 2.7 | 0.02 | 0.01 | >50 |
| DH1530.22 | >50 | 7.5 | 0.03 | 0.02 | >50 |
| DH1530.21 | >50 | 0.46 | 0.05 | 0.001 | >50 |
| DH1530.20 | >50 | 1 | 0.06 | 0.003 | >50 |
| DH1541.5 | >50 | 17 | 0.15 | 0.02 | >50 |
| DH1541.7 | >50 | 10 | 0.13 | 0.01 | >50 |
| DH1541.3 | >50 | 0.59 | 0.04 | 0.001 | >50 |
| DH1741.5 | >50 | 36 | 0.03 | 7.4 | 2.6 |
| CH103_4.A | >50 | 1.3 | 0.1 | 0.004 | 0.01 |
| CH235UCA_tkLL_v3_4A/29 | >50 | >50 | >50 | >50 | >50 |
| DH235.12 | >50 | 0.12 | 0.08 | 0.001 | 0.01 |

| Neutralization Titer (IC50) µg/mL | <0.1 | 0.1-1.0 | 2.0-5.0 | 6.0-10.0 | 11.0-50.0 | >50 |
|---|---|---|---|---|---|---|

**Figure S1.**

**Figure S1. Immunization completion and immunogenetic and functional characterization of CH505 TF–specific memory B-cell responses in HVTN 300.**

**(A) Immunization completion by trial participant.** Individual scheduled vaccination visits are shown for each participant, with "v" indicating that the immunization was received and blank cells indicating that it was not received. The number of participants who completed each total number of immunizations is summarized.

**(B) Representative FACS gating strategy for the isolation of CH505 TF double-positive (DP) antigen-specific memory B cells from post-immunization leukapheresis samples.** Sequential gating excluded dead cells and non-B-cell lineages, identified the memory B-cell compartment, and selected cells that bound both VB515- and AF647-labeled CH505 TF SOSIP probes.

**(C) Distribution of IgG concentrations in culture supernatants from 14,190 viable Nojima single-cell cultures.** The geometric mean concentration was 3.14 μg/mL, with a geometric standard deviation factor of 32.3. The median concentration was 9.37 μg/mL, with an interquartile range of 0.30–47.0 μg/mL. The summary bar indicates the geometric mean ± geometric standard deviation.

**(D) Enrichment of heavy-chain V-gene usage within the vaccine-elicited CH505 TF–specific memory B-cell repertoire.** The heatmap shows participant-level differences in VH-gene usage among CH505 TF DP memory B cells collected at week 34 relative to each participant's matched pre-vaccination baseline repertoire, as determined by 10x BCR sequencing. Rows represent individual HVTN 300 Part A participants, and columns represent VH genes. Colors indicate the direction and magnitude of differential representation, with red indicating enrichment and blue indicating depletion relative to baseline.

**(E) Paired VH- and VL-gene usage among S365P-sensitive CH505 TF-neutralizing clones.** The bipartite plot shows heavy- and light-chain V-gene pairings among S365P-sensitive autologous CH505 TF-neutralizing clones recovered by Nojima culture. Each connection represents a paired VH–VL assignment from an individual neutralizing clone, with repeated connections indicating recurrent use of the same immunogenetic pairing. The broad distribution of VH–VL pairings indicates that S365P-sensitive CH505 TF neutralization arose from diverse paired BCR genotypes rather than being restricted to a single heavy- and light-chain combination.

**(F) Sankey diagram summarizing single-cell RT-PCR amplification and paired VH/VL recovery from conventionally sorted CD4-binding-site (CD4bs)-reactive B cells from four participants.**

**(G) Heatmap of ELISA area-under-the-curve (AUC) values showing the CH505 CD4bs-binding profiles of 41 candidate CD4bs-reactive monoclonal antibodies recovered by conventional single-cell RT-PCR.**

**(H) Neutralization profiles of 11 candidate CD4bs-reactive monoclonal antibodies recovered by conventional single-cell RT-PCR that mediated autologous tier-2 neutralization of CH505 TF.**

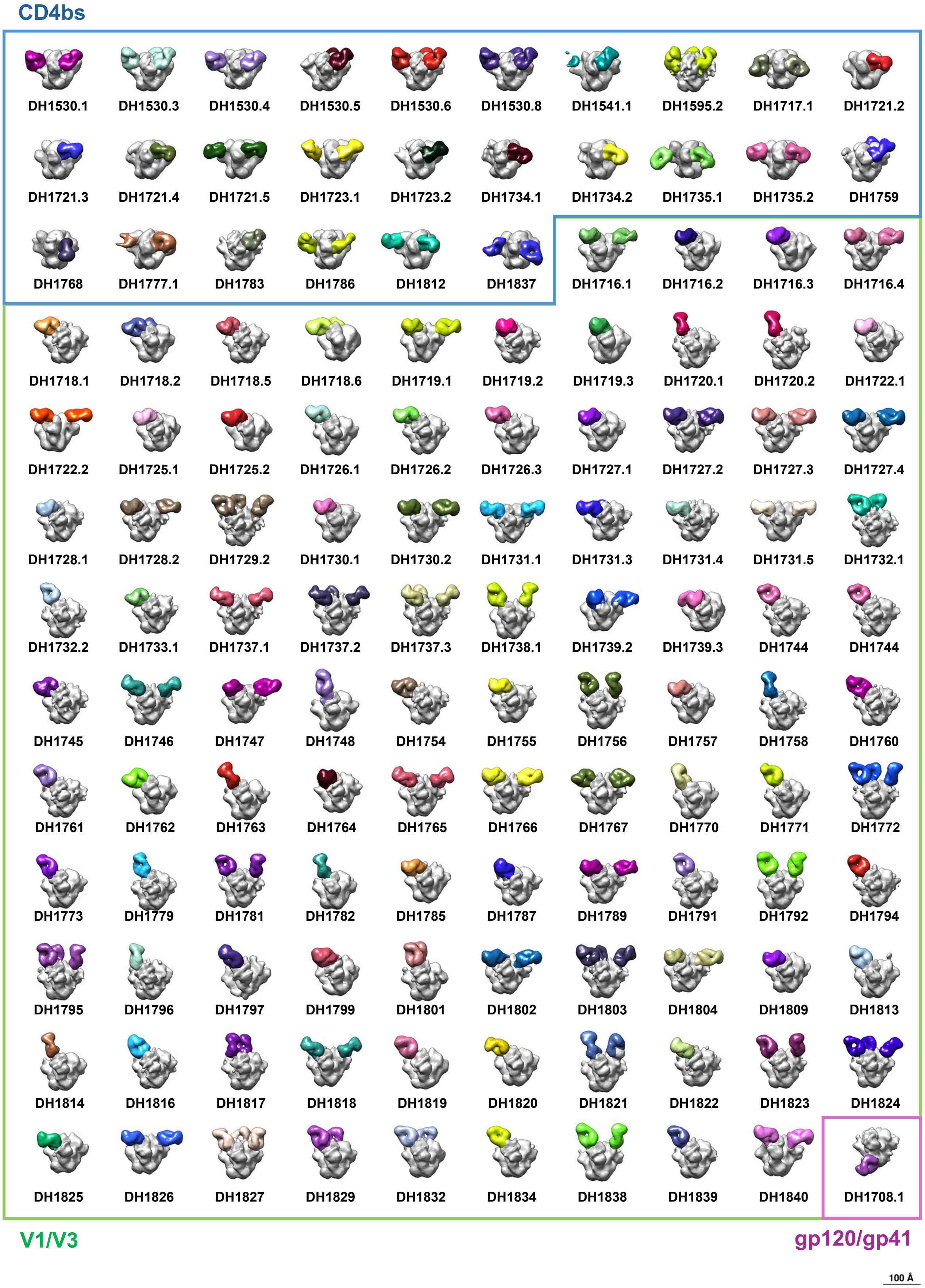
CD4bs
DH1530.1
DH1530.3
DH1530.4
DH1530.5
DH1530.6
DH1530.8
DH1541.1
DH1595.2
DH1717.1
DH1721.2
DH1721.3
DH1721.4
DH1721.5
DH1723.1
DH1723.2
DH1734.1
DH1734.2
DH1735.1
DH1735.2
DH1759
DH1768
DH1777.1
DH1783
DH1786
DH1812
DH1837
DH1716.1
DH1716.2
DH1716.3
DH1716.4
DH1718.1
DH1718.2
DH1718.5
DH1718.6
DH1719.1
DH1719.2
DH1719.3
DH1720.1
DH1720.2
DH1722.1
DH1722.2
DH1725.1
DH1725.2
DH1726.1
DH1726.2
DH1726.3
DH1727.1
DH1727.2
DH1727.3
DH1727.4
DH1728.1
DH1728.2
DH1729.2
DH1730.1
DH1730.2
DH1731.1
DH1731.3
DH1731.4
DH1731.5
DH1732.1
DH1732.2
DH1733.1
DH1737.1
DH1737.2
DH1737.3
DH1738.1
DH1739.2
DH1739.3
DH1744
DH1744
DH1745
DH1746
DH1747
DH1748
DH1754
DH1755
DH1756
DH1757
DH1758
DH1760
DH1761
DH1762
DH1763
DH1764
DH1765
DH1766
DH1767
DH1770
DH1771
DH1772
DH1773
DH1779
DH1781
DH1782
DH1785
DH1787
DH1789
DH1791
DH1792
DH1794
DH1795
DH1796
DH1797
DH1799
DH1801
DH1802
DH1803
DH1804
DH1809
DH1813
DH1814
DH1816
DH1817
DH1818
DH1819
DH1820
DH1821
DH1822
DH1823
DH1824
DH1825
DH1826
DH1827
DH1829
DH1832
DH1834
DH1838
DH1839
DH1840
DH1708.1
V1/V3
gp120/gp41
100 Å

**Figure S2.**

**Figure S2. Negative-stain electron microscopy of CH505 TF–mAb complexes.** NSEM images are shown for 130 of the 131 CH505 TF–mAb complexes analyzed. Env trimers are shown in gray, and bound Fabs are shown in color. Colored borders denote three epitope classes: CD4bs-directed (blue), V1V3-directed (green), and gp120/gp41-directed (purple). DH1749 is not shown here and is presented in **Fig. 4A**. Scale bar, 10 nm.

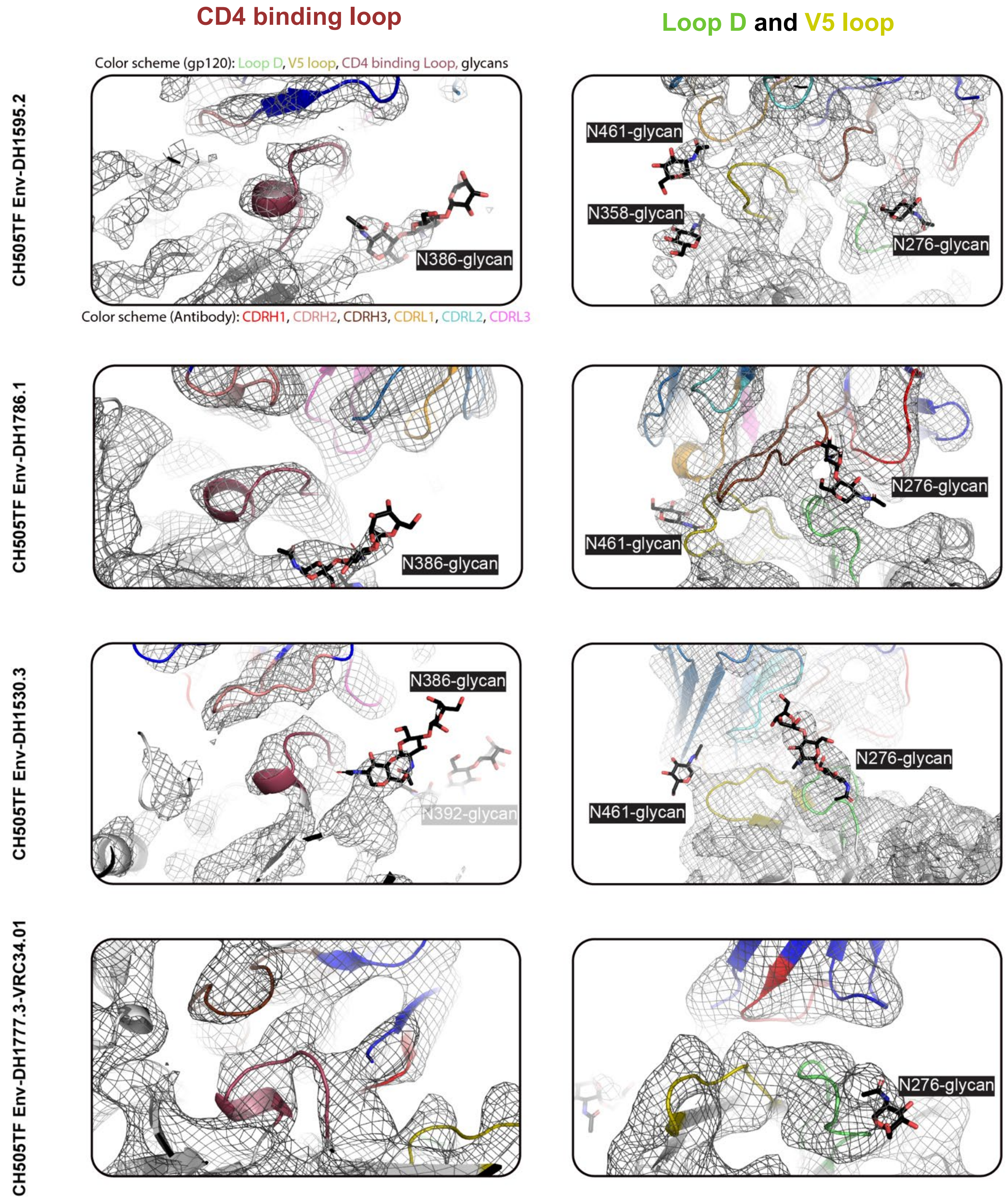


**Figure S3. Cryo-EM coordinates fitting for vaccine-elicited CD4bs antibodies isolated in the HVTN 300 clinical trial.** Views around the CD4 binding loop, Loop D, and V5 loop of the complexes between CH505 TF Env and CD4bs targeting antibodies are shown. The cryo-EM map is shown as a gray mesh, with the fitted model in cartoon representation and glycans shown as sticks. The CD4 binding loop is colored raspberry, Loop D mint, V5 loop olive, CDRH1 red, CDRH2 salmon, CDRH3 brown, gp41 dark gray, heavy chains blue, light chain sky blue, glycans black.

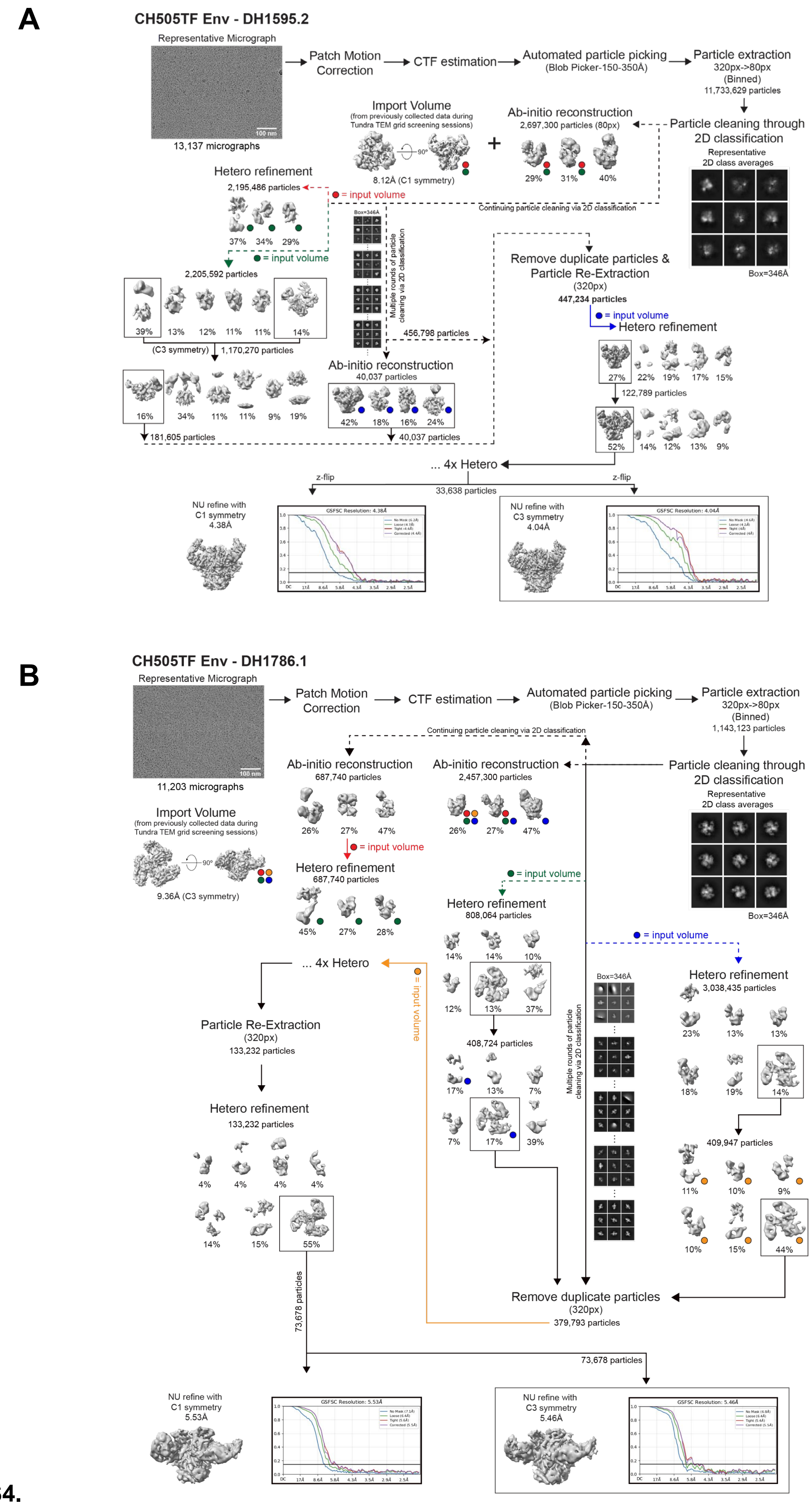
A
CH505TF Env - DH1595.2
Representative Micrograph
100 nm
13,137 micrographs
Patch Motion Correction
CTF estimation
Automated particle picking
(Blob Picker-150-350Å)
Particle extraction
320px->80px
(Binned)
11,733,629 particles
Particle cleaning through 2D classification
Representative 2D class averages
Box=346Å
Import Volume
(from previously collected data during Tundra TEM grid screening sessions)
90°
8.12Å (C1 symmetry)
Ab-initio reconstruction
2,697,300 particles (80px)
29%
31%
40%
Continuing particle cleaning via 2D classification
= input volume
Hetero refinement
2,195,486 particles
37%
34%
29%
= input volume
2,205,592 particles
39%
13%
12%
11%
11%
14%
(C3 symmetry)
1,170,270 particles
16%
34%
11%
11%
9%
19%
181,605 particles
Box=346Å
Multiple rounds of particle cleaning via 2D classification
456,798 particles
Ab-initio reconstruction
40,037 particles
42%
18%
16%
24%
40,037 particles
Remove duplicate particles & Particle Re-Extraction
(320px)
447,234 particles
= input volume
Hetero refinement
27%
22%
19%
17%
15%
122,789 particles
52%
14%
12%
13%
9%
... 4x Hetero
z-flip
33,638 particles
z-flip
NU refine with C1 symmetry
4.38Å
GSFSC Resolution: 4.38Å
NU refine with C3 symmetry
4.04Å
GSFSC Resolution: 4.04Å
B
CH505TF Env - DH1786.1
Representative Micrograph
100 nm
11,203 micrographs
Patch Motion Correction
CTF estimation
Automated particle picking
(Blob Picker-150-350Å)
Particle extraction
320px->80px
(Binned)
1,143,123 particles
Continuing particle cleaning via 2D classification
Ab-initio reconstruction
687,740 particles
26%
27%
47%
Ab-initio reconstruction
2,457,300 particles
26%
27%
47%
Particle cleaning through 2D classification
Representative 2D class averages
Box=346Å
Import Volume
(from previously collected data during Tundra TEM grid screening sessions)
90°
9.36Å (C3 symmetry)
= input volume
Hetero refinement
687,740 particles
45%
27%
28%
= input volume
Hetero refinement
808,064 particles
14%
14%
10%
12%
13%
37%
408,724 particles
17%
13%
7%
7%
17%
39%
... 4x Hetero
= input volume
Particle Re-Extraction
(320px)
133,232 particles
Hetero refinement
133,232 particles
4%
4%
4%
4%
14%
15%
55%
Box=346Å
Multiple rounds of particle cleaning via 2D classification
= input volume
Hetero refinement
3,038,435 particles
23%
13%
13%
18%
19%
14%
409,947 particles
11%
10%
9%
10%
15%
44%
Remove duplicate particles
(320px)
379,793 particles
73,678 particles
73,678 particles
NU refine with C1 symmetry
5.53Å
GSFSC Resolution: 5.53Å
NU refine with C3 symmetry
5.46Å
GSFSC Resolution: 5.46Å

Figure S4.

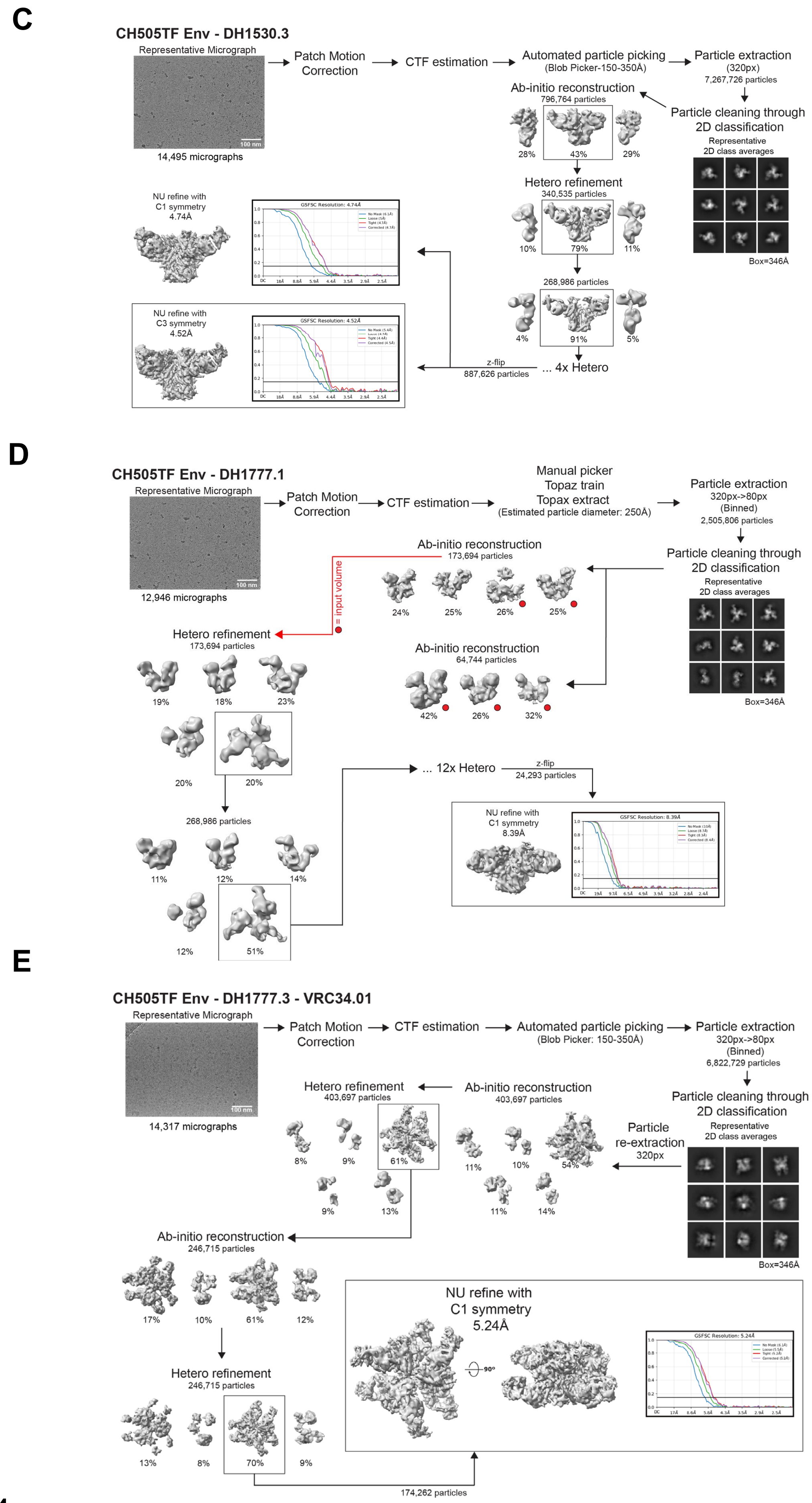

C
CH505TF Env - DH1530.3
Representative Micrograph
100 nm
14,495 micrographs
Patch Motion Correction
CTF estimation
Automated particle picking
(Blob Picker-150-350Å)
Particle extraction
(320px)
7,267,726 particles
Particle cleaning through 2D classification
Representative 2D class averages
Box=346Å
Ab-initio reconstruction
796,764 particles
28%
43%
29%
Hetero refinement
340,535 particles
10%
79%
11%
268,986 particles
4%
91%
5%
... 4x Hetero
z-flip
887,626 particles
NU refine with C1 symmetry 4.74Å
GSFSC Resolution: 4.74Å
NU refine with C3 symmetry 4.52Å
GSFSC Resolution: 4.52Å
D
CH505TF Env - DH1777.1
Representative Micrograph
100 nm
12,946 micrographs
Patch Motion Correction
CTF estimation
Manual picker
Topaz train
Topax extract
(Estimated particle diameter: 250Å)
Particle extraction
320px->80px
(Binned)
2,505,806 particles
Particle cleaning through 2D classification
Representative 2D class averages
Box=346Å
Ab-initio reconstruction
173,694 particles
24%
25%
26%
25%
= input volume
Ab-initio reconstruction
64,744 particles
42%
26%
32%
Hetero refinement
173,694 particles
19%
18%
23%
20%
20%
268,986 particles
11%
12%
14%
12%
51%
... 12x Hetero
z-flip
24,293 particles
NU refine with C1 symmetry 8.39Å
GSFSC Resolution: 8.39Å
E
CH505TF Env - DH1777.3 - VRC34.01
Representative Micrograph
100 nm
14,317 micrographs
Patch Motion Correction
CTF estimation
Automated particle picking
(Blob Picker: 150-350Å)
Particle extraction
320px->80px
(Binned)
6,822,729 particles
Particle cleaning through 2D classification
Representative 2D class averages
Box=346Å
Particle re-extraction
320px
Ab-initio reconstruction
403,697 particles
11%
10%
54%
11%
14%
Hetero refinement
403,697 particles
8%
9%
61%
9%
13%
Ab-initio reconstruction
246,715 particles
17%
10%
61%
12%
Hetero refinement
246,715 particles
13%
8%
70%
9%
174,262 particles
NU refine with C1 symmetry 5.24Å
90°
GSFSC Resolution: 5.24Å


**Figure S4.**

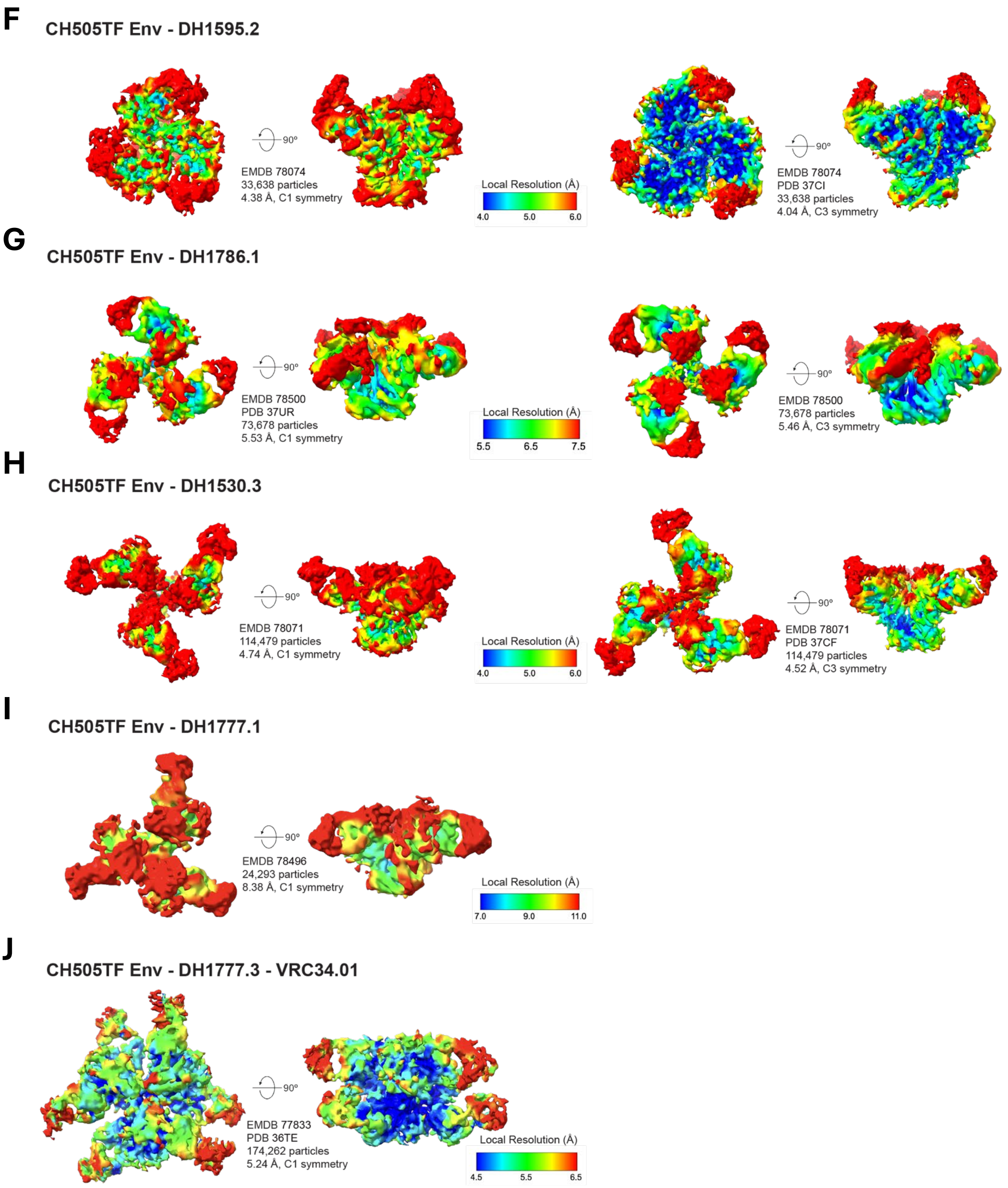
F
CH505TF Env - DH1595.2
90°
EMDB 78074
33,638 particles
4.38 Å, C1 symmetry
Local Resolution (Å)
4.0
5.0
6.0
90°
EMDB 78074
PDB 37CI
33,638 particles
4.04 Å, C3 symmetry
G
CH505TF Env - DH1786.1
90°
EMDB 78500
PDB 37UR
73,678 particles
5.53 Å, C1 symmetry
Local Resolution (Å)
5.5
6.5
7.5
90°
EMDB 78500
73,678 particles
5.46 Å, C3 symmetry
H
CH505TF Env - DH1530.3
90°
EMDB 78071
114,479 particles
4.74 Å, C1 symmetry
Local Resolution (Å)
4.0
5.0
6.0
90°
EMDB 78071
PDB 37CF
114,479 particles
4.52 Å, C3 symmetry
I
CH505TF Env - DH1777.1
90°
EMDB 78496
24,293 particles
8.38 Å, C1 symmetry
Local Resolution (Å)
7.0
9.0
11.0
J
CH505TF Env - DH1777.3 - VRC34.01
90°
EMDB 77833
PDB 36TE
174,262 particles
5.24 Å, C1 symmetry
Local Resolution (Å)
4.5
5.5
6.5

**Figure S4.**

**Figure S4. Cryo-EM data processing workflows and map quality for CH505 TF Env complexes with vaccine-elicited CD4-binding-site antibodies.**

**(A–E) Cryo-EM data processing workflows for CH505 TF Env in complex with (A) DH1595.2, (B) DH1786.1, (C) DH1530.3, (D) DH1777.1, and (E) DH1777.3-VRC34.01.** Each workflow shows a representative cryo-EM micrograph, representative two-dimensional class averages, reconstructed maps generated by ab initio reconstruction and heterogeneous refinement, maps obtained after non-uniform (NU) refinement, and a Fourier shell correlation (FSC) curve from gold-standard refinement. The horizontal blue line indicates the FSC = 0.143 resolution criterion. In **(A)** and **(B)**, volumes used as inputs for heterogeneous refinement are indicated by colored circles, and the corresponding refinement jobs are connected by arrows of the same color. For DH1595.2, an 8.12-Å map of the same complex reconstructed from data collected using a 100-keV Tundra cryo-transmission electron microscope was imported and used as an initial volume. For DH1786.1, a corresponding 9.36-Å Tundra-derived map was imported and used as an initial volume.

**(F–J) Refined cryo-EM maps colored according to local resolution for CH505 TF Env bound to (F) DH1595.2, (G) DH1786.1, (H) DH1530.3, (I) DH1777.1, and (J) DH1777.3-VRC34.01.**

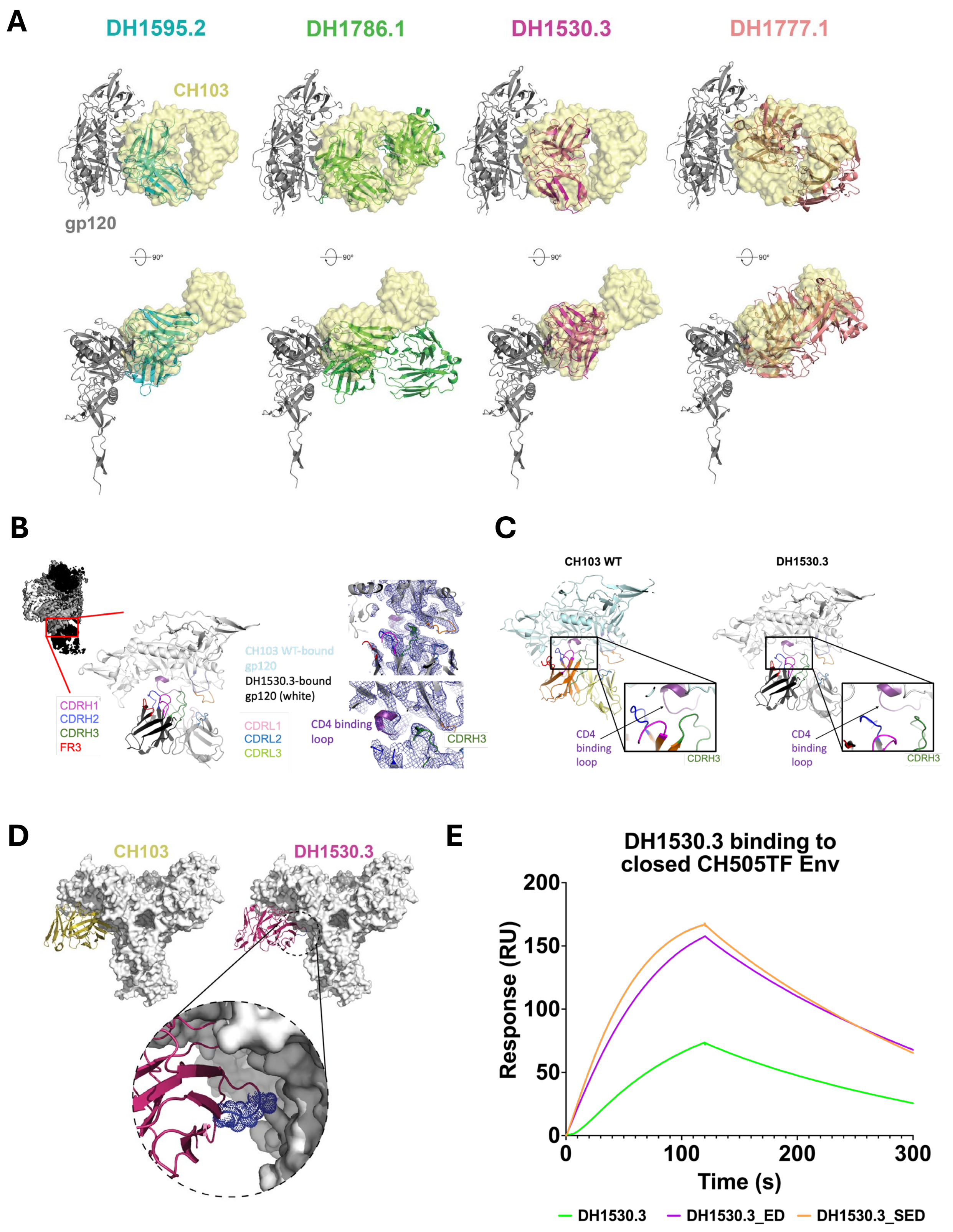
A
DH1595.2
DH1786.1
DH1530.3
DH1777.1
CH103
gp120
90°
B
CDRH1
CDRH2
CDRH3
FR3
CH103 WT-bound gp120
DH1530.3-bound gp120 (white)
CDRL1
CDRL2
CDRL3
CD4 binding loop
CDRH3
C
CH103 WT
DH1530.3
CD4 binding loop
CDRH3
D
CH103
DH1530.3
E
DH1530.3 binding to closed CH505TF Env
Response (RU)
Time (s)
0
50
100
150
200
300
DH1530.3
DH1530.3_ED
DH1530.3_SED

Figure S5.

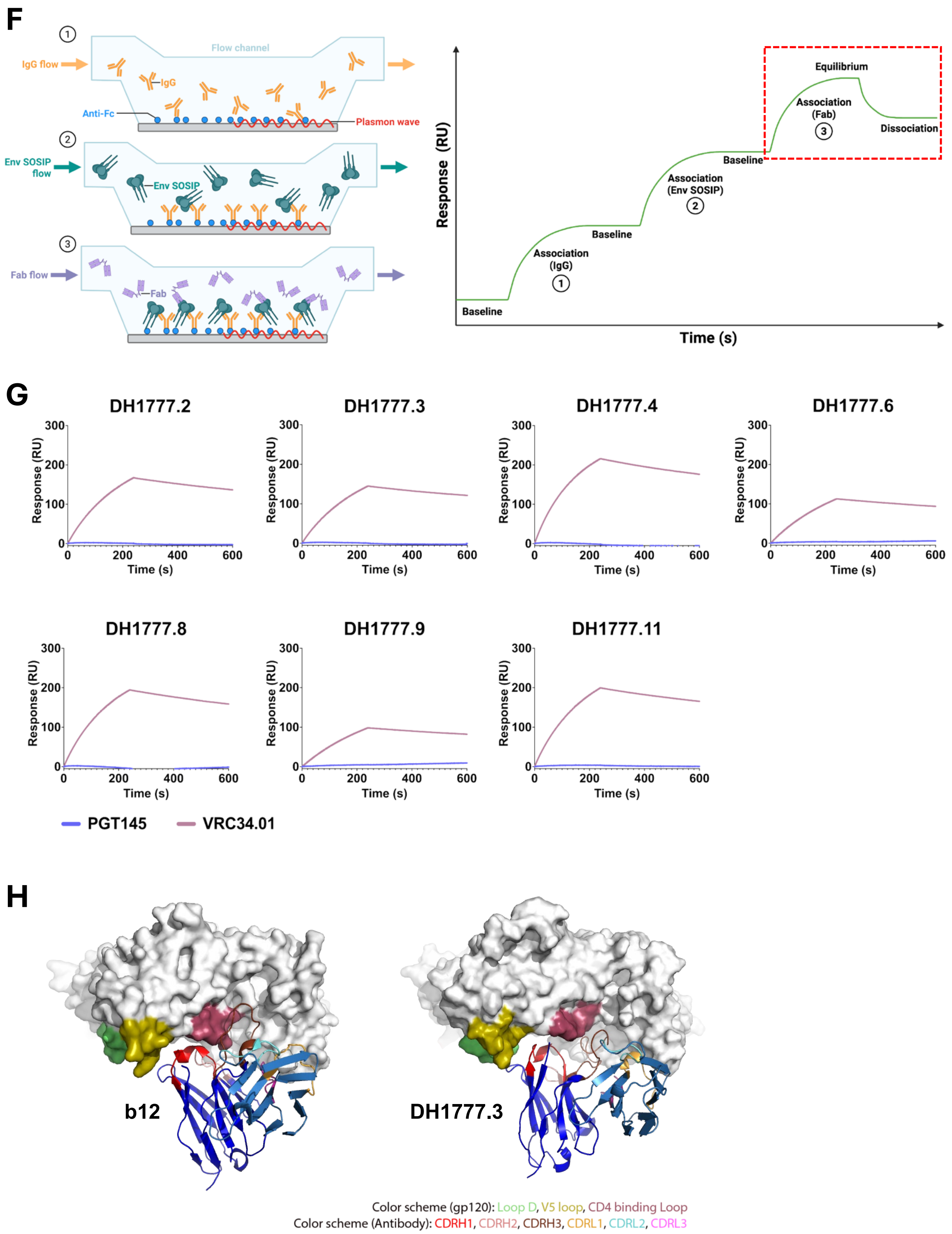
F
IgG flow
Flow channel
IgG
Anti-Fc
Plasmon wave
Env SOSIP flow
Env SOSIP
Fab flow
Fab
Response (RU)
Time (s)
Baseline
Association (IgG)
Association (Env SOSIP)
Association (Fab)
Equilibrium
Dissociation
G
DH1777.2
DH1777.3
DH1777.4
DH1777.6
DH1777.8
DH1777.9
DH1777.11
PGT145
VRC34.01
H
b12
DH1777.3
Color scheme (gp120): Loop D, V5 loop, CD4 binding Loop
Color scheme (Antibody): CDRH1, CDRH2, CDRH3, CDRL1, CDRL2, CDRL3

Figure S5.

**Figure S5. Structural and biophysical characterization of HVTN 300-elicited, CD4bs-targeting antibodies**.

**(A) Comparison of antibody angles of approach.** Two 90° rotated views are shown for a single CH505 TF gp120 protomer bound to the Fabs of DH1595.2, DH1786.1, DH1530.3, and DH1777.1. For each comparison, the CH103-bound gp120 structure was superimposed using the gp120 chains, and the CH103 Fab is shown as a transparent yellow surface.

**(B) Structural details of the DH1530.3-gp120 interface. (Left)** Zoomed-in views of the binding interface between CH505 TF gp120 and DH1530.3 are shown. **(Right)** Zoomed-in views of the antibody interactive region comparing DH1530.3 with CH103. The cryo-EM density is shown as a blue mesh, with the fitted atomic model shown in cartoon representation. CH505 TF gp120 is colored white, CH103-bound gp120 is pale cyan, and the CD4-binding loop is purple. CDRH1, CDRH2, CDRH3, CDRL1, CDRL2, CDRL3, and FR3 are colored magenta, blue, green, pink, sky blue, lime, and red, respectively.

**(C) CDRH3-dominated binding mode of DH1530.3.** Zoomed-in views of the antibody interactive region comparing DH1530.3 with mature CH103 bnAb.

**(D) Structure-guided engineering of DH1530.3 to mimic CH103-like quaternary interactions.** DH1530.3 variants containing either the G75E/N76D substitutions or the A74S/G75E/N76D substitutions were designed to introduce potential contacts with the adjacent Env protomer. The substituted residues are shown as blue spheres in the enlarged view.

**(E) Binding of DH1530.3 wild-type and engineered constructs to CH505TF Env SOSIP restrained in the closed conformation.** CH505 TF SOSIP was captured through PGT145, and each Fab was flowed over the captured Env surface.

**(F) Schematic of the surface plasmon resonance experimental design.** PGT145 or VRC34.01 IgG was captured on an anti-human Fc surface, followed by capture of CH505 TF SOSIP. Fab was flowed over to measure binding to Env. The area boxed in red, dotted lines represent the main association/dissociation steps shown in the SPR sensorgrams of panel G.

**(G) Binding of CD4bs-targeting Fabs to CH505 TF SOSIP captured in distinct conformational states.** Each Fab was flowed over a CH505 TF Env surface captured by either PGT145, which preferentially recognizes closed trimers (blue line) or VRC34.01, which captures Env through the fusion peptide region (ruby line).

**(H) Comparison of the DH1777.3 and b12 binding modes.** The structure of CH505 disulfide-stapled SOSIP bound to b12 Fab (PDB 8SXI) was aligned with CH505 TF SOSIP bound to DH1777.3 using the gp120 chains. The CD4-binding loop is colored raspberry, loop D mint, the V5 loop olive, CDRH1 red, CDRH2 salmon, CDRH3 brown, gp41 dark gray, antibody heavy chains blue, and antibody light chains sky blue.

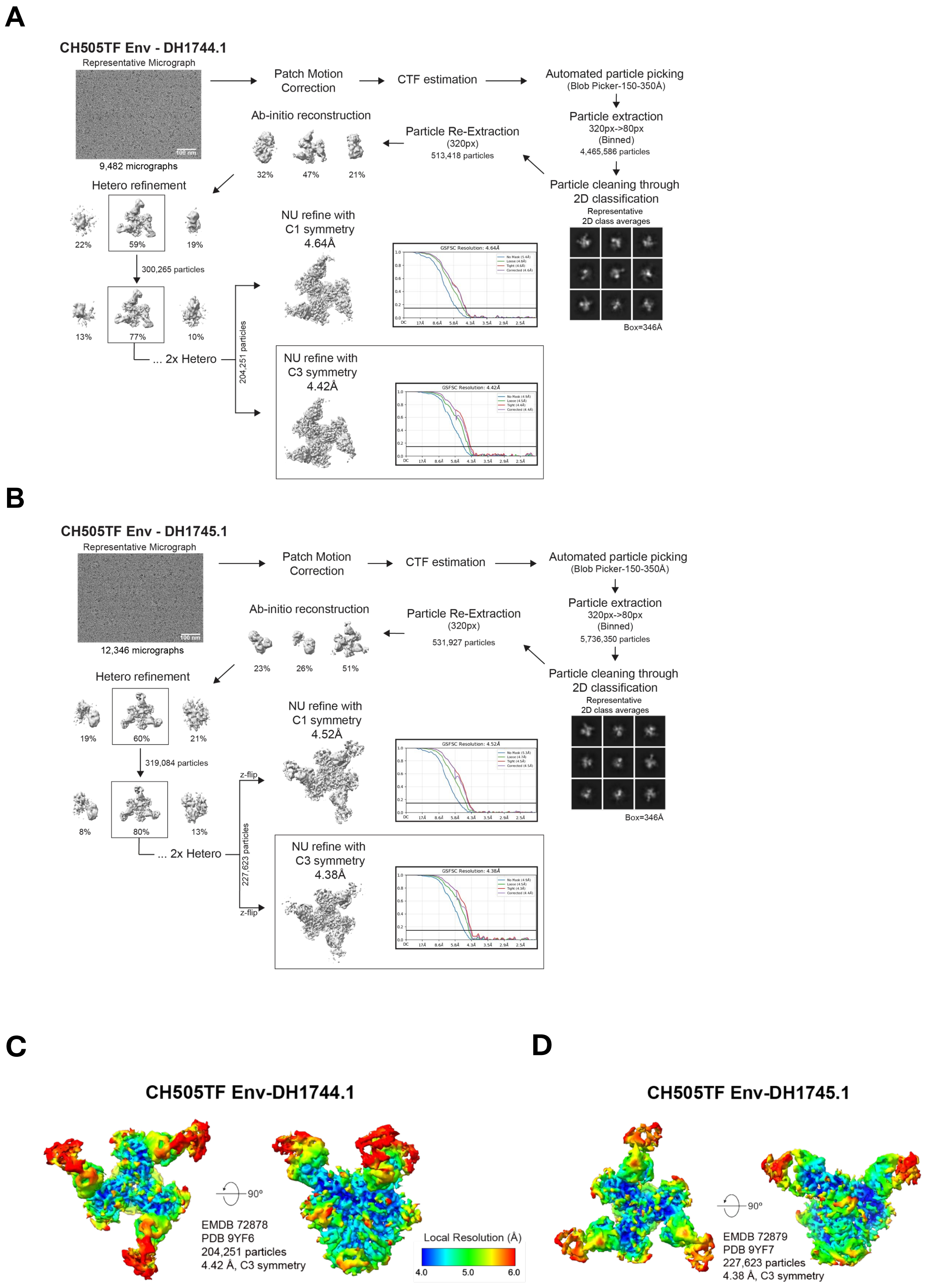
A
CH505TF Env - DH1744.1
Representative Micrograph
100 nm
9,482 micrographs
Patch Motion Correction
CTF estimation
Automated particle picking
(Blob Picker-150-350Å)
Particle extraction
320px->80px
(Binned)
4,465,586 particles
Particle cleaning through 2D classification
Representative 2D class averages
Box=346Å
Particle Re-Extraction
(320px)
513,418 particles
Ab-initio reconstruction
32%
47%
21%
Hetero refinement
22%
59%
19%
300,265 particles
13%
77%
10%
... 2x Hetero
204,251 particles
NU refine with C1 symmetry 4.64Å
GSFSC Resolution: 4.64Å
NU refine with C3 symmetry 4.42Å
GSFSC Resolution: 4.42Å
B
CH505TF Env - DH1745.1
Representative Micrograph
100 nm
12,346 micrographs
Patch Motion Correction
CTF estimation
Automated particle picking
(Blob Picker-150-350Å)
Particle extraction
320px->80px
(Binned)
5,736,350 particles
Particle cleaning through 2D classification
Representative 2D class averages
Box=346Å
Particle Re-Extraction
(320px)
531,927 particles
Ab-initio reconstruction
23%
26%
51%
Hetero refinement
19%
60%
21%
319,084 particles
8%
80%
13%
... 2x Hetero
227,623 particles
z-flip
z-flip
NU refine with C1 symmetry 4.52Å
GSFSC Resolution: 4.52Å
NU refine with C3 symmetry 4.38Å
GSFSC Resolution: 4.38Å
C
CH505TF Env-DH1744.1
90º
EMDB 72878
PDB 9YF6
204,251 particles
4.42 Å, C3 symmetry
Local Resolution (Å)
4.0
5.0
6.0
D
CH505TF Env-DH1745.1
90º
EMDB 72879
PDB 9YF7
227,623 particles
4.38 Å, C3 symmetry

**Figure S6.**

E

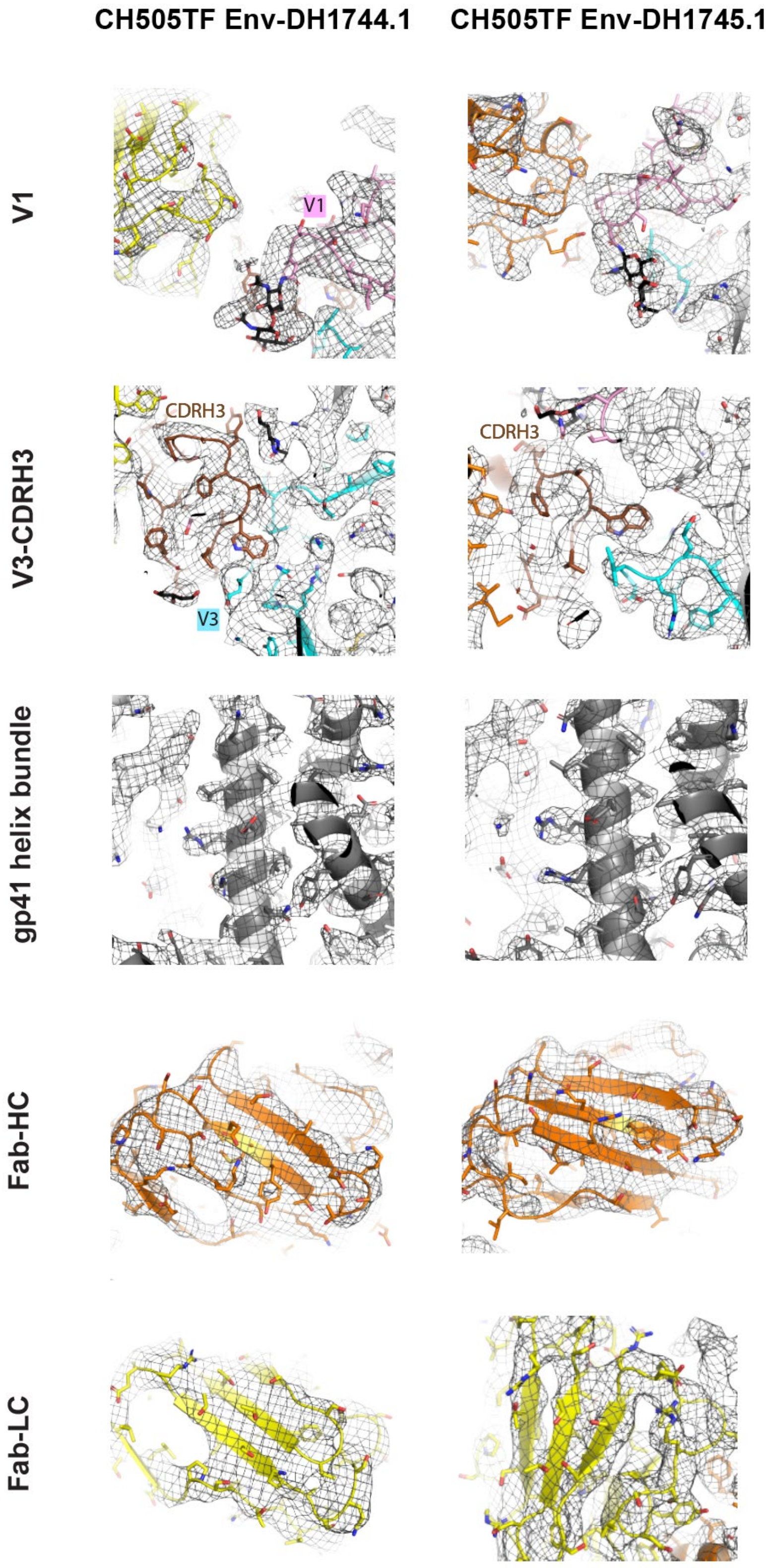

CH505TF Env-DH1744.1
CH505TF Env-DH1745.1
V1
V1
V3-CDRH3
CDRH3
CDRH3
V3
gp41 helix bundle
Fab-HC
Fab-LC


Figure S6.

**Figure S6. Cryo-EM data processing, map quality, and model fitting for CH505 TF Env complexes with DH1744.1 and DH1745.1.**

**(A, B) Cryo-EM data processing workflows for CH505 TF Env bound to (A) DH1744.1 and (B) DH1745.1.** Each workflow shows a representative cryo-EM micrograph, representative two-dimensional class averages, reconstructed maps generated by ab initio reconstruction and heterogeneous refinement, maps obtained after non-uniform (NU) refinement, and a Fourier shell correlation (FSC) curve from gold-standard refinement. The horizontal blue line indicates the FSC = 0.143 resolution criterion.

**(C, D) Refined cryo-EM maps colored according to local resolution for CH505 TF Env bound to (C) DH1744.1 and (D) DH1745.1.**

**(E) Zoomed-in views showing the fit of the atomic models to the cryo-EM maps.** The cryo-EM density is shown as a gray mesh, with the fitted models shown in cartoon representation and selected residues shown as sticks. The V1 loop is colored pink, V3 cyan, CDRH3 brown, gp41 dark gray, antibody heavy chains orange, and antibody light chains yellow.

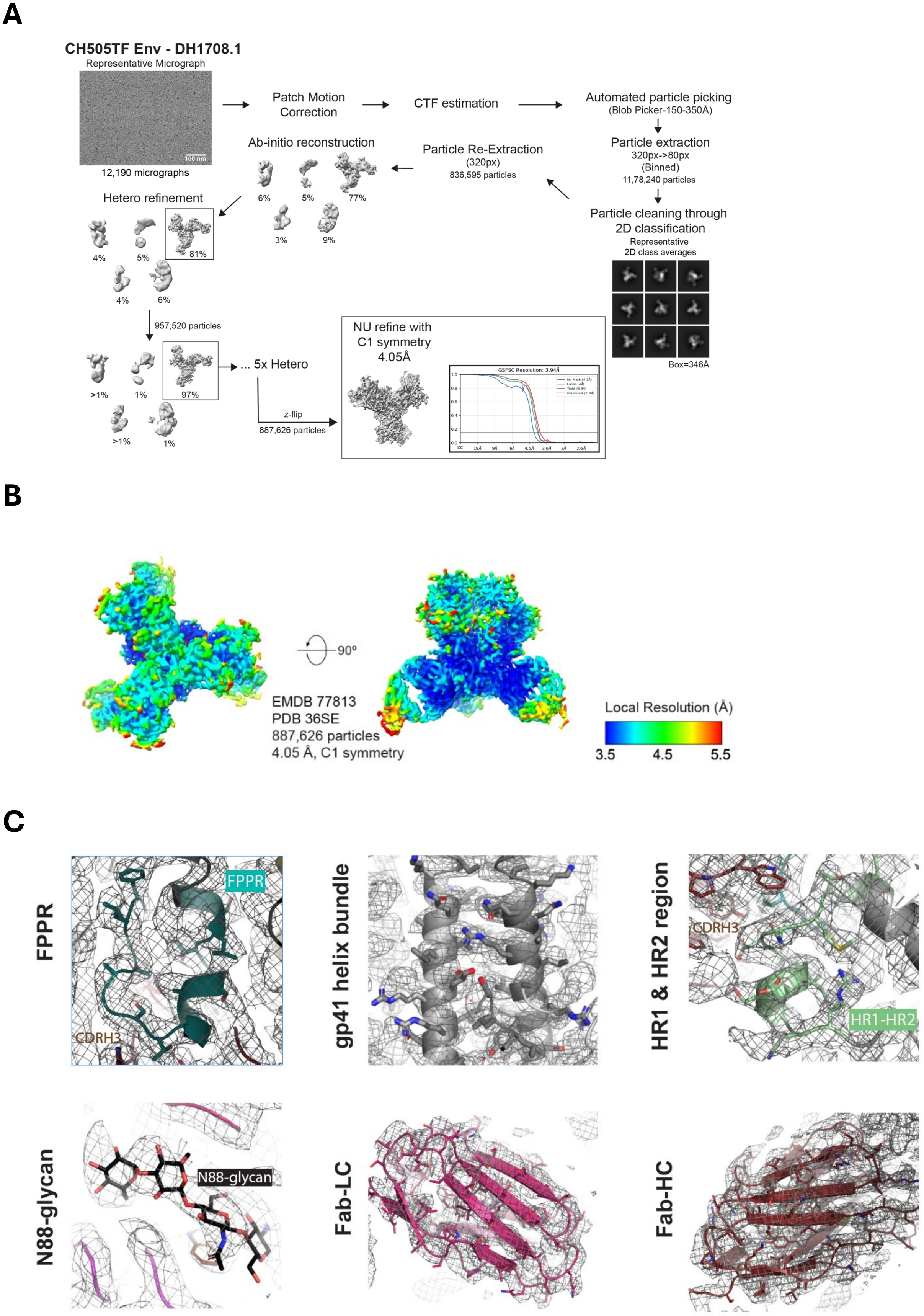
A
CH505TF Env - DH1708.1
Representative Micrograph
100 nm
12,190 micrographs
Patch Motion Correction
CTF estimation
Automated particle picking
(Blob Picker-150-350Å)
Particle extraction
320px->80px
(Binned)
11,78,240 particles
Particle cleaning through 2D classification
Representative 2D class averages
Box=346Å
Particle Re-Extraction
(320px)
836,595 particles
Ab-initio reconstruction
6%
5%
77%
3%
9%
Hetero refinement
4%
5%
81%
4%
6%
957,520 particles
>1%
1%
97%
>1%
1%
... 5x Hetero
z-flip
887,626 particles
NU refine with C1 symmetry 4.05Å
GSFSC Resolution: 3.94Å
B
90º
EMDB 77813
PDB 36SE
887,626 particles
4.05 Å, C1 symmetry
Local Resolution (Å)
3.5
4.5
5.5
C
FPPR
CDRH3
gp41 helix bundle
HR1 & HR2 region
HR1-HR2
N88-glycan
Fab-LC
Fab-HC

Figure S7.

**Figure S7. Cryo-EM data processing, map quality, and model fitting for CH505 TF Env complexes with DH1708.1.**

**(A) Data processing workflow of CH505TF bound to DH1708.1.** Workflow shows a representative cryo-EM micrograph, representative two-dimensional class averages, reconstructed maps generated by ab initio reconstruction and heterogeneous refinement, maps obtained after non-uniform (NU) refinement, and a Fourier shell correlation (FSC) curve from gold-standard refinement. The horizontal blue line indicates the FSC = 0.143 resolution criterion.

**(B) Refined cryo-EM maps colored according to local resolution for CH505 TF Env bound to DH1708.1.**

**(C) Zoomed-in views showing the fit of the atomic models to the cryo-EM maps.** The cryo-EM density is shown as a gray mesh, with the fitted models shown in cartoon representation and selected residues shown as sticks. The FPPR is colored deep teal, glycan at gp120 residue N88 is colored black, gp41 is colored dark gray, heavy chain is colored ruby, and light chain is colored warm pink.

**A**

| Cell Counts | |
|---|---|
| Sorted Cells | ~19,000 |
| GEX Cells | 11,562 |
| VDJ Cells | 11,111 |
| Functional 1:1 VDJ Cells* | 6,298 |
| Functional 1:1 VDJ + BEAM Cells | 6,296 |
| Functional 1:1 VDJ + BEAM + GEX Cells | 6,061 |

**B**

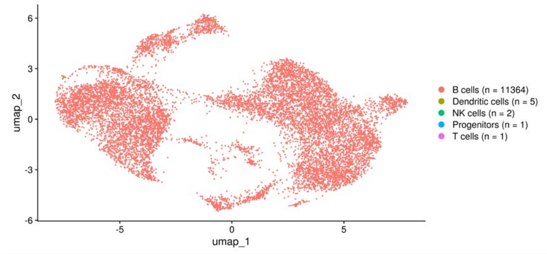


**C**

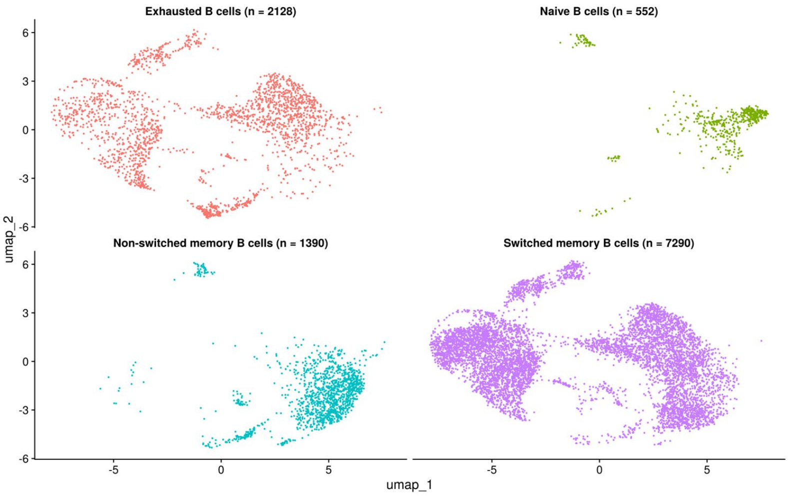


**D**

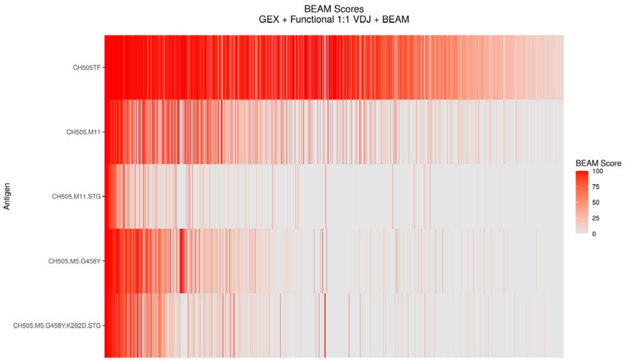


**E**

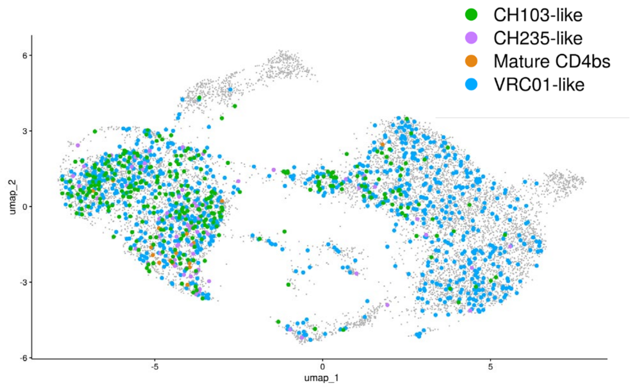


**F**

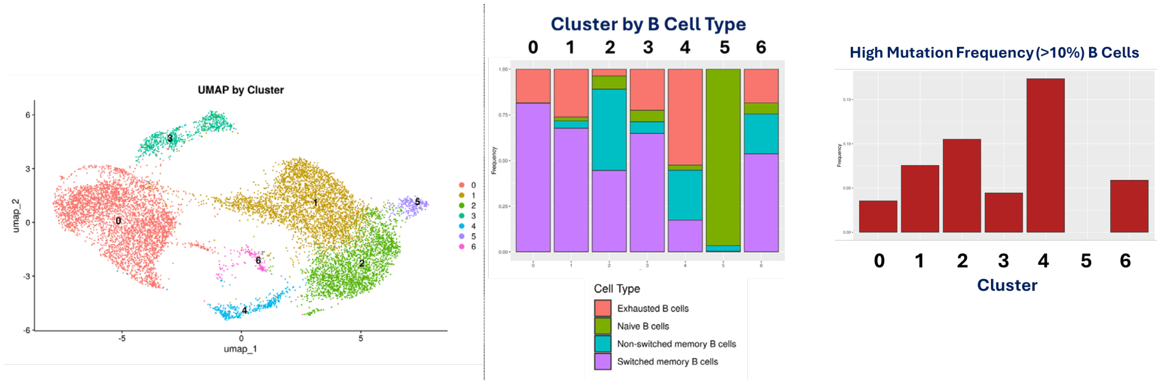


**Figure S8.**

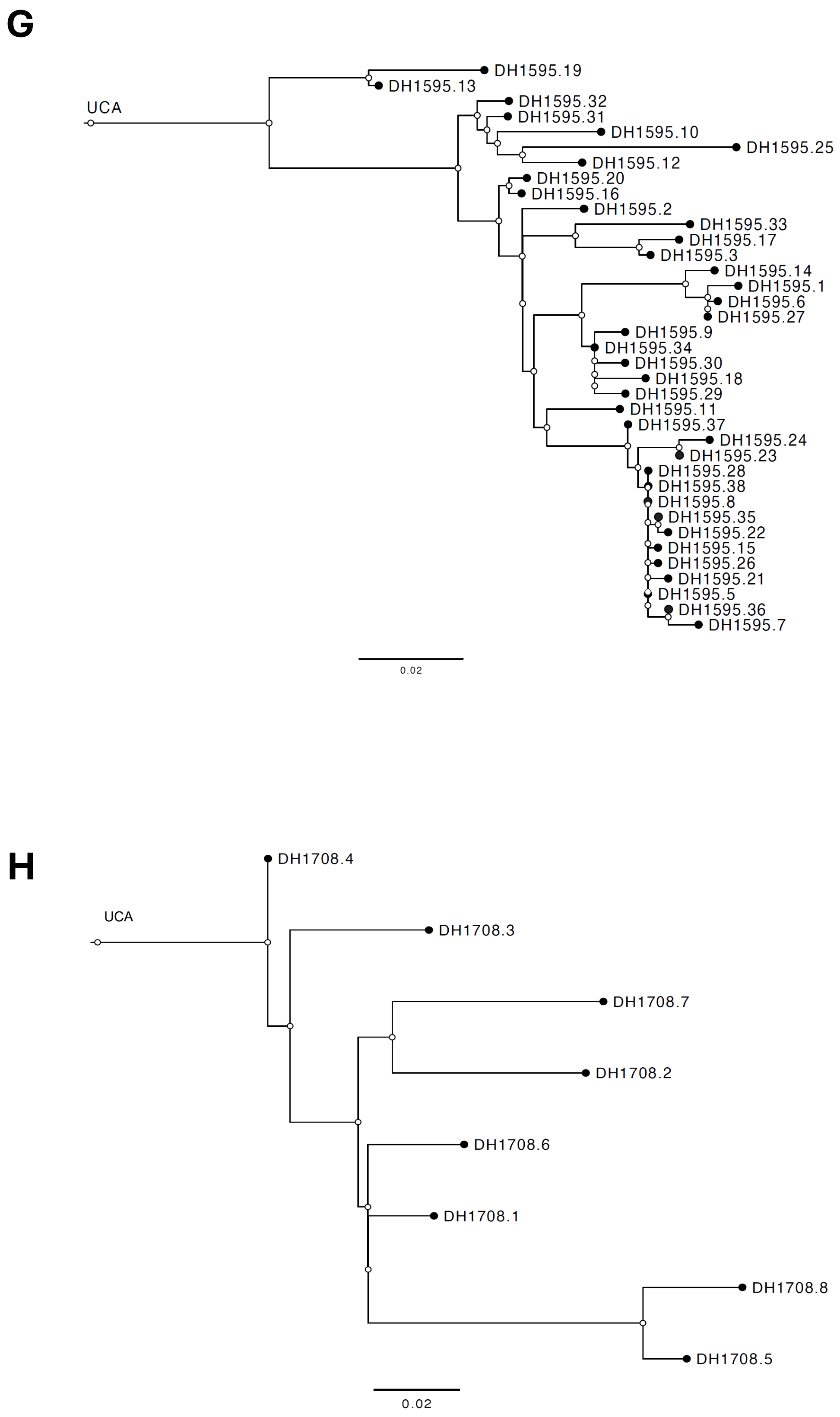
G
UCA
DH1595.19
DH1595.13
DH1595.32
DH1595.31
DH1595.10
DH1595.25
DH1595.12
DH1595.20
DH1595.16
DH1595.2
DH1595.33
DH1595.17
DH1595.3
DH1595.14
DH1595.1
DH1595.6
DH1595.27
DH1595.9
DH1595.34
DH1595.30
DH1595.18
DH1595.29
DH1595.11
DH1595.37
DH1595.24
DH1595.23
DH1595.28
DH1595.38
DH1595.8
DH1595.35
DH1595.22
DH1595.15
DH1595.26
DH1595.21
DH1595.5
DH1595.36
DH1595.7
0.02
H
DH1708.4
UCA
DH1708.3
DH1708.7
DH1708.2
DH1708.6
DH1708.1
DH1708.8
DH1708.5
0.02

**Figure S8.**

**Figure S8. Single-cell BEAM profiling and clonal expansion of neutralizing memory B-cell responses in elite responder participant UID123.**

**(A) Experimental overview and cell recovery.** The BEAM antigen panel comprised five CH505 Env SOSIP variants (CH505 TF, CH505.M11, CH505.M11-STG, CH505.M5.G458Y, and CH505.M5.G458Y.K282D-STG), each carrying dual barcode labels, alongside human serum albumin as a non-specific binding control. $CD20^+$ B cells were negatively enriched from the post-fourth immunization leukapheresis sample, and $IgG^+$ antigen-reactive cells were flow-sorted using a double-positive antigen strategy prior to 10x library preparation. From approximately 19,000 sorted cells, 6,061 cells with complete paired functional VDJ sequences, antigen barcode assignments, and matched transcriptomes were recovered for downstream analysis. Cell type classification by transcriptome confirmed that 11,364 of 11,373 recovered cells (99.9%) were B cells, with negligible contamination from non-B lineages (dendritic cells, n = 5; NK cells, n = 2; progenitors, n = 1; T cells, n = 1), validating the specificity of the enrichment strategy.

**(B) UMAP visualization of cell type composition following enrichment and sorting.** Cell type classification by transcriptome confirmed that 11,364 of 11,373 recovered cells (99.9%) were B cells, with negligible contamination from non-B lineages (dendritic cells, n = 5; NK cells, n = 2; progenitors, n = 1; T cells, n = 1), validating the specificity of the enrichment strategy.

**(C) UMAP transcriptome landscape with B cell subtype annotations, including switched memory, non-switched memory, exhausted, and naïve B cell populations.**

**(D) Per-cell BEAM score heatmap of the 2,082 cells exhibiting appreciable antigen reactivity (sum of BEAM scores > 10).** Each column represents a single cell; rows correspond to the five CH505 Env SOSIP panel antigens. BEAM score intensity (0–100) is shown on a red gradient scale. Distinct binding patterns across antigen rows reflect heterogeneous epitope specificities within the antigen-reactive population. BnAb-precursor-class specificities were defined by differential BEAM score thresholds anchored to canonical lineage binding signatures: CH103-like ($IgG^+$, CH505.M11 > 50 and CH505.M11-STG < 25), CH235-like ($IgG^+$, CH505.M5.G458Y > 50 and CH505.M5.G458Y.K282D-STG < 25), and VRC01-like ($IgG^+$, CH505TF > 50 and CH505.M5.G458Y < 25). By these criteria, 877 VRC01-like, 304 CH103-like, and 67 CH235-like B cells were identified. BnAb-like cell distributions are overlaid on the UMAP in panel **C.**

**(E) UMAP projection of all recovered B cells with BnAb-like specificities overlaid.** Gray dots represent cells without appreciable antigen reactivity or below BnAb-like score thresholds. Colored dots indicate cells classified as VRC01-like (blue, n = 877), CH103-like (green, n = 304), CH235-like (purple, n = 67), or Mature CD4bs (orange), as defined by the differential BEAM score thresholds described in panel **D**. BnAb-like cells are distributed across multiple transcriptional regions, indicating that antigen-reactive precursors span diverse B cell differentiation states rather than being confined to a single transcriptional cluster.

**(F) Distribution of highly mutated (somatic hypermutation frequency > 10%) and exhausted B cells across transcriptional clusters.** Highly mutated cells are disproportionately concentrated in Cluster 4, which is enriched for exhausted B cell signatures, suggesting sustained antigen-driven selection in a subset of clones by the post-fourth immunization timepoint.

**(G, H) BEAM expended clonal lineage trees of neutralizing antibody lineages from UID 123.** Each node represents a unique clonal member, and branch lengths indicate the degree of somatic hypermutation relative to the inferred UCA. **(G)** Lineage tree of the CD4bs-directed DH1595 lineage, incorporating 30 additional clonal members identified by BEAM. **(H)** Lineage tree of the FPPR-directed DH1708 lineage, incorporating seven additional clonal members identified by BEAM. DH1708 was the only FPPR-directed neutralizing lineage recovered among all HVTN 300 participants analyzed. The expanded lineage trees reveal more complete mutational trajectories from the inferred UCA toward CD4bs- and FPPR-directed neutralization than could be resolved from culture-derived sequences alone.

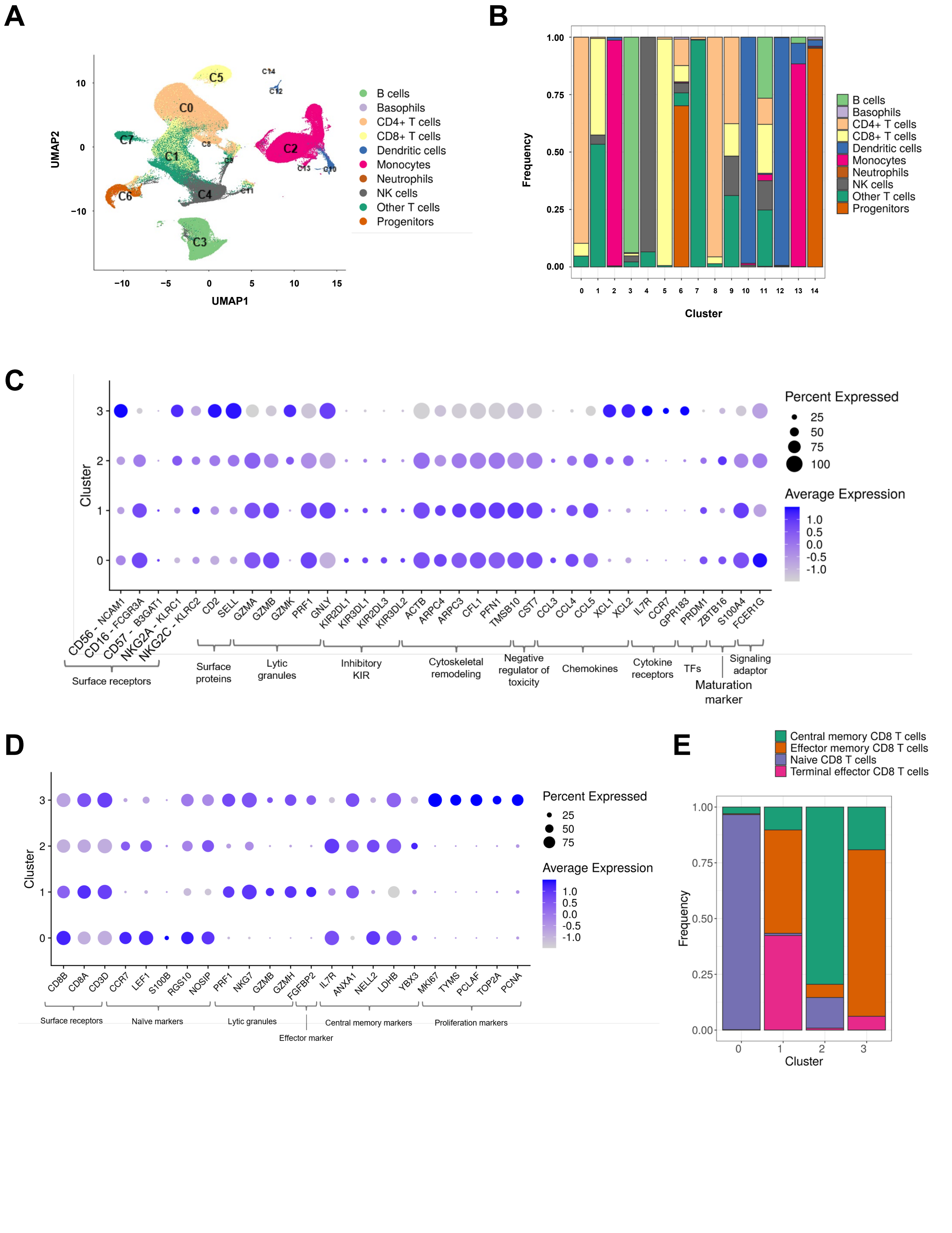
A
B
C
D
E
UMAP1
UMAP2
C0
C1
C2
C3
C4
C5
C6
C7
B cells
Basophils
CD4+ T cells
CD8+ T cells
Dendritic cells
Monocytes
Neutrophils
NK cells
Other T cells
Progenitors
Frequency
Cluster
Percent Expressed
Average Expression
Surface receptors
Surface proteins
Lytic granules
Inhibitory KIR
Cytoskeletal remodeling
Negative regulator of toxicity
Chemokines
Cytokine receptors
TFs
Signaling adaptor
Maturation marker
Naïve markers
Effector marker
Central memory markers
Proliferation markers
Central memory CD8 T cells
Effector memory CD8 T cells
Naive CD8 T cells
Terminal effector CD8 T cells

**Figure S9.**

**Figure S9. Annotation of PBMC, NK-cell and CD8+ T-cell clusters used for responder-associated single-cell analyses.**

**(A) UMAP visualization of total leukapheresis-derived PBMCs annotated by major immune-cell identity.** Cell-type annotations were assigned using SingleR annotation based on the Monaco immune cell reference (main labels, see Methods).

**(B) Composition of each unsupervised PBMC cluster shown in Fig. 6B, displayed as the proportional contribution of annotated immune-cell identities within each cluster.** This analysis supports the assignment of major PBMC clusters used for frequency comparisons between S365P-sensitive CDRH3-dominant CD4bs high and low responders in **Fig. 6C**.

**(C-D) Dot plots of canonical marker gene expression across NK cell (C) and CD8$^+$ T cell (D) subclusters used in Fig. 6D–G.** Dot size indicates the percentage of cells expressing each gene, and color indicates scaled average expression.

**(C)** These transcriptional profiles support the annotation of NK-cell subclusters, including NK.C0: CD56$^{dim}$CD16$^+$ mature, NK.C1: CD56$^{dim}$CD16$^+$ mature adaptive, NK.C2: CD56$^{dim}$ intermediate and NK.C3: CD56$^{bright}$ NK cells states.

**(D)** These transcriptional profiles support the annotation of CD8$^+$ T-cell subclusters as CD8.C0: naïve, CD8.C1: central memory, CD8.C2: effector memory, and CD8.C3: terminal effector CD8$^+$ T-cell states.

**(E) Composition of each CD8$^+$ T-cell subcluster shown in (D), displayed as the proportional contribution of SingleR/Monaco-annotated CD8$^+$ T-cell states within each cluster.** Cluster 0 was dominated by naïve CD8$^+$ T cells, cluster 1 contained a mixture of effector memory and terminal effector CD8$^+$ T cells, and clusters 2 and 3 showed higher proportions of central memory and effector memory CD8$^+$ T cells. These annotations, together with cross-referencing known marker expression in (D) to PBMC and CD8+ T cell single-cell atlases (see Methods), were used to interpret CD8$^+$ T-cell states associated with responder-group differences.
