## Supplementary material for "Priming of Multiple HIV Neutralizing B Cell Precursors in Humans": Table S1-S3 and Data S1-S2: Table_S1_V3.docx

**Table S1.** Participant-level counts and frequencies of CH505.TF double-positive memory B cell cultures, CH505-reactive cultures, and epitope-defined neutralizing antibody classes.

| **Participant UID** | **No. of CH505.TF DP Memory B Cell Cultures Plated** | **No. of B Cell Cultures Grown** | **No. of CH505-Binding Cultures (ELISA)** | **No. of CH505 Neutralizing Cultures (TZM-bl)** | **No. of CH505.TF Neutralizing Cultures (TZM-bl)** | **No. of V3-Targeting Neutralizing Cultures** | **No. of CD4bs Neutralizing Cultures** | **No. of S365P-sensitive CD4bs Neutralizing Cultures** |
| --- | --- | --- | --- | --- | --- | --- | --- | --- |
| 67 | 6598 | 3380 | 1931 | 162 | 35 | 14 | 6 | 6 |
| 86 | 2490 | 1126 | 884 | 201 | 20 | 12 | 1 | 0 |
| 123 | 7197 | 1181 | 1034 | 166 | 55 | 19 | 21 | 9 |
| 185 | 4992 | 1330 | 1027 | 112 | 30 | 15 | 14 | 14 |
| 192 | 2880 | 1180 | 994 | 71 | 23 | 9 | 4 | 4 |
| 283 | 2833 | 846 | 581 | 119 | 29 | 12 | 0 | 0 |
| 332 | 2782 | 1435 | 1061 | 342 | 122 | 86 | 10 | 7 |
| 440 | 2182 | 1165 | 727 | 154 | 20 | 11 | 5 | 3 |
| 468 | 1664 | 863 | 537 | 99 | 7 | 5 | 1 | 1 |
| 469 | 4512 | 1633 | 1313 | 232 | 74 | 28 | 29 | 29 |
| 475 | 4032 | 771 | 652 | 58 | 14 | 13 | 0 | 0 |
| **Sum.** | 42162 | 14910 | 10741 | 1716 | 429 | 224 | 91 | 73 |

| **Participant UID** | **Freq. of CH505.TF DP Memory B Cells (Flow cytometry)** | **Cloning Efficiency** | **Freq. of CH505-Binding Cultures (ELISA)** | **Freq. of CH505 Neutralizing Cultures (TZM-bl)** | **Freq. of CH505.TF Neutralizing Cultures (TZM-bl)** | **Freq. of V3-Targeting Neutralizing Cultures** | **Freq. of CD4bs Neutralizing Cultures** | **Freq. of S365P-sensitive CD4bs Neutralizing Cultures** |
| --- | --- | --- | --- | --- | --- | --- | --- | --- |
| 67 | 1.33% | 51.23% | 0.76% | 0.0637% | 0.0138% | 0.0055% | 0.0024% | 0.0024% |
| 86 | 0.70% | 45.22% | 0.55% | 0.1250% | 0.0124% | 0.0075% | 0.0006% | 0.0000% |
| 123 | 2.07% | 16.41% | 1.81% | 0.2910% | 0.0964% | 0.0333% | 0.0368% | 0.0158% |
| 185 | 1.33% | 26.64% | 1.03% | 0.1120% | 0.0300% | 0.0150% | 0.0140% | 0.0140% |
| 192 | 0.70% | 40.97% | 0.59% | 0.0421% | 0.0136% | 0.0053% | 0.0024% | 0.0024% |
| 283 | 0.59% | 29.86% | 0.41% | 0.0830% | 0.0202% | 0.0084% | 0.0000% | 0.0000% |
| 332 | 0.40% | 51.58% | 0.30% | 0.0953% | 0.0340% | 0.0240% | 0.0028% | 0.0020% |
| 440 | 1.09% | 53.39% | 0.68% | 0.1441% | 0.0187% | 0.0103% | 0.0047% | 0.0028% |
| 468 | 0.50% | 51.86% | 0.31% | 0.0574% | 0.0041% | 0.0029% | 0.0006% | 0.0006% |
| 469 | 0.84% | 36.19% | 0.68% | 0.1193% | 0.0381% | 0.0144% | 0.0149% | 0.0149% |
| 475 | 0.80% | 19.12% | 0.68% | 0.0602% | 0.0145% | 0.0135% | 0.0000% | 0.0000% |
| **Median** | 0.80% | 38.41% | 0.68% | 0.0953% | 0.0187% | 0.0103% | 0.0028%* | 0.0026%* |
| **1 in x Bmem** | 125 |  | 148 | 1049 | 5344 | 9716 | 35875 | 38612 |

DP, double-positive; ELISA, enzyme-linked immunosorbent assay; TZM-bl, Neutralizing Antibody Assay for HIV-1 in TZM-bl Cells; V3, variable loop 3; CD4bs, CD4-binding site;

Freq., frequency; Bmem, memory B cell.

* Median frequency was calculated among participants with non-zero values.
