## Supplementary material for "Priming of Multiple HIV Neutralizing B Cell Precursors in Humans": Table S1-S3 and Data S1-S2: Table_S2_V2.docx

**Table S2.** NSEM epitope classification of participant-derived monoclonal antibodies from HVTN300.

| **DH Number** | **Participant UID** | **VH Gene** | **VL Gene** | **NSEM epitope classification** |
| --- | --- | --- | --- | --- |
| DH1530.1 | 469 | IGHV3-11*01 | IGLV3-1*01 | CD4bs (CH103-like) |
| DH1530.10 | 469 | IGHV3-11*01 | IGLV3-1*01 | CD4bs (CH103-like) |
| DH1530.11 | 469 | IGHV3-11*01 | IGLV3-1*01 | CD4bs (CH103-like) |
| DH1530.12 | 469 | IGHV3-11*01 | IGLV3-1*01 | CD4bs (CH103-like) |
| DH1530.3 | 469 | IGHV3-11*01 | IGLV3-1*01 | CD4bs (CH103-like) |
| DH1530.4 | 469 | IGHV3-11*01 | IGLV3-1*01 | CD4bs (CH103-like) |
| DH1530.5 | 469 | IGHV3-11*01 | IGLV3-1*01 | CD4bs (CH103-like) |
| DH1530.6 | 469 | IGHV3-11*01 | IGLV3-1*01 | CD4bs (CH103-like) |
| DH1530.8 | 469 | IGHV3-11*01 | IGLV3-1*01 | CD4bs (CH103-like) |
| DH1530.9 | 469 | IGHV3-11*01 | IGLV3-1*01 | CD4bs (CH103-like) |
| DH1541.1 | 185 | IGHV3-23*01 | IGLV3-1*01 | CD4bs (CH103-like) |
| DH1595.1 | 123 | IGHV1-2*02 | IGLV3-25*i01 | CD4bs |
| DH1595.2 | 123 | IGHV1-2*02 | IGLV3-25*i01 | CD4bs |
| DH1595.3 | 123 | IGHV1-2*02 | IGLV3-25*i01 | CD4bs |
| DH1708.1 | 123 | IGHV1-2*02 | IGKV1D-13*01 | gp120/gp41 interface |
| DH1716.1 | 332 | IGHV3-33*01 | IGKV2-28*01 | V3 |
| DH1716.2 | 332 | IGHV3-33*01 | IGKV2-28*01 | V3 |
| DH1716.3 | 332 | IGHV3-33*01 | IGKV2-28*01 | V3 |
| DH1716.4 | 332 | IGHV3-33*01 | IGKV2-28*01 | V3 |
| DH1717.1 | 332 | IGHV3-11*06 | IGLV2-23*02 | CD4bs |
| DH1717.2 | 332 | IGHV3-11*06 | IGLV2-23*02 | CD4bs |
| DH1718.1 | 332 | IGHV3-23*01 | IGLV1-40*01 | V3 |
| DH1718.2 | 332 | IGHV3-23*01 | IGKV3-20*01 | V3 |
| DH1718.5 | 332 | IGHV3-23*01 | IGLV1-40*01 | V3 |
| DH1718.6 | 332 | IGHV3-23*01 | IGLV1-40*01 | V3 |
| DH1719.1 | 332 | IGHV3-30*18 | IGKV1-12*01 | V3 |
| DH1719.2 | 332 | IGHV3-30*18 | IGKV1-12*01 | V3 |
| DH1719.3 | 332 | IGHV3-30*18 | IGKV1-12*01 | V3 |
| DH1720.1 | 332 | IGHV3-15*01 | IGKV3-15*01 | V3 |
| DH1720.2 | 332 | IGHV3-15*01 | IGKV3-15*01 | V3 |
| DH1721.2 | 332 | IGHV1-18*01 | IGKV3-20*01 | CD4bs |
| DH1721.3 | 332 | IGHV1-18*01 | IGKV3-20*01 | CD4bs |
| DH1721.4 | 332 | IGHV1-18*01 | IGKV3-20*01 | CD4bs |
| DH1721.5 | 332 | IGHV1-18*01 | IGKV3-20*01 | CD4bs |
| DH1722.1 | 332 | IGHV3-33*01 | IGKV1-39*01 | V3 |
| DH1722.2 | 332 | IGHV3-33*06 | IGKV1-39*01 | V3 |
| DH1723.1 | 123 | IGHV2-5*02 | IGKV1-12*01 | CD4bs |
| DH1723.2 | 123 | IGHV2-5*02 | IGKV1-12*01 | CD4bs |
| DH1725.1 | 67 | IGHV3-33*01 | IGKV3-11*01 | V3 |
| DH1725.2 | 67 | IGHV3-33*01 | IGKV3-11*01 | V3 |
| DH1726.1 | 185 | IGHV3-30-3*01 | IGLV7-46*01 | V3 |
| DH1726.2 | 185 | IGHV3-30-3*01 | IGLV7-46*01 | V3 |
| DH1726.3 | 185 | IGHV3-30-3*01 | IGLV7-46*01 | V3 |
| DH1727.1 | 185 | IGHV3-33*01 | IGKV3-11*01 | V3 |
| DH1727.2 | 185 | IGHV3-33*01 | IGKV3-11*01 | V3 |
| DH1727.3 | 185 | IGHV3-33*01 | IGKV3-11*01 | V3 |
| DH1727.4 | 185 | IGHV3-33*01 | IGKV3-11*01 | V3 |
| DH1728.1 | 185 | IGHV3-30*18 | IGKV3-15*01 | V3 |
| DH1728.2 | 185 | IGHV3-30*18 | IGKV3-15*01 | V3 |
| DH1729.2 | 192 | IGHV1-69*01 | IGLV2-14*03 | V3 |
| DH1730.1 | 192 | IGHV3-33*01 | IGKV2-28*01 | V3 |
| DH1730.2 | 192 | IGHV3-33*01 | IGKV2-28*01 | V3 |
| DH1731.1 | 469 | IGHV3-15*01 | IGKV1-39*01 | V3 |
| DH1731.3 | 469 | IGHV3-15*01 | IGKV1-39*01 | V3 |
| DH1731.4 | 469 | IGHV3-15*01 | IGKV1-39*01 | V3 |
| DH1731.5 | 469 | IGHV3-15*01 | IGKV1-39*01 | V3 |
| DH1732.1 | 86 | IGHV1-69*06 | IGLV2-23*02 | V3 |
| DH1732.2 | 86 | IGHV1-69*01 | IGLV2-23*02 | V3 |
| DH1733.1 | 440 | IGHV3-23*01 | IGLV1-40*01 | V3 |
| DH1734.1 | 192 | IGHV3-21*01 | IGKV3-20*01 | CD4bs |
| DH1734.2 | 192 | IGHV3-21*01 | IGKV3-20*01 | CD4bs |
| DH1735.1 | 67 | IGHV3-21*01 | IGKV1-5*03 | CD4bs |
| DH1735.2 | 67 | IGHV3-21*01 | IGKV1-5*03 | CD4bs |
| DH1737.1 | 475 | IGHV1-2*02 | IGKV1-5*01 | V3 |
| DH1737.2 | 475 | IGHV1-2*02 | IGKV1-5*01 | V3 |
| DH1737.3 | 475 | IGHV1-2*02 | IGKV1-5*01 | V3 |
| DH1738.1 | 86 | IGHV3-74*01 | IGKV1-39*01 | V3 |
| DH1739.2 | 468 | IGHV3-33*06 | IGKV3-20*01 | V3 |
| DH1739.3 | 468 | IGHV3-33*06 | IGKV3-20*01 | V3 |
| DH1744.1 | 123 | IGHV1-69*10 | IGKV3-20*01 | V3 |
| DH1745.1 | 469 | IGHV3-23*01 | IGKV3-20*01 | V3 |
| DH1746 | 469 | IGHV1-2*02 | IGKV3-15*01 | V3 |
| DH1747.1 | 469 | IGHV3-30*18 | IGKV2-28*01 | V3 |
| DH1748.1 | 469 | IGHV3-11*01 | IGKV1-NL1*01 | V3 |
| DH1749 | 469 | IGHV1-2*02 | IGKV3-20*01 | V3 |
| DH1754.1 | 475 | IGHV3-23*01 | IGKV3-20*01 | V3 |
| DH1755 | 475 | IGHV3-33*01 | IGKV1-5*03 | V3 |
| DH1756 | 475 | IGHV1-69*12 | IGKV3-11*01 | V3 |
| DH1757 | 475 | IGHV3-33*01 | IGKV1-39*01 | V3 |
| DH1758 | 475 | IGHV3-15*01 | IGLV2-14*01 | V3 |
| DH1759.1 | 123 | IGHV2-5*02 | IGKV4-1*01 | CD4bs |
| DH1760 | 123 | IGHV1-18*01 | IGKV1D-13*01 | V3 |
| DH1761.1 | 123 | IGHV1-18*01 | IGKV1-12*01 | V3 |
| DH1762 | 123 | IGHV3-23*01 | IGLV1-40*01 | V3 |
| DH1763 | 123 | IGHV3-30*02 | IGLV2-14*01 | V3 |
| DH1764 | 185 | IGHV3-33*01 | IGKV3-20*01 | V3 |
| DH1765 | 185 | IGHV1-2*02 | IGKV1-5*03 | V3 |
| DH1766 | 185 | IGHV3-33*01 | IGKV2-28*01 | V3 |
| DH1767 | 185 | IGHV3-23*01 | IGLV1-40*01 | V3 |
| DH1768.1 | 185 | IGHV3-33*01 | IGLV2-11*01 | CD4bs |
| DH1770.1 | 283 | IGHV1-69*01 | IGKV2-28*01 | V3 |
| DH1771 | 283 | IGHV3-11*06 | IGLV1-51*01 | V3 |
| DH1772 | 192 | IGHV1-69*12 | IGLV1-51*01 | V3 |
| DH1773 | 192 | IGHV3-33*01 | IGLV2-8*01 | V3 |
| DH1777.1 | 86 | IGHV4-59*01 | IGKV3-20*01 | CD4bs |
| DH1779 | 86 | IGHV4-61*02 | IGKV1-39*01 | V3 |
| DH1781 | 468 | IGHV1-69*01 | IGKV3-20*01 | V3 |
| DH1782 | 468 | IGHV3-15*01 | IGKV1-39*01 | V3 |
| DH1783 | 440 | IGHV3-48*03 | IGLV3-1*01 | CD4bs (CH103-like) |
| DH1785 | 440 | IGHV3-33*01 | IGKV2-28*01 | V3 |
| DH1786.1 | 440 | IGHV5-51*01 | IGLV3-25*i01 | CD4bs |
| DH1787 | 283 | IGHV4-39*01 | IGKV3-11*01 | V3 |
| DH1789 | 283 | IGHV3-30*18 | IGKV1-9*01 | V3 |
| DH1791 | 283 | IGHV3-11*06 | IGKV1-9*01 | V3 |
| DH1792 | 283 | IGHV1-69*17 | IGKV3-11*01 | V3 |
| DH1794 | 283 | IGHV3-49*05 | IGKV1D-33*01 | V3 |
| DH1795.1 | 283 | IGHV1-69*06 | IGKV3-20*01 | V3 |
| DH1796 | 283 | IGHV3-15*01 | IGKV2-28*01 | V3 |
| DH1797 | 192 | IGHV1-2*02 | IGKV1-5*03 | V3 |
| DH1799 | 192 | IGHV3-33*01 | IGKV2-28*01 | V3 |
| DH1801 | 192 | IGHV3-21*01 | IGKV6D-21*02 | V3 |
| DH1802.1 | 67 | IGHV3-33*01 | IGKV2-28*01 | V3 |
| DH1803 | 67 | IGHV3-30-3*01 | IGKV3-20*01 | V3 |
| DH1804 | 67 | IGHV3-30*18 | IGKV1D-12*01 | V3 |
| DH1809 | 468 | IGHV3-30*02 | IGKV3-20*01 | V3 |
| DH1812 | 440 | IGHV3-30*02 | IGKV1-5*03 | CD4bs |
| DH1813 | 440 | IGHV3-7*01 | IGKV2-28*01 | V3 |
| DH1814 | 440 | IGHV3-30*18 | IGLV1-40*01 | V3 |
| DH1816 | 440 | IGHV3-30*02 | IGKV2-24*01 | V3 |
| DH1817.1 | 440 | IGHV3-15*01 | IGKV3-20*01 | V3 |
| DH1818 | 440 | IGHV1-2*02 | IGKV1-5*03 | V3 |
| DH1819 | 332 | IGHV3-15*07 | IGLV7-46*01 | V3 |
| DH1820 | 123 | IGHV3-43*01 | IGKV3-15*01 | V3 |
| DH1821 | 123 | IGHV3-15*01 | IGLV1-40*01 | V3 |
| DH1822 | 123 | IGHV1-2*02 | IGKV1-5*03 | V3 |
| DH1823.1 | 123 | IGHV4-39*01 | IGKV1-5*03 | V3 |
| DH1824 | 123 | IGHV3-7*01 | IGKV3-15*01 | V3 |
| DH1825.1 | 123 | IGHV3-15*01 | IGKV6D-21*01 | V3 |
| DH1826.1 | 123 | IGHV3-33*06 | IGKV1D-13*02 | V3 |
| DH1827.1 | 469 | IGHV3-30*18 | IGKV6D-21*01 | V3 |
| DH1829 | 469 | IGHV4-59*01 | IGKV3-20*01 | V3 |
| DH1832.1 | 469 | IGHV1-69*01 | IGLV2-14*01 | V3 |
| DH1834 | 332 | IGHV1-46*01 | IGKV3-20*01 | V3 |
| DH1837 | 67 | IGHV3-74*01 | IGLV3-10*01 | CD4bs |
| DH1838 | 67 | IGHV1-69*06 | IGLV7-46*04 | V3 |
| DH1839.1 | 67 | IGHV1-69*01 | IGLV2-8*01 | V3 |
| DH1840.1 | 67 | IGHV3-33*01 | IGKV2-28*01 | V3 |
