## Supplementary material for "Priming of Multiple HIV Neutralizing B Cell Precursors in Humans": Table S1-S3 and Data S1-S2: Table_S3.docx

**Table S3.** Heterologous neutralization IC50 values for DH1777 lineage mAbs.

| **DH numbers** | **IC50 (μg/mL)** | | | | | | | | | |
| --- | --- | --- | --- | --- | --- | --- | --- | --- | --- | --- |
|  | **001428-2.42** | **CNE19** | **CNE20** | **CNE8** | **3365.v2.c20** | **6480.v4.c25** | **CNE58** | **R2184.c04** | **25710-2.43** | **ZM247v1(Rev-)** |
| **DH1777.1** | 28 | 26.7 | 82 | 14 | 38 | 12 | 13.6 | 7.8 | >100.0 | 54 |
| **DH1777.2** | 31 | 9.8 | 42 | 81 | 13 | 10 | 2.4 | 6.6 | 94.0 | 43 |
| **DH1777.3** | 32 | 22 | >100.0 | >100.0 | 26 | 14 | 17 | 8.6 | >100.0 | >100.0 |
| **DH1777.4** | 47 | 14 | 65 | >100.0 | 29 | 15 | 4.2 | 6.5 | >100.0 | 72 |
| **DH1777.5** | >100.0 | >100.0 | >100.0 | >100.0 | >100.0 | >100.0 | >100.0 | >100.0 | >100.0 | >100.0 |
| **DH1777.6** | 32 | 10 | 21 | 46 | 26 | 15 | 15 | 8.0 | >100.0 | 82 |
| **DH1777.7** | >100.0 | >100.0 | >100.0 | >100.0 | >100.0 | 66 | >100.0 | >100.0 | >100.0 | >100.0 |
| **DH1777.8** | 41 | 35 | 57 | 92 | 58 | 21 | 22 | 16 | >100.0 | >100.0 |
| **DH1777.9** | 77 | 17 | 97 | >100.0 | 32 | 18 | 6.2 | 18 | >100.0 | 78 |
| **DH1777.10** | >100.0 | 65 | >100.0 | >100.0 | >100.0 | 50 | >100.0 | >100.0 | >100.0 | >100.0 |
| **DH1777.11** | 44 | 13 | 87 | >100.0 | >100.0 | 27 | 5.7 | 25 | >100.0 | 42 |

**Footnote:** IC50 values are reported in μg/mL. Heterologous HIV-1 Env pseudoviruses were generated using the SG3Δenv backbone and 293T/17 producer cells. Values of >100.0 indicate no detectable neutralization at the highest antibody concentration tested.
